# Multi-organ AI Endophenotypes Chart the Heterogeneity of Pan-disease in the Brain, Eye, and Heart

**DOI:** 10.1101/2025.08.09.25333350

**Authors:** The MULTI consortium, Aleix Boquet-Pujadas, Filippos Anagnostakis, Zhijian Yang, Ye Ella Tian, Michael R. Duggan, Guray Erus, Dhivya Srinivasan, Cassandra M. Joynes, Wenjia Bai, Praveen J. Patel, Keenan A. Walker, Andrew Zalesky, Christos Davatzikos, Junhao Wen

## Abstract

Disease heterogeneity and commonality pose significant challenges to precision medicine, as traditional approaches frequently focus on single disease entities and overlook shared mechanisms across conditions^1^. Inspired by pan-cancer^2^ and multi-organ research^3^, we introduce the concept of “pan-disease” to investigate the heterogeneity and shared etiology in brain, eye, and heart diseases. Leveraging individual-level data from 129,340 participants, as well as summary-level data from the MULTI consortium, we applied a weakly-supervised deep learning model (Surreal-GAN^4,5^) to multi-organ imaging, genetic, proteomic, and RNA-seq data, identifying 11 AI-derived biomarkers – called Multi-organ AI Endophenotypes (MAEs) – for the brain (Brain 1–6), eye (Eye 1–3), and heart (Heart 1–2), respectively. We found Brain 3 to be a risk factor for Alzheimer’s disease (AD) progression and mortality, whereas Brain 5 was protective against AD progression. Crucially, in data from an anti-amyloid AD drug (solanezumab^6^), heterogeneity in cognitive decline trajectories was observed across treatment groups. At week 240, patients with lower brain 1-3 expression had slower cognitive decline, whereas patients with higher expression had faster cognitive decline. A multi-layer causal pathway pinpointed Brain 1 as a mediational endophenotype^7^ linking the FLRT2 protein to migraine, exemplifying novel therapeutic targets and pathways. Additionally, genes associated with Eye 1 and Eye 3 were enriched in cancer drug-related gene sets with causal links to specific cancer types and proteins. Finally, Heart 1 and Heart 2 had the highest mortality risk and unique medication history profiles, with Heart 1 showing favorable responses to antihypertensive medications and Heart 2 to digoxin treatment. The 11 MAEs provide novel AI dimensional representations for precision medicine and highlight the potential of AI-driven patient stratification for disease risk monitoring, clinical trials, and drug discovery.

## Main

Disease heterogeneity within single disease entities, cross-disease commonalities, and etiologic overlap present significant challenges for precision medicine^8^. In the case of single disease entities, artificial intelligence (AI) has been applied to brain magnetic resonance imaging (MRI) data, which revealed distinct disease subtypes or dimensions, highlighting the neuroanatomical heterogeneity of brain disorders, such as Alzheimer’s disease (AD)^9,10^. On the other hand, recent findings from genetics and transcriptomics have unraveled overlapping molecular and neuropathological signatures across brain disorders, highlighting their shared biological underpinnings^11,12^. We argue that AI holds great potential to jointly model human aging and disease concurrently from the two abovementioned perspectives to advance precision medicine.

Addressing disease heterogeneity and transcending traditional classifications, such as those based on International Classification of Diseases (ICD) codes, may reveal new insights into aging and disease. Recent initiatives have focused on identifying transdiagnostic disease subtypes and dimensions, particularly within brain disorders like psychiatric (depression and psychosis^13^) and developmental conditions^14^. This has provided new avenues for understanding polygenic and etiologically multi-faceted diseases. In our recent study, by leveraging a weakly-supervised learning framework^15^ (e.g., Surreal-GAN^4,5^), we derived 9 AI-derived endophenotypes to capture the within-disease heterogeneity and cross-disease similarities in 4 brain diseases^16^, including autism spectrum disorder, schizophrenia, late-life depression, and AD, as well as aging^5^. Critically, the 9 AI-derived phenotypes were generated within a specific disease entity, such as AD. The observed neuroanatomical overlap underscores the need for new approaches that derive disease subtypes or dimensions that transcend traditional disease boundaries, enabling a more integrated understanding of disease mechanisms.

Modeling human aging and disease necessitates a comprehensive, multi-scale approach spanning both spatial and temporal granularities^17^. Recent advances in multi-organ research^18,19,20,3,21^ have opened up new possibilities for holistic modeling of human aging and disease. For example, Tian et al.^22^ used machine learning (ML) to calculate the biological age gap (BAG) in nine organ systems, linking these biomarkers to lifestyle and mortality in the UK Biobank (UKBB). In subsequent analyses, Wen et al.^21,23^ further investigated the genetic architecture of these multi-organ BAGs. Similarly, multi-omics approaches, such as combining brain MRI data with genetics and proteomics, offer enhanced diagnostic precision and granularity. For instance, Yang et al. recently showed that incorporating genetic data into imaging-based models (e.g., Gene-SGAN^24^) enhances disease subtyping accuracy and outperforms approaches developed solely on imaging data (e.g., Smile-GAN^9^). In another study, we established the brain-heart-eye axis using imaging-derived phenotypes (IDPs) from *in vivo* imaging, genetic, and proteomic data of these 3 organs, demonstrating that no organ system is an island^25^. Taken together, this evidence underscores the importance of integrating multi-organ, multi-omics data across multiple human organ systems and omics data types to model disease heterogeneity.

Dissecting disease heterogeneity and commonality using AI in combination with *in vivo* imaging techniques (e.g., MRI for the brain^26^ and heart^27^, and optical coherence tomography (OCT)^28–30^ for the eye) also aligns with the endophenotype hypothesis^7^. Originally proposed in psychiatric genetics, this hypothesis suggests that intermediate phenotypes – like those identified through imaging – bridge the causal pathway from genetic variants to disease endpoints (DEs) and clinical symptoms. Two potential models of endophenotypes have been proposed: *i*) the liability-index model, wherein genetics simultaneously influences both DEs and endophenotypes (i.e., horizontal pleiotropy), and *ii*) the mediational model, in which genetic effects on DEs are (exclusively) mediated through intermediate phenotypes (i.e., vertical pleiotropy). While the former phenomenon is ubiquitous in population genetics, the latter holds significant potential for identifying causal and actionable target genes for therapeutic development and intervention. In prior work^31^, we demonstrated that AI-derived subtypes of schizophrenia exhibited lower polygenicity and weaker negative purifying selection compared to traditional case-control diagnoses of schizophrenia, providing proof-of-concept evidence to support one assumption of this hypothesis. That is, the endophenotype is less polygenic than the DE itself, thereby being closer to the underlying genetic etiology of the disease^7^.

Here, we introduced 6, 3, and 2 multi-organ AI endophenotypes (MAEs) to digitize individual-level morphological heterogeneity in brain, eye, and heart diseases to address the outlined challenges. Intuitively, each organ-specific MAE captures a distinctive imaging pattern, reflecting diverse disease spectrums within each organ system. This was achieved by applying 3 separate Surreal-GAN^4,5^ models to organ-specific imaging data of the brain, eye, and heart consolidated via the ongoing MULTI consortium (refer to **Method 1** and **Supplementary eTable 1**). Subsequently, we linked the 11 MAEs to other omic data, including genetics, proteomics, and RNA-seq data, both at individual and summary levels. The scientific advancements are threefold. First, drawing inspiration from pan-cancer research^2^, which explores shared mechanisms across different cancer types, we introduce the concept of “pan-disease” to address the complexity of disease heterogeneity and commonalities across conditions/diseases in each organ. Second, recognizing that no organ system functions in isolation^25^, we leverage large-scale, multi-organ, and multi-omics biomedical data to capture the morphological heterogeneity of the pan-disease of the three organs. Lastly, we presented putative evidence to support the endophenotype hypothesis^7^. In particular, we demonstrate how the mediational model of the endophenotype hypothesis can be validated in certain cases, offering novel insights that could inform drug discovery and personalized therapeutic strategies.

We present the definition of pan-disease, our analytic approaches, the network architecture of Surreal-GAN, the datasets used, and the selection of the optimal number of MAEs (*k*) in **Fig. 1**. Our analytic workflow is the following. We first derived the 11 MAEs (**Method 2**) via Surreal-GAN^4,5^ (**Method 3**) using specific imaging features from each organ, and evaluated the expression of the identified imaging patterns in test datasets from the same study and other independent studies. We then performed a phenome-wide association study (PWAS, **Method 4**) to establish cross-organ phenotypic landscapes between the 11 MAEs and other clinical traits. A proteome-wide association study (ProWAS, **Method 5**) was conducted to link the 11 MAEs to 2923 plasma proteins (Olink) from UKBB and generate their expression profiles using organ/tissue-specific RNA-seq and protein data^32^. We conducted genome-wide associations (GWAS) to link the 11 MAEs with common SNPs. Subsequently, we performed several post-GWAS analyses to partially validate the genetic signals, such as genetic correlation and Mendelian randomization (**Method 6**). Lastly, we evaluated the clinical relevance of the 11 MAEs in a set of prediction tasks, including mortality, AD progression, incidence of DEs, medication status, and preclinical AD drug (i.e., Solanezumab) outcome (**Method 7**). All the results and code, including the GWAS summary statistics, are publicly accessible through the MEDICINE knowledge portal: https://labs-laboratory.com/medicine/.

**Figure 1:**
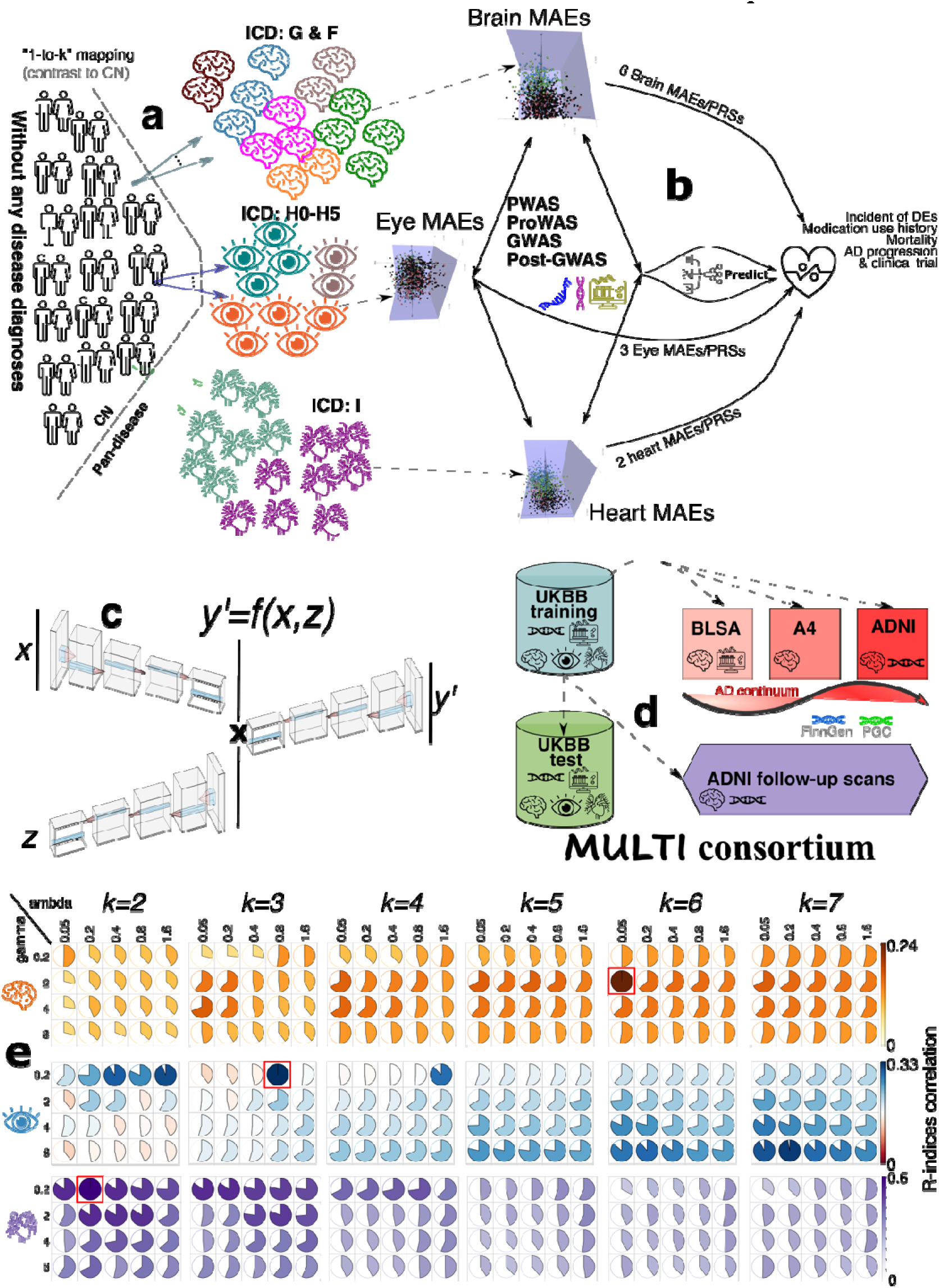
Schematic diagram of the definition of pan-disease, method of analysis, network architecture underlying Surreal-GAN, datasets, and model selection procedure. **a)** Using a weakly-supervised representation deep model called Surreal-GAN, we introduce a novel approach to conceptualizing “pan-disease” in the brain, eye, and heart. In this framework, the reference domain consists of a healthy control (CN) population, and the target domain consists of a pan-disease patient population. Surreal-GAN, separately applied to the brain, eye, and heart pan-disease populations, seeks a “*1-to-k*” mapping to model distinct disease dimensions identified as MAEs. CN participants are defined as those with no diagnosed diseases across multiple sources in the UKBB, while the pan-disease group for each organ includes patients with any organ-specific disease identified using ICD-10 codes. **b**) The 11 MAEs for the brain, eye, and heart served as phenotypes for an extensive array of downstream analyses, including phenotype-wide association studies (PWAS), proteome-wide association studies (ProWAS), genome-wide association studies (GWAS), post-GWAS, and prediction modeling tasks. **c**) The network architecture for the key mapping function of the Surreal-GAN model. It learns a function *f*, which maps the CN data x to synthesized pan-disease data y^f^ = f(x, z), where z is the latent variable indicating the transformation directions. d) Using data from the MULTI consortium, we trained Surreal-GAN on UKBB training data and then applied it to UKBB test data, as well as independent datasets representing the AD continuum. These included BLSA for aging, A4 for preclinical AD, ADNI for MCI and AD stages, longitudinal follow-up brain MRI from ADNI, and GWAS summary data from FinnGen and PGC. **e**) We performed a grid search to select three optimal parameters (*k*, lambda, and gamma). The reproducibility index (R-index correlation) was used to select the three models. The red rectangle for each organ obtained the highest R-index correlation and was chosen for the optimal model in the subsequent analyses.

## Results

### Reproducible imaging patterns of the 11 MAEs of pan-disease in the brain, eye, and heart

By applying multi-organ imaging features (**Method 2**) to the methodologically-enhanced Surreal-GAN model in the UKBB training set (**Fig. 1** and **Supplementary eTable 1**), we derived 11 MAEs of pan-disease: 6 for the brain (Brain 1-6), 3 for the eye (Eye 1-3), and 2 for the heart (Heart 1-2). Refer to **Method 3** and **Fig. 1** for defining the healthy controls (CN: participants without any disease diagnosis from various resources) and pan-disease patients (PT: patients with any ICD-based disease diagnosis from each organ system). We performed a grid search to choose three hyperparameters to define the optimal number of dimensions (*k*) in each organ pan-disease (**Fig. 1e**).

In the case-control (CN-PT) setting, pan-disease of the brain showed brain atrophy in the frontal, temporal, and occipital lobes (**Fig. 2a**). Brain 1 exhibited a pattern of global atrophy, characterized by pronounced volume loss in the temporal lobe and inferior cerebellum (e.g., https://labs-laboratory.com/bridgeport/music/C128_9), accompanied by a concomitant enlarged brain volume in the superior cerebellum. Brain 2 displayed brain atrophy in the parietal lobe and enlarged brain volume in the frontal lobe (e.g., https://labs-laboratory.com/bridgeport/music/C128_84). Brain 3 showed global atrophy over the entire brain (e.g., https://labs-laboratory.com/bridgeport/music/C128_31) and enlarged brain volume in the cerebellum. Brain 4 showed enlarged brain volume, mainly in the frontal and occipital lobes (e.g., https://labs-laboratory.com/bridgeport/music/C128_13), but without affecting the temporal poles and deep subcortical structures. Brain 5 showed enlarged brain volume in the frontal and temporal lobes (e.g., https://labs-laboratory.com/bridgeport/music/C128_10). Brain 6 displayed frontal lobe atrophy and enlarged brain volume in deep subcortical structures (e.g., https://labs-laboratory.com/bridgeport/music/C128_1). Both similarities and differences were observed across the 6 brain MAEs, echoing the underlying modeling considerations of Surreal-GAN (**Method 3b**). For example, Brains 1, 2, and 3 displayed a common feature of atrophy in deep subcortical structures (C128_1). In contrast, Brain 6 exhibited a distinct pattern characterized by increased volume (**Fig. 2a**). These imaging patterns manifested in the UKBB test dataset (**Supplementary eFigure 1a and 2**). Previous studies have primarily highlighted disease heterogeneity in brain disorders within individual conditions, such as AD^33^, late-life depression^34^, and schizophrenia^35,36^, as well as across transdiagnostic phenotypes^13^. These offer data-driven evidence supporting neuroanatomical similarities across these disease entities, as highlighted by our previous studies involving 9 disease-specific dimensions/subtypes^16^.

**Figure 2:**
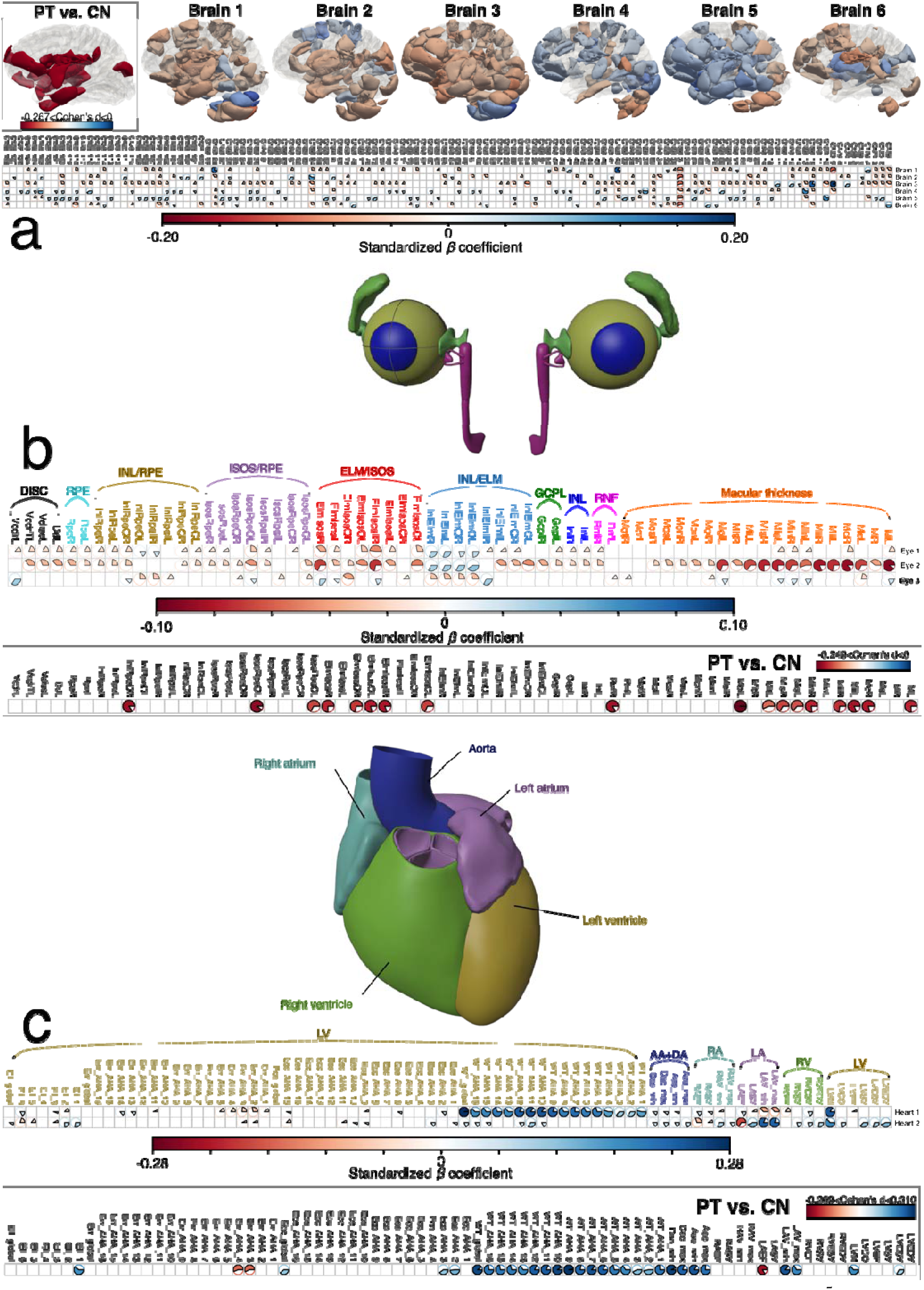
The morphological patterns of the 11 MAEs for pan-disease of the brain, eye, and heart. We employed a weakly-supervised representation learning method (Surreal-GAN^5^) to dissect the morphological heterogeneity of pan-disease of the brain, eye, and heart. Surreal-GAN derives multiple multi-organ AI endophenotypes (MAEs), digitizing multiple distinctive yet co-occurring imaging patterns within the same patient, considering disease severity. We used the standardized beta coefficients from linear regression models as effect sizes to represent significant associations, applying Bonferroni correction based on the total number of imaging features and MAEs. **a**) The imaging patterns of the 6 MAEs of the brain pan-disease. The ICD-10 codes G (Diseases of the nervous system) and F (Mental and behavioral disorders) were used to define the brain pan-disease patient (PT) population. We identified the healthy control (CN) population by excluding all individuals with an ICD diagnosis recorded at any inpatient session. **b)** The imaging patterns of the 3 MAEs of the eye pan-disease. The ICD-10 codes H0-H5 (Diseases of the eye and adnexa) were used to define the eye pan-disease patient population. **c**) The imaging patterns of the 2 MAEs of the heart pan-disease. We used the ICD-10 code I (Diseases of the circulatory system) to define the heart pan-disease patient population. We also showed the imaging patterns of the three pan-diseases in a case-control logistic regression analysis (PT vs. CN). Readers can visualize the brain imaging MRI features used to compute the brain MAEs via the BRIDGEPORT knowledge portal: https://labs-laboratory.com//bridgeport. The eye and heart imaging features are publicly available at https://labs-laboratory.com/medicine/eye and https://labs-laboratory.com/medicine/cardiovascular. The ICD-10 code is documented at: https://biobank.ndph.ox.ac.uk/ukb/field.cgi?id=41270.

The CN-PT analyses showed global retinal morphological thinning for pan-disease of the eye. For the three eye MAEs, Eye 1 showed the most pronounced thinning of the retina between the external limiting membrane (ELM) to the inner and outer photoreceptor segments (ISOS) (ELM/ISOS) (e.g., https://labs-laboratory.com/medicine/elmisos_thickness_of_inner_subfield_right_f28517_0_0). Eye 2 showed a globally thinner retina, especially for overall macular thickness (e.g., https://labs-laboratory.com/medicine/overall_macular_thickness_left_f27800_0_0) and a thicker outer nuclear and outer plexiform layer (between the inner nuclear layer (INL) and the external limiting membrane (ELM)) (e.g., https://labs-laboratory.com/medicine/inlelm_thickness_of_the_outer_subfield_left_f28510_0_0). Eye 3 displayed a greater macular thickness, vertical cup-to-disc ratio (e.g., https://labs-laboratory.com/medicine/vertical_cup_to_disc_ratio_vcdr_left_f27857_0_0), as well as other macular thickness measures (**Fig. 2b**). These imaging patterns manifested in the UKBB test dataset (**Supplementary eFigure 1b**). Ocular disorders, such as age-related macular degeneration and glaucoma, exhibit pronounced phenotypic heterogeneity^37,38^, and our study elucidates the complex morphological alterations underlying these diseases through 3 eye dimensions with distinct imaging patterns.

For pan-disease of the heart, the CN-PT analyses showed globally increased measures regarding cardiac and aortic function, except for the left atrial (LA) ejection fraction (LAEF: https://labs-laboratory.com/medicine/la_ejection_fraction_f24113_2_0) and the left ventricle (LV) radial strain AHA (American Heart Association) 3 (Err_AHA_3) and Err_AHA_3. Heart 1 displayed the most profound increased measures in the LV mean myocardial wall thickness (WT; e.g., https://labs-laboratory.com/medicine/lv_mean_myocardial_wall_thickness_aha_9_f24132_2_0). Heart 2 showed a global increase in cardiac and aortic function but with a decrease in LAEF and RAEF (**Fig. 2c**). These imaging patterns manifested in the UKBB test dataset (**Supplementary eFigure 1c**). Cardiovascular diseases exhibit widespread heterogeneity across conditions and ethnic populations^39^, and we present a comprehensive, data-driven framework elucidating the morphological heterogeneity underlying various heart disease pathophysiologies. **Supplementary eFile 1-3** presents the detailed results for the three organs.

We applied the 6 brain MAEs to three independent brain MRI datasets covering the entire AD continuum, including aging (BLSA^40,41^), preclinical AD (A4^6,42^), and mild cognitive impairment (MCI)/AD (ADNI^43^) cohorts, which were systematically consolidated and statistically harmonized (**Method 1** and **Supplementary eTable 1**). The imaging patterns of these 6 brain MAEs were consistently expressed throughout the AD continuum (**Extended Data Fig. 1**a-c), as well as their longitudinal follow-ups from ADNI^43^ (**Extended Data Fig. 1**d). Brain 1 and Brain 3 showed positive associations with cerebrospinal fluid (CSF) levels of total-tau (t-tau_181p_; Brain 1: P-value=3.90×10^−4^, *r*=0.17; Brain 3: P-value=3.06×10^−5^, *r*=0.17) and phosphorylated-tau181 (p-tau_181p_; Brain 1: P-value=6.80×10^−4^, *r*=0.16; Brain 3: P-value=1.25×10^−4^, *r*=0.15), while Brain 5 was positively linked to CSF levels of Aβ_1-42_in ADNI (**Extended Data Fig. 1**e; P-value=9.11×10^−6^, *r*=0.17). A weak negative correlation was detected between Brain 6 and CSF p-tau_181p_ in ADNI (P-value=5.40×10^−4^, *r*=-0.01), which was then confirmed using plasma p-tau_181p_ in UKBB (P-value=0.01, *r*=-0.06). These findings align with previous studies connecting AD with these CSF neuropathological biomarkers^44^. Additionally, Brain 2 was most strongly associated with fluid intelligence (P-value=6.94×10^−21^, *r*=-0.05) and numerical memory (P-value=1.56×10^−16^, *r*=-0.05) in UKBB, among other significant associations with cognition (**Extended Data Fig. 1**f). **Supplementary eFile 4** and **eTable 2-3** present the detailed results. **Supplementary eNote 1** discusses our strategies and considerations for harmonizing multi-site brain MRI data.

### The phenotypic landscape between the 11 MAEs and 9 multi-organ biological age gaps and 2101 DEs

We then associated the 11 MAEs with 9 multi-organ biological age gaps (BAGs) from our previous study^21^ and 2101 ICD-10-defined DEs from UKBB. We hypothesized that the PWAS (**Method 4**) results demonstrated both within-organ specificity (brain MAEs linked to brain-related phenotypes/traits) and cross-organ interactions (brain MAEs linked to other organ systems).

We observed stronger (standardized *β* coefficient) MAE-BAG associations for within-organ than for cross-organ MAE-BAG pairs. For example, the strongest association was observed between Eye 2 and eye BAG (*β=*0.059±0.0016), higher than the association between Eye 2 and brain BAG (*β=*0.026±0.0053) and between Eye 3 and pulmonary BAG (*β=*0.011±0.0025). When comparing the effect sizes across the three organs, the eye MAEs showed the strongest correlations between organ-specific MAE-BAG pairs, followed by the heart and brain MAEs. Several factors can contribute to this observation. For example, the eye’s structure and function are highly conserved across species^45^, making it more susceptible to age-related changes. Furthermore, the eye has a high metabolic rate, leading to increased oxidative stress and cellular damage^46^. In contrast, the brain’s compensatory mechanisms, such as neuroplasticity and functional redundancy^47^, may help to mitigate age-related declines; the brain comprises diverse cell types, each with unique aging profiles, which might contribute to its relative resilience^48^ (**Fig. 3a** and **Supplementary eTable 4**). **Extended Data Fig. 2**a-c showcases the scatterplot of representative MAE-BAG associations of the three organs.

**Figure 3:**
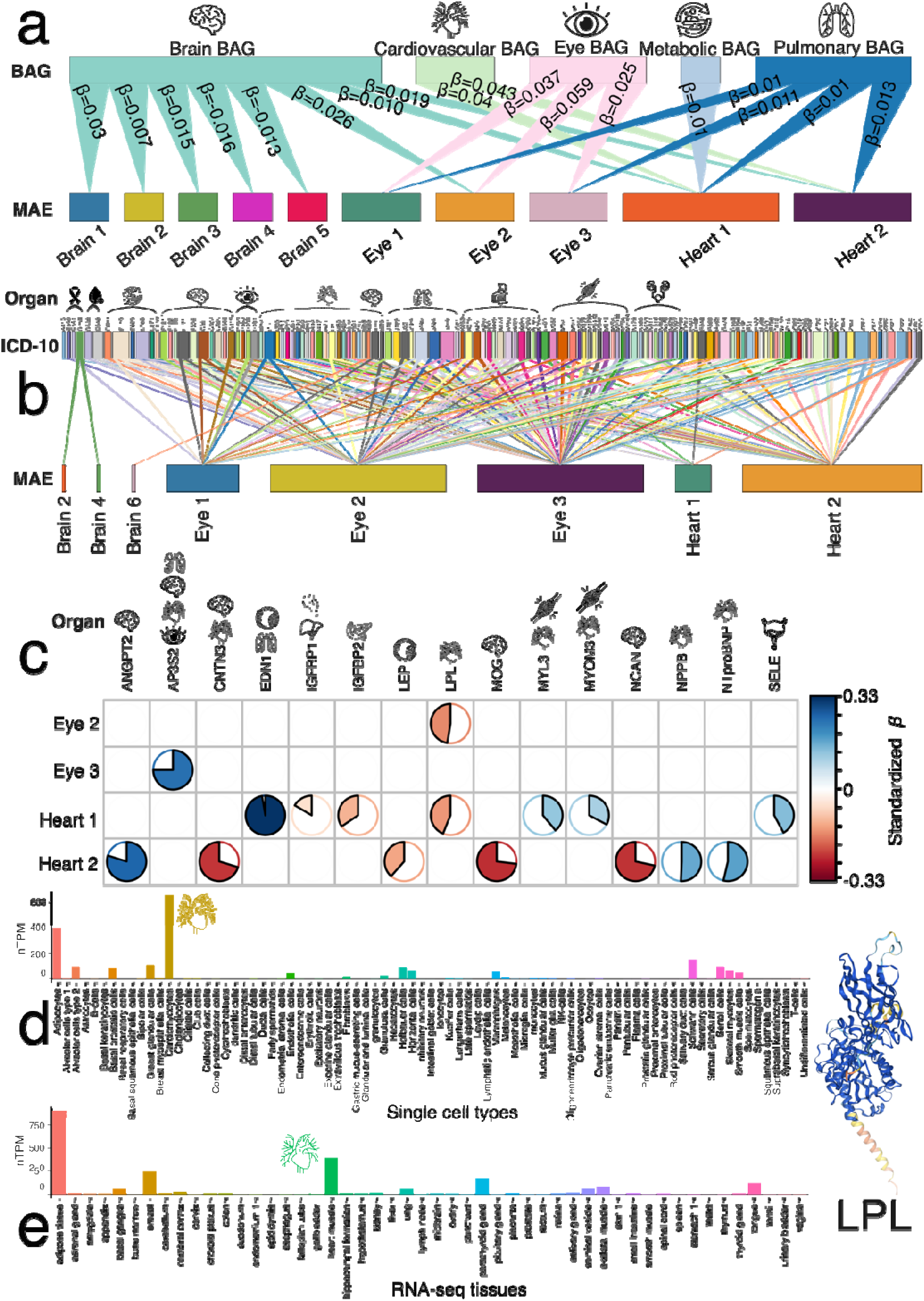
The phenotypic landscape and proteomic profile of the 11 MAEs of the brain, eye, and heart. **a)** Phenotypic associations (PWAS) between the 11 MAEs and 9 AI-derived biological age gaps (BAGs) from our previous study^21^. A linear regression model was constructed to predict the MAEs using the BAGs, adjusting for various covariates (e.g., age and sex) and organ-specific covariates (e.g., blood pressure for heart MAEs and scanner positions for brain MAEs). The *β* values were standardized. Significant associations were determined by applying Bonferroni correction based on the 9 BAGs and the 11 MAEs (P-value<0.05/9/11). **b**) PWAS between the 11 MAEs and disease endpoints (DEs) defined by the ICD-10 code (>20 patients) were evaluated by constructing a logistic regression model to predict the DE of interest using the BAGs and related covariates. Significant associations were determined by applying Bonferroni correction based on the 11 MAEs and the 2101 DEs (P-value<0.05/2101/11). **c**) Proteome-wide association studies (ProWAS) between the 11 MAEs and 2923 plasma proteins via a linear regression model. Using RNA-seq and single-cell data, we also annotated the primary organ systems linked to each significant protein based on their data-type-specific expression levels. Significant associations were determined by applying Bonferroni correction based on the 11 MAEs and the 2923 proteins (P-value<0.05/2923/11). **d**) The single-cell-type expression (i.e., nTPM) for the LPL protein using single-cell transcriptomics data from the Human Protein Atlas (HPA). **e**) The tissue-specific RNA expression overview for the LPL protein using data from HPA and the Genotype-Tissue Expression (GTEx) project. We present the LPL protein structure as predicted by the AlphaFold model (Monomer v2.0^51^).

We then associated the 11 MAEs with ICD-10-based DEs (at least >20 patients) using UKBB data. Among the 11 MAEs and 2101 DEs, we identified 3 significant MAE-DE associations (P-value < 0.05/2101/11) for the brain, 127 for the eye, and 66 for the heart MAEs (**Fig. 3b**). For example, Brain 6 was positively linked to cerebral infarction (ICD-10: I639; *β=*0.40±0.08; P-value=2.16×10^−6^). The eye MAEs were not only associated with DEs of the eye but also those of other organ systems. For example, primary open-angle glaucoma (ICD-10: H401) was positively associated with both Eye 1 (*β=*0.44±0.09; P-value=3.74×10^−7^) and Eye 3 (*β=*0.33±0.07; P-value=6.02×10^−9^). Multiple sclerosis was positively linked to all three eye MAEs (P-value<4.97×10^−9^; *β*>0.35). Heart diseases like hypertension (ICD-10: I10) were also positively linked to all MAEs (P-value<6.02×10^−9^; *β*>0.11). The two heart MAEs were associated with heart-related diseases, including angina pectoris (I209 vs. Heart 2; *β=*0.42±0.04; P-value=9.49×10^−24^), acute subendocardial myocardial infarction (I214 vs. Heart 2; *β=*0.41±0.07; P-value=2.47×10^−8^), and atherosclerotic heart disease (I251 vs. Heart 1; *β=*0.21±0.04; P-value=1.22×10^−7^). Diseases of other organs were also linked to the two heart MAEs, such as cataracts (H269 vs. Heart 2; *β=*0.20±0.03; P-value=4.88×10^−8^), non-insulin-dependent diabetes mellitus (H269 vs. Heart 1 & 2; *β>*0.25; P-value<4.41×10^−10^), and obesity (H669 vs. Heart 2; *β=*0.25±0.04; P-value=1.64×10^−7^). Finally, we also found that different types of cancer were also linked to the MAEs, such as malignant neoplasm of bronchus and lung (C349; Eye 2), malignant neoplasm of corpus uteri (C541; Eye1), and submucous leiomyoma of the uterus (D250; Brain 2 and Brain 4). Increasing evidence pinpoints potential connections between cancer and certain brain and eye diseases, though these relationships are intricate and not yet fully understood. For example, it has been proposed that AD progresses through mechanisms and pathways similar to certain cancers. Intriguingly, the two have a noted negative association^49^ (**Supplementary 2**).

In our secondary PWAS (**Method 4b**), we also linked the 11 MAEs to all other clinical variables not directly related to the three organs, exemplified by the strong association between Heart 2 and “alcohol drinker status” (Field ID=20117) (*β=-*0.01±0.001; P-value=4.00×10^−31^). The potential beneficial effect of alcohol consumption on cardiovascular diseases, as suggested by observational studies, has been a continued debate in the community because Mendelian randomization studies challenge this notion^50^. The detailed results are discussed in **Supplementary eFigure 3, eFile 6,** and **eNote 2**. **Extended Data Fig. 2**d-f showcases the boxplot of representative MAE-DE associations of the three organs.

### The proteomic profile between the 11 MAEs and 2923 plasma proteins

Our ProWAS analyses (**Method 5a**) linked the 11 MAEs to 2,923 plasma proteins using the Olink platform and evaluated the expression profile (with-organ or cross-organ) of significant proteins using organ/tissue-specific (single-cell) RNA-seq data (**Method 5b**).

After correcting for multiple comparisons (P-value<0.05/2923/11), we found 16 significant associations. For example, Heart 1 (*β=-*0.14±0.03; P-value=9.98×10^−7^; *N*=3130) and Eye 2 (*β=-*0.15±0.03; P-value=1.46×10^−6^; *N*=3352) were negatively associated with the LPL protein (**Fig. 3c**). LPL plays a pivotal role in triglyceride metabolism by catalyzing the hydrolysis of triglycerides from circulating chylomicrons and VLDL, thereby facilitating lipid clearance from the bloodstream and regulating lipid utilization and storage. We demonstrated an organ-specific expression pattern for LPL by showing it exhibited the highest expression level (655.2 nTPM) in cardiomyocytes among 81 single-cell types (**Fig. 3d**). At the tissue level, LPL expression was prominently elevated in the heart muscle (394.5 nTPM) compared to other tissues, further supporting its cardiac-enriched expression profile (**Fig. 3e**). Detailed statistics of our ProWAS are presented in **Supplementary eFile 7a**. Another significant signal was observed between Eye 3 and the AP3S2 protein, which showed a high RNA expression in the tissues of the brain, eye, and heart (**Supplementary eFigure 4**). **Extended Data Fig. 2**g-j showcases the scatterplot of representative MAE-protein associations of the eye and heart, the protein-protein interaction network (**Method 5d**) for the LPL protein, and the volcano plot for Heart 2 ProWAS results. Heart 1 and 2 demonstrated more significant plasma protein associations than brain and eye MAEs. Biologically, this may be due to several factors. First, the heart’s proximity to circulation allows direct interaction with many plasma proteins, often produced by or related to cardiovascular function and disease^52,53^. Furthermore, heart-related diseases typically cause systemic changes that can be easily detected in the blood, whereas brain diseases often involve localized molecular changes relatively confined behind the blood-brain barrier^54^.

The results of ProWAS of brain MAE using UKBB’s Olink and the comparison with BLSA SomaScan proteomics data are discussed in **Supplementary eNote 3, eFile 7b,** and **eFigure 12**. Importantly, we observed that SomaScan proteins exhibited stronger associations with the 6 brain MAEs than Olink proteins, despite BLSA SomaScan having a much smaller sample size (*N*=924). The *β* coefficients of the 44 common significant MAE-protein pairs showed moderate correlation (Pearson’s *r*=0.26), which agrees with a previous investigation^55^. This observation may be attributed to SomaScan’s greater sensitivity and broader protein coverage, which likely enhances its ability to detect significant associations.

### The genetic architecture of the 11 MAEs

We first conducted GWASs to identify common genomic loci and regions shared among the 11 MAEs across the three organs. Next, we estimated three key genetic parameters: *i*) SNP-based heritability (h^2^), *ii*) natural selection signatures (*S*), and *iii*) polygenicity (*Pi*). Finally, we performed gene-set enrichment analyses to determine whether MAE-associated genes were enriched in any drug categories, providing insights for potential drug repurposing and cell- and tissue-specific partitioned heritability estimates (**Method 6**).

For the 11 primary GWASs (**Method 6a**) using European ancestry populations, we identified 42 (P-value<5×10^−8^/11), 56, and 7 genomic locus-MAE associations for the 6 brain, 3 heart, and 2 heart MAEs, respectively. We denoted the genomic loci using their top lead SNPs (**Supplementary eMethod 1**) defined by FUMA fully considering linkage disequilibrium (LD); the genomic loci are presented in **Supplementary eFile 8**. To visually present the shared genomic region among the 11 MAEs, we showed that only 5, 1, and 1 common cytogenetic regions (based on the GRCh37 cytoband) were jointly linked to the brain, eye, and heart MAE pairs, respectively (**Fig. 4a**). In general, the 11 MAEs showed limited genetic overlap based on their physical locations in the genome, which was subsequently supported by the weak pair-wise genetic and phenotypic correlations observed (**Fig. 5a**). We performed several quality checks to scrutinize the robustness of the primary GWAS and a detailed discussion is presented in **Supplementary eNote 4**. We also performed a phenome-wide association look-up analysis (PheWAS) using the GWAS Atlas^56^ platform (**Method 6b**) to link the top lead SNPs of each locus with previous literature (**Supplementary eNote 5 and eFigure 5**). Manhattan and QQ plots of the 11 primary GWASs are presented in our MEDICINE portal (e.g., eye MAEs: https://labs-laboratory.com/medicine/eye) and **Supplementary eFigure 6**. **Extended Data Fig. 3** presents the trumpet plots of the effective allele frequency vs. the *β* coefficient of the 11 primary MAE GWASs.

**Figure 4:**
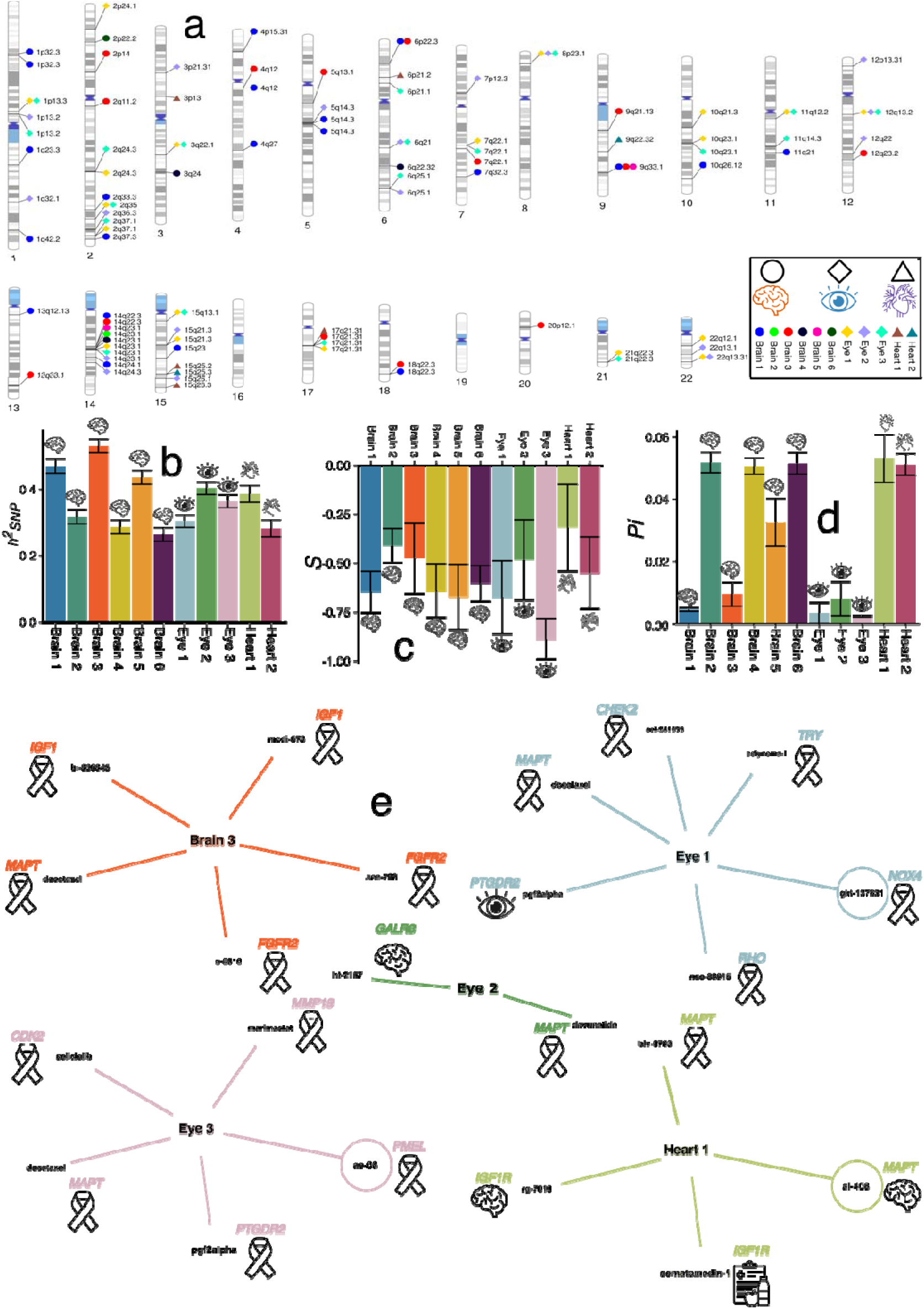
The genetic architecture of the 11 MAEs of the brain, eye, and heart. **a)** Cytogenetic regions where the genomic region was jointly linked to the three organ-specific MAEs. Bonferroni correction was applied to denote significant genomic loci associated with the brain PSCs (P-value<5×10^−8^/11). **b**) The SNP-based heritability () was estimated using the GCTA software for the 11 MAEs. **c**) The selection signature (*S*) was estimated using the SBayesS software for the 11 MAEs. **d**) The polygenicity (*Pi*) was estimated using the SBayesS software for the 11 MAEs. The mean and the standard error of each inferred statistic are shown. **e**) A gene-drug-disease network quantifies an enrichment of the MAE-related gene sets in the target of clinical indication categories and captures potentially repositionable drugs targeting the gene set using the DrugBank database.

**Figure 5:**
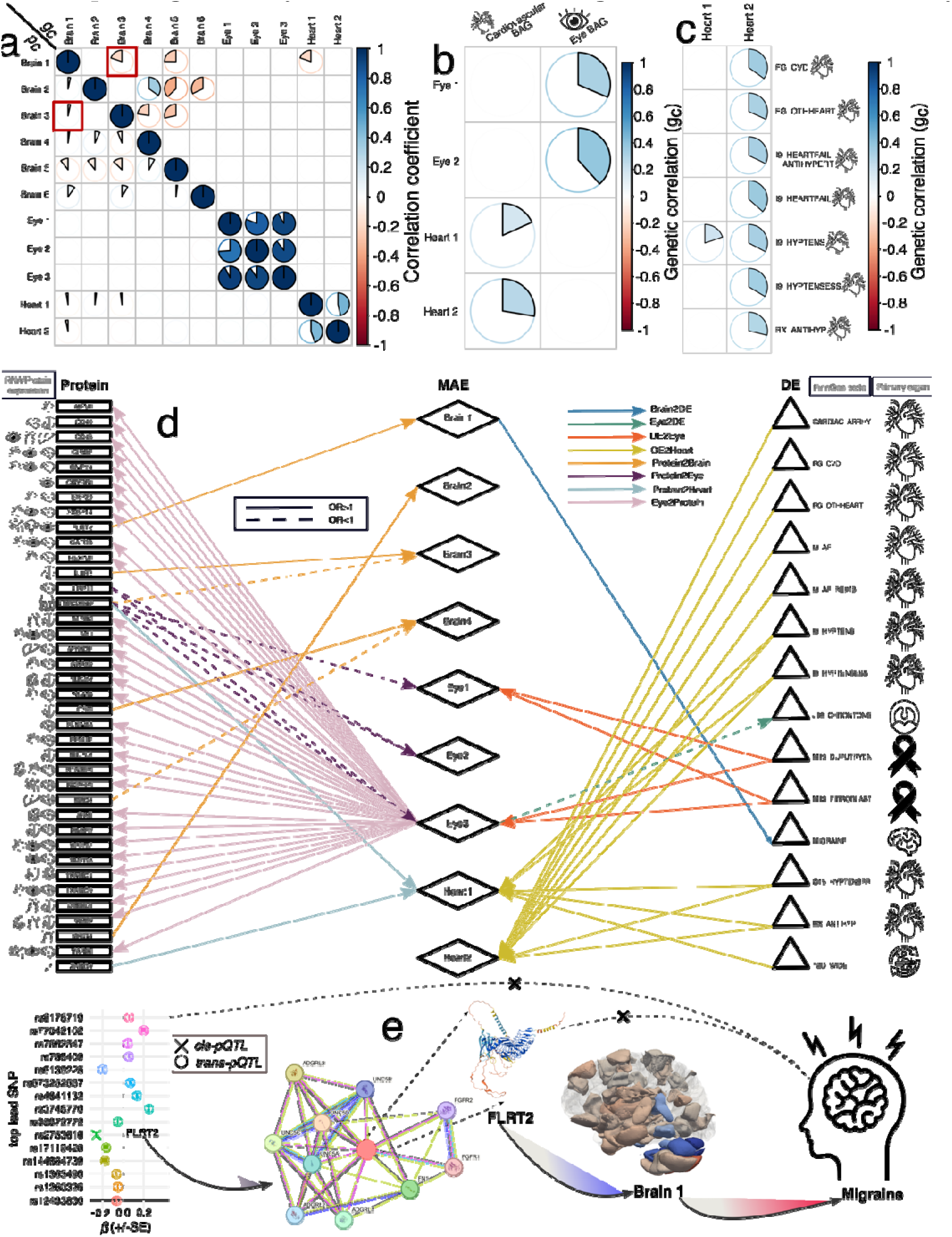
Deciphering multi-layer causal networks from genetic correlation to causality. **a**) Phenotypic association (*p_c_*) and genetic association (*g_c_*) between each pair of the 11 MAEs. Genetic correlations largely mirror phenotypic correlations^69^ of the MAE pairs except for the pair between Brain 1 and Brain 3. **b**) Genetic correlation between the 11 MAEs and the 9 BAGs from our previous multi-organ BAG study^21^. **c**) Genetic correlation between the 11 MAEs and the 525 disease endpoints (DEs) using GWAS summary statistics from FinnGen and PGC. For **a c**, we employed the Bonferroni correction to account for multiple comparisons using stringent P-value thresholds adjusted based on the total number of MAEs and phenotypes or DEs examined. **d**) We constructed a three-layer protein-MAE-DE causal network by employing bi-directional two-sample Mendelian randomization, following a rigorous quality control procedure to select exposure and instrumental variables (number of IVs>8), correct for multiple comparisons (based on either the number of exposure or outcome variables), and perform sensitivity analyses (e.g., horizontal pleiotropy and removing overlap populations) to scrutinize the robustness of our results. The *Heart2DE* and *Heart2Protein* networks were not included due to insufficient IVs after a quality check (<8). The *DE2Brain* and *Heart2Protein* networks are not shown because the results did not survive the correction for multiple comparisons. **e**) A representative causal pathway example demonstrates how pQTLs (both *cis* and *trans*) influence the FLRT2 protein, affecting Brain 1 and increasing the risk of migraine disorder. We present the FLRT2 protein’s structure as predicted by the AlphaFold model (Monomer v2.0^51^), along with a protein-protein interaction network. This network emphasizes its interaction with UNC5D and other members of the UNC-5 family, which are all crucial for processes such as cell-cell adhesion, axon guidance, and cell migration at the cellular level, as well as the development of cortical neurons and cardiac morphogenesis at the pathway level.

We estimated the h^2^ to quantify the proportion of phenotypic variance attributable to common genetic variants across the genome and organs (**Method 6c**). The GCTA^57^ software revealed significant SNP-based heritability in all 6 brain MAEs (0.26<h^2^ <0.53; P-value<1×10^−20^) using individual-level genotype data, followed by the 3 eye MAEs (0.30<h^2^ <0.40; P-value<1×10^−20^), and the 2 heart MAEs (0.28<h^2^ <0.39; P-value<1×10^−20^) (**Fig. 4b**). The results generated by LDSC^58^ (h^2^ =0.20±0.05) and SbayesS^59^ (h^2^ =0.19±0.05), which use GWAS summary data and a reference panel for LD information, showed a lower estimate than GCTA (h^2^ =0.37±0.09; t-value>5.49; P-value<2.24×10^−5^); the three sets of estimates were significantly correlated with each other (P-value<1.42×10^−6^; *r*>0.97). Detailed statistics are presented in **Supplementary eTable 5a-c**.

Next, we explored the evolutionary processes of pan-diseases across the three organs, particularly how these traits evolve and adapt through natural (e.g., negative/purifying) selection and genetic variation; we applied the SBayesS^59^ method to calculate the selection signature (*S*) and polygenicity (*Pi*) for the 11 MAEs (**Method 6d**). Eye 3 showed the most prominent negative selection signature (*S=-*0.88±0.10) (**Fig. 4c**), echoing its lowest polygenicity estimate (*Pi=*0.002±0.0004). Eye 1-3 showed the lowest polygenicity estimates (**Fig. 4d** and **Supplementary eTable 6**). The observed cross-organ trends in negative signature and polygenicity estimates can be attributed to multiple factors. First, the evolutionary conservation of the retina’s structure and function across species^45^ suggests it is subject to strong evolutionary pressure to maintain stability. Even minor genetic changes affecting retinal function may have detrimental effects, resulting in stronger purifying (negative) selection against harmful variants. In contrast, organs such as the brain and heart may display greater evolutionary flexibility or redundancy (i.e., more complicated compensatory pathways) in their genetic architecture, allowing for a higher tolerance of mutations. In addition, brain disorders are often more polygenic than eye and heart diseases due to the brain’s complexity and the large number of genes involved in its development, function, and maintenance^60,61^.

We performed a gene-drug-disease enrichment analysis (**Method 6e**) using positionally and functionally mapped genes linked to the 11 MAEs within drug-target gene sets from the DrugBank database^62^. This analysis generated a gene-drug-disease network to identify drugs with repurposing potential, a strategy proven to enhance the success rate of drug development according to existing literature^63,64^. For the Brain 3-mapped genes (i.e., *MAPT*, *FGFR2*, and *IGF1*), we found 5 significant interactions with 5 drugs for treating neoplasms of uncertain or unknown behavior. For example, Docetaxel is a chemotherapy drug used to treat various types of cancer, including lung cancer (e.g., NCT code: NCT04303780) and breast cancer, often combined with other medications. The mapped *MAPT* gene is a targeted gene for several drugs, including neurodegenerative conditions like AD. Another example is BIIB080, also known as IONIS-MAPTRx (NCT code: NCT03186989), an investigational drug targeting the microtubule-associated protein tau (MAPT) gene to treat AD^65^. We also found abundant cancer-related (e.g., Docetaxel) interactions for Eye 1 and Eye 3. For instance, PGF2alpha (Prostaglandin F2alpha; mapped gene: *PTGDR2*) is a naturally occurring prostaglandin with several pharmaceutical uses, primarily related to its role in reproductive health, as well as for treating glaucoma (e.g., latanoprost^66^). NSC8895 (mapped gene: *RHO*) is a small-molecule drug investigated for its potential as an anti-cancer agent or for other therapeutic uses. Finally, Heart 1-mapped genes were linked to drugs/molecules treating several organ-specific diseases. For example, RG-7010 was developed by Roche to initially treat several motor neuron diseases, including amyotrophic lateral sclerosis (**Fig. 4e**). Detailed statistics are presented in **Supplementary eFile 9**.

We further provided evidence for the significant enrichment of cancer-related drugs in our gene-drug-disease network using RNA-seq data in 17 cancer types from the TCGA database. This was exemplified by the MAPT protein linked to Brain 2, Eye 1, Eye2, and Heart 2 (**Fig. 4e**). Enhanced cancer specificity was observed in breast cancer (6.7 FPKM) and glioma (12.2 FPKM) (**Supplementary eFigure 7a**). Correspondingly, the MAPT protein showed high single-cell type-specific expression profiles in the brain (**Supplementary eFigure 7b**) and breast tissues (**Supplementary eFigure 7c**). Research has shown that breast cancer tissues and glioma exhibit increased levels of tau protein, which may contribute to tumor progression by promoting key processes, including enhanced cell proliferation and migration, increased invasiveness, modulation of signaling pathways, and facilitation of metastasis^67^. Finally, we also performed cell- and tissue-specific partitioned heritability estimates^68^ using data from three cell types, multi-tissue gene expression, and chromatin interactions (**Method 6f**) to support further the organ-specific patterns of our GWAS signals (**Supplementary eFile 10**). Significant heritability enrichment (P-value<0.05/697/11) was found for Brain 5 in ganglion eminence-derived primary cultured neurospheres (P-value=3.39×10^−6^) and fetal brain tissues (P-value=1.79×10^−10^) in H3K4me.

### The genetic correlation between the 11 MAEs and other clinical traits

We estimated the genetic correlation between the 11 MAEs for three biomarker sets: *i*) between the 11 MAEs, *ii*) with the 9 BAGs, and *iii*) with 525 DEs (**Method 6g**).

We estimated the pair-wise phenotypic correlation (*p_c_*via Pearson’s *r*) and genetic correlation (*g_c_*) for the 11 MAEs. First, our results revealed a pronounced within-organ correlation structure. In contrast, cross-organ correlations were much weaker, reflecting the scarcity of the shared genomic regions among the three organs (**Fig. 4a)**. An exception emerged between Brain 1 and Heart 1, which displayed a positive genetic correlation (*g_c_*=0.20±0.06; P-value=8.0×10^−4^) (**Fig. 5a**). Furthermore, our analysis revealed a strong concordance between the genetic correlations and phenotypic correlations across the 11 MAEs, providing empirical support for the long-standing Cheverud Conjecture^69^. One exception was observed between Brain 1 and Brain 3, where a negative *g_c_* and a weak positive *p_c_* were obtained. This phenomenon is likely influenced by multiple factors, including gene-environment interactions^70^, epistasis^71^, pleiotropy, LD, and genetic heterogeneity. Detailed statistics are presented in **Supplementary eTable 7**.

We then observed strong within-organ associations between the MAEs and their respective organ BAGs (**Fig. 5b**) and DEs (**Fig. 5c**). For example, we found that Eye 1 (*g_c_*=0.31±0.08; P-value=1.0×10^−4^<0.05/11/9) and Eye 2 (*g_c_*=0.38±0.06; P-value=1.0×10^−10^) were positively associated with eye BAG, and Heart 1(*g_c_*=0.18±0.05; P-value=5.0×10^−4^) and Heart 2 (*g_c_*=0.28±0.07; P-value=8.0×10^−5^) with cardiovascular BAG. Detailed statistics are presented in **Supplementary eTable 8**. Among 525 unique DEs from FinnGen and PGC, we found 8 significant MAE-DE associations, highlighting prominent genetic associations between Heart 2 and various heart and vascular diseases, such as cardiovascular disease (FinnGen code: FG_CVD; *g_c_*=0.34±0.05; P-value<1.0×10^−10^), hypertension (I9_HYPTENS; *g_c_*=0.33±0.06; P-value<1.0×10^−10^), and heart failure (I9_ I9_HEARTFAIL_EXMORE; *g_c_*=0.37±0.08; P-value<1.0×10^−10^). Detailed statistics are presented in **Supplementary eFile 11**.

### Multi-layer causal networks support the endophenotype hypothesis: MAE functions as a causal intermediate phenotype

We further deciphered the genetic association by building a detailed causal network, spanning from underlying genetics to proteomics, MAEs, and DEs. **Supplementary eFigure 8** illustrates a causal pathway inspired by the endophenotype hypothesis^7^. This was accomplished using two-sample, bi-directional Mendelian randomization^72^, which combined large-scale GWAS summary data from UKBB, FinnGen, and PGC. We ensured the robustness of our results by performing systematic quality checks and sensitivity analyses (**Method 6h**).

We performed Mendelian randomization analyses among the 2923 plasma proteins, 11 MAEs, and 525 DEs, resulting in 12 directional causal networks (e.g., *Brain2DE* and *Protein2Eye*) depending on the exposure and outcome variables. After a quality check procedure for instrumental variable (IV) selection (>8 IVs) and Bonferroni correction, we identified in total 44 significant protein-MAE associations (P-value<0.05/number of proteins) and 24 MAE-DE associations (P-value < 0.05/number of DEs) (**Fig. 5d**). For the *Protein2Brain* network, we identified 6 significant causal relationships from 6 proteins to 4 brain MAEs. For example, a potential causal relationship was established from the LRRC37A2 protein to Brain 3 [P-value=7.64×10^−8^; OR (95% CI)=0.95 (0.93, 0.96); number of IVs=9]. Within the *Protein2Eye* network, we found 5 significant signals from 2 proteins to 3 eye MAEs. For instance, the LRP11 protein showed a potential causal effect on Eye 2 [P-value=1.57×10^−13^; OR (95% CI)=0.93 (0.91, 0.96); number of IVs=13]. For the *Protein2Heart* network, 2 potential causal relationships were found from 2 proteins to Heart 1. For example, LRRC37A2 was positively linked to Heart 1 [P-value=5.03×10^−10^; OR (95% CI)=1.07 (1.04, 1.09); number of IVs=9]. For the *Eye2Protein* network, we found 31 causal relationships from Eye 3 to 30 proteins. For example, Eye 3 was causally linked to the TXNDC9 protein [P-value=1.61×10^−6^; OR (95% CI)=1.17 (1.10, 1.24); number of IVs=21], with prominent expression levels in the brain, eye, and heart.

For the *Brain2DE* network, we found that Brain 1 was causally linked to migraine disorder [FinnGen code: MIGRAINE_TRIPTAN; P-value=8.77×10^−5^; OR (95% CI)=1.16 (1.08, 1.26); number of IVs=24]. For the *Eye2DE* network, Eye 3 was negatively linked to chronic diseases of tonsils and adenoids [J10_CHRONTONSADEN; P-value=2.0×10^−5^; OR (95% CI)=0.87 (0.81, 0.93); number of IVs=20]. We did not find any significant signals for the *Heart2DE* and *Heart2Protein* due to the limited power of the heart MAE GWASs (<8 IVs), and neither did the *DE2Brain* network, which did not survive the multiple comparisons. For the *DE2Eye* network, we found 5 causal links from 2 DEs to Eye 1 and Eye 3. For instance, fibroma was positively linked to Eye 1 [M13_FIBROBLASTIC; P-value=2.23×10^−6^; OR (95% CI)=1.07 (1.04, 1.10); number of IVs=18]. For the *DE2Heart* network, we found 19 causal links from 10 DEs to Heart 1 and Heart 2. For example, type-2 diabetes was positively linked to Heart 1 [T2D_WIDE; P-value=1.225×10^−3^; OR (95% CI)=1.08 (1.03, 1.12); number of IVs=62] (**Fig. 5d**). Detailed statistics are presented in **Supplementary eFile 12**. The results for sensitivity analyses are presented in **Supplementary eFolder 1**.

We constructed a multi-layer causal network and showcased an example to support the endophenotype hypothesis, especially through the mediational model. Our causal analysis revealed a causal path whereby the FLRT2 protein exerted a causal influence on Brain 1 [P-value=3.83×10^−5^; OR (95% CI)= 1.07 (1.04, 1.11); number of IVs=16], which then subsequently increased the risk of migraine disorder [MIGRAINE_TRIPTAN; P-value=8.76×10^−5^; OR (95% CI)= 1.17 (1.08, 1.25); number of IVs=24], suggesting a sequential causal relationship between FLRT2, AI-derived brain structure, and migraine susceptibility. To further confirm the vertical pleiotropy (i.e., mediational model) in this pathway, we first defined pQTLs of the FLRT2 protein and found that these pQTLs were not directly associated with migraine disorder. Additionally, we performed an additional causal analysis using FLRT2 as the exposure and migraine as the outcome, excluding a direct causal link between the two (**Fig. 5e**). **Extended Data Fig. 4** shows sensitivity checks for the two causal signals, confirming that the results are robust to potential violations of Mendelian randomization assumptions. **Supplementary eNote 6** details the sensitivity check analyses. Additionally, we compared the ProGWAS findings of the FLRT2 protein with the *cis*- and *trans*-pQTLs identified by Sun et al.^73^, demonstrating a strong agreement despite differences in sample sizes, the controlled covariates, and the GWAS models employed. A detailed discussion is presented in **Supplementary eNote 7** and **eFigure 9**. To further elucidate the biological relevance of this pQTL, we conducted SNP-to-gene mapping using eQTL data from GTEx v8, focusing on brain gene expression. This analysis identified 40 eQTL-linked SNPs associated with the *FLRT2* gene, with the strongest association showing a P-value of 2.68×10, particularly in the putamen, basal ganglia, and spinal cord regions. SNPs or genes that are both an eQTL and pQTL are likely to have a critical role in gene regulation and protein expression, making them a valuable target for understanding disease mechanisms and developing therapies.

We also validated this causal pathway’s potential cellular and molecular mechanisms using single-cell RNA-seq and proteomics data. We first performed a protein-protein interaction (PPI) network analysis (**Method 5d**). Our PPI analysis identified FLRT2 as associated with other 10 proteins, with a PPI enrichment P-value of 8.62×10^−8^, providing strong evidence that the proteins are partially biologically connected as a group. The associated proteins, including ADGRL3 and UNC-5 family members, are functionally linked to FLRT2. Biologically, FLRT2 plays a crucial role in cell-cell adhesion, migration, and axon guidance. It facilitates cell adhesion through interactions with ADGRL3 and potentially other latrophilins on adjacent cell surfaces. FLRT2 is involved in migrating cortical neurons during brain development by interacting with UNC5D, and mediates axon growth cone collapse, acting as a repellent in neuron guidance, likely in conjunction with other UNC-5 family members. Further functional enrichment analyses reinforced the involvement of relevant biological pathways, including biological processes like anterior/posterior axon guidance (GO-term: GO:0033564; FDR-corrected P-value=4.86×10^−5^), molecular function like netrin receptor activity (GO:0005042; FDR-corrected P-value=5.43×10^−8^), and cellular components like neuron projection (GO:0043005; FDR-corrected P-value=0.017).

Previous studies on migraine patients have demonstrated atrophy in cortical brain areas^74^ involved in pain processing, likely as a result of chronic stimulation in those areas. To support these morphological changes, we further observed that the cerebral cortex exhibited the highest RNA expression score compared to other brain regions in the human brain (nTPM=5.6), pig brain (nTPM=36.7), and mouse brain (nTPM=26.0) (**Supplementary eFigure 10a**). Additional animal experiments by Kaag Rasmussen et al.^75^ using rodent models demonstrated that cortical spreading depression can trigger changes in the cerebrospinal fluid proteome, leading to an increased expression of proteins capable of activating the trigeminal nerve, thus ultimately triggering headache. Single-cell tissue enrichment analyses in the brain showed higher expression value in neuronal cells (e.g., nTPM=902.4 for excitatory neurons c-9) than in glial cells (**Supplementary eFigure 10b**) (**Method 5b**). Additionally, migraines are linked to an increased risk of cardiovascular disease (CVD), including stroke, heart attack, and other serious health problems^76^. FLRT2 participates in fibroblast growth factor signaling pathways at the cellular and biological pathway levels. It is essential for proper cardiac basement membrane organization and epicardium and heart morphogenesis during embryonic development. Similarly, we found single-cell tissue enrichment in the vascular tissue, highlighting the prominent enrichment of mesenchymal cells (e.g., fibroblasts c-7 for nTPM=114.5), as well as endothelial cells (e.g., endothelial cells c-19=129.4 nTOM) (**Supplementary eFigure 10c**). Fibroblasts are directly involved in heart tissue maintenance and repair and may contribute indirectly to the inflammation and vascular changes associated with migraines. Endothelial cells are specialized cells that form the inner lining of blood vessels and lymphatic vessels, potentially supporting the endothelial hypothesis in migraine^77^.

### The clinical promise of the 11 MAEs for precision medicine

We demonstrate the clinical promise of the 11 MAEs in predicting various clinical outcomes through survival analysis and logistic regression: *i*) the incident of DEs, *ii*) AD progression (CN→MCI→AD), *iii*) the risk of mortality, and *iv*) medication use status (**Method 7a-d**).

Utilizing ICD-10-based DEs from UKBB, we demonstrated that MAEs and their corresponding polygenic risk scores (PRSs; **Method 6i**) were significantly linked to the incident of chronic diseases, both specific to their respective organs and across different organ systems (**Method 7a**). For the brain MAE analyses (306 DEs tested), Brain 2 [HR (95% CI)=1.09 (1.04, 1.13); P-value=3.37×10^−5^], Brain 6 [HR (95% CI)=1.08 (1.04, 1.12); P-value=8.97×10^−5^], and Brain 6-PRS [HR (95% CI)=1.05 (1.01, 1.10); P-value=8.44×10^−3^] were significantly associated with primary hypertension (ICD-10 code: I10) (**Fig. 6a**). The 3 eye MAEs demonstrated limited predictive power for the onset of the 415 DEs analyzed. Notably, the only eye-related disease examined was glaucoma (ICD-10 code: H409), but the analysis had extremely imbalanced data, with only 12 cases compared to 674 non-cases. Finally, Heart 1 and Heart 2 were significantly linked to the incident of hypertension (I10; **Fig. 6b**), Hypercholesterolaemia (E780; **Fig. 6c**), and psychoactive substance abuse (Z864; **Fig. 6d**). Detailed statistics, including HR, P-value, and sample sizes, are presented in **Supplementary eFile 13**.

**Figure 6:**
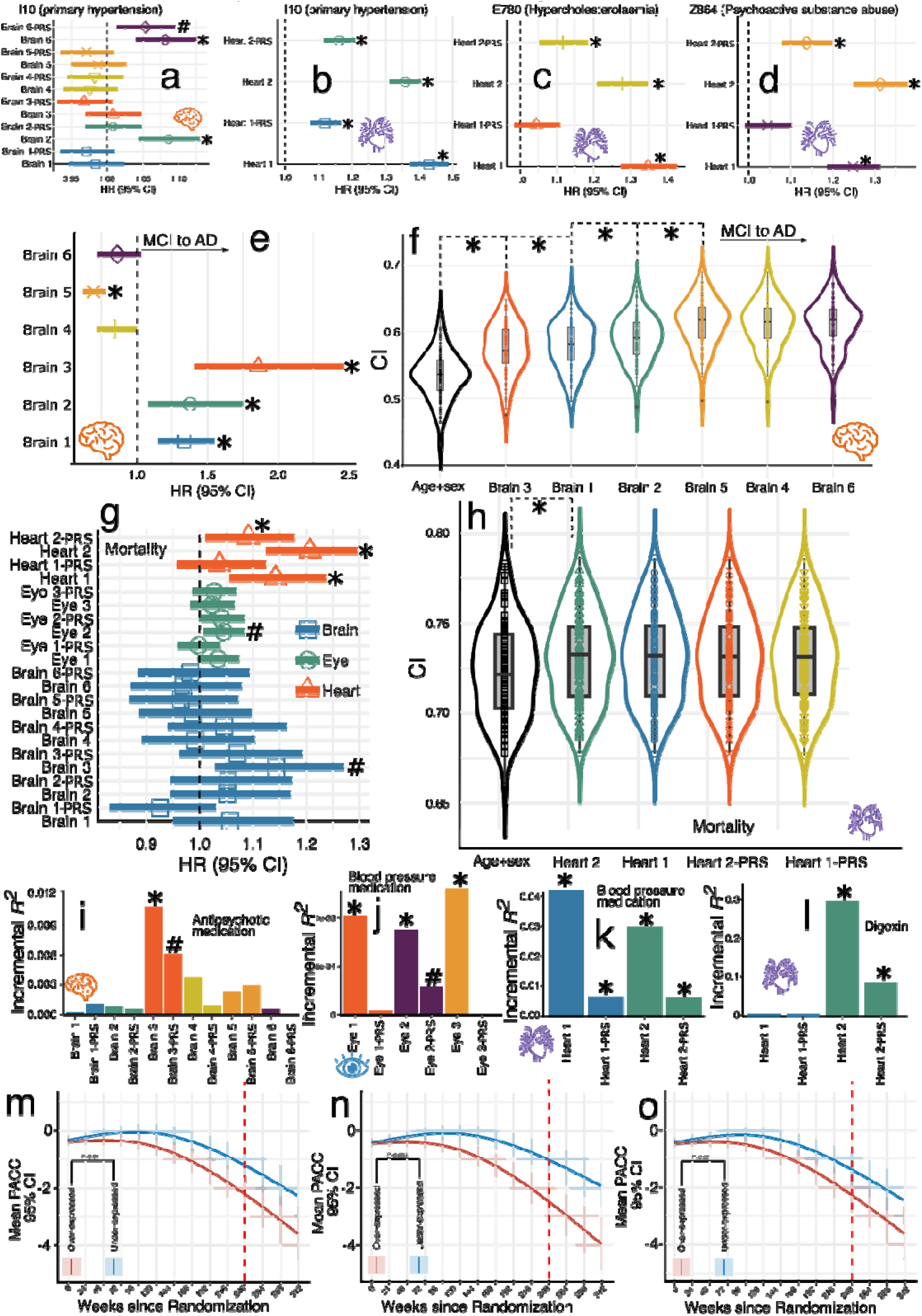
Prediction of and association with disease endpoints, AD progression, mortality risk, medication use history, and preclinical AD drug response. **a**) Brain 2 and Brain 6 exhibit significant associations with an elevated risk of hypertension, as indicated by the corresponding hazard ratios (HRs). Cox proportional hazard models were used to test associations and adjusted for age and sex. **b**) Heart 1 and 2 and their respective PRSs exhibit significant associations with an increased risk of hypertension. **c**) Heart 1, Heart 2, and Heart 2-PRS exhibit significant associations with an increased risk of hypercholesterolemia. **d**) Heart 1, Heart 2, and Heart 2-PRS exhibit significant associations with an increased risk of using psychoactive substances abuse. **e**) Brains 1, 2, and 3 are significantly linked to an increased risk of progressing from mild cognitive impairment (MCI) to AD, while Brain 5 is associated with a reduced risk. **f**) Based on the significance of the brain MAEs demonstrated in **e**, we progressively incorporated additional brain MAEs as features when fitting the Cox proportional hazard model. For each combination of features, 100 iterations of 20% holdout cross-validation were performed to derive concordance indices. **g**) Heart 1, Heart 2, and Heart 2-PRS are significantly linked to an increased mortality risk. **h**) Based on the significance of the brain MAEs demonstrated in **g**, we progressively incorporated additional heart MAEs as features when fitting the Cox proportional hazard model. **i**) Brain 3 and its PRS offer additional predictive power for antipsychotic medication status on top of age and sex. **j**) Eye MAEs offer additional predictive power for blood pressure (BP) medication status on top of age and sex. **k**) Heart MAEs offer additional predictive power for BP medication status on top of age and sex. **l**) Heart MAEs offer additional predictive power for the medication status of Digoxin on top of age and sex. The symbol * indicates statistical significance after the Bonferroni correction, and the symbol # shows the signals that passed the nominal significance threshold (P-value<0.05) but did not survive the correction. CI: concordance index. **m**) The heterogeneity of the cognitive decline trajectories was assessed on the primary outcome, the Preclinical Alzheimer Cognitive Composite (PACC), by comparing two groups based on the expression levels of Brain 1 within the drug group. Participants were stratified into low and high-expression groups by the median Brain 1 value (i.e., under-expressed vs. over-expressed Brain 1 patients). The red dashed line indicates the timeline for evaluating the drug’s effects at week 240. **n**) The heterogeneity of the cognitive decline trajectories on Brain 2. **o**) The heterogeneity of the cognitive decline trajectories on Brain 3.

Using the ADNI data, we demonstrated the prediction power of the 6 brain MAEs for AD progression: CN→MCI→AD (**Supplementary eTable 9** and **Method 7b**). We found that Brain 1 [HR (95% CI)=1.33 (1.15, 1.54); P-value=1.40×10^−4^], Brain 2 [HR (95% CI)=1.38 (1.08, 1.75); P-value=9.35×10^−3^], and Brain 3 [HR (95% CI)=1.86 (1.41, 2.45); P-value=1.00×10^−5^] were positively associated with the risk of MCI → AD progression, whereas Brain 5 showed a negative association [HR (95% CI)=0.69 (0.62, 0.78); P-value=8.48×10^−10^] (**Fig. 6e**). We then cumulatively added the most significant predictors on top of age and sex to evaluate their incremental power for MCI→AD progression, resulting in the highest concordance index (CI=0.61±0.03 in a 100 repeated hold-out cross-validation) when combining age, sex, Brain 3, Brain 1, Brain 2, and Brain 5 (**Fig. 6f**).

We also used the 11 MAEs and their PRSs to predict mortality risk using UKBB data (**Method 7c**). Our analysis revealed that Heart 1 [HR (95% CI)=1.14 (1.06, 1.23); P-value=9.04×10^−4^], Heart 2 [HR (95% CI)=1.20 (1.12, 1.30); P-value=2.01×10^−7^], and Heart 2-PRS [HR (95% CI)=1.09 (1.01, 1.17); P-value=2.53×10^−2^] exhibited a significant association with mortality risk; Brain 3 and Eye 2 showed a nominally significant relationship (P-value < 0.05) (**Fig. 6g**). Given the considerable associations from the heart MAEs, we performed a cumulative prediction analysis. However, only Heart 2 added additional power on top of age and sex, achieving an average CI of 0.73±0.032 (**Fig. 6h**). Detailed statistics, including HR, P-value, and sample sizes, are presented in **Supplementary eTable 10**.

We predicted the medication status for the four drug categories (i.e., antipsychotic medications, blood pressure medications, cholesterol-lowering medication, and insulin) using the 11 MAEs and their PRSs from UKBB data, quantified by the incremental *R*^2^ for additional variance explained by the respective MAE/PRS for the binary medication status (**Method 7d**). We showed that Brain 3 was most predictive of antipsychotic medications (incremental *R*^2^=1.08%; P-value=4.87×10^−3^; *N*=1243) (**Fig. 6i**). Eye 1, Eye 2, and Eye 3 were significant predictors of blood pressure medication status (**Fig. 6j**). However, their contribution to the predictive model was relatively small, as indicated by a smaller incremental *R*^2^ value (incremental *R*^2^<0.13%; P-value<2.2×10^−7^; *N*=19,925), compared to the more substantial impact of the heart MAEs (**Fig. 6k**). Detailed statistics, including P-value, and sample sizes, are presented in **Supplementary eFile 14**. For the 164 individual-level medication predictions, we found that the most significantly associated drug for the brain MAEs was achieved between Brain 6 and amlodipine (incremental *R*^2^=0.311%; P-value<=9.03×10^−9^; *N*=8763), used for blood pressure and cardiovascular disease treatment. The most significant drug association for the heart MAEs was identified with Heart 2 (incremental *R*^2^=30%; P-value<=2.05×10^−19^; *N*=4811), along with the Heart 2-PRS, but not Heart 1, and Digoxin, a drug primarily used to treat heart failure (**Fig. 6l** and **Supplementary eFigure 11**). Finally, the most significant drug for MAEs of the eye was obtained between Eye 2 and tramadol to treat chronic pain. Detailed statistics, including P-value and sample sizes, are presented in **Supplementary eFile 15**.

Given the elevated risks of progression from MCI to AD associated with Brain 1-3, we further explored their potential for patient stratification to improve clinical outcomes in an AD drug trial (**Method 7e**). As illustrated in **Fig 6m-o**, participants with under-expression levels (MAE below the median) of Brain 1-3 demonstrated better cognitive performance than those with higher expression levels in each respective brain MAE. For example, Brain 2 showed the strongest differences on PACC between the under-expressed and over-expressed patient groups (t-statistics=3.519; P-value=0.004). Detailed statistics are presented in **Supplementary eTable 11**.

## Discussion

In this study, we introduce 11 novel AI-derived biomarkers to understand disease heterogeneity through the concept of pan-disease, which bridges transdiagnostic insights^78^ across multiple organ systems. Integrating AI with multi-omics and multi-organ biomedical data provides a holistic framework to model disease heterogeneity, further supporting the endophenotype hypothesis^2^. The 11 MAEs complement traditional disease classification systems and offer a data-driven representation for stratifying individuals at higher risk for specific dimensions of neurobiological and etiologic factors. These MAEs enhance our understanding of disease biology by uncovering their distinct phenotypic, genetic, and proteomic architectures. Importantly, they demonstrate differential predictive power for chronic disease incidents, mortality, AD progression, and medication status, offering a unique dimensional tool for advancing personalized medicine and drug development strategies. **Extended Data Fig. 5** summarizes the characteristics of the 11 MAEs.

### 11 MAEs capture reproducible and distinctive morphological changes for pan-disease of the brain, eye, and heart

We identified 11 MAEs characterized by reproducible and distinct imaging patterns that complement and challenge traditional disease classification frameworks. These imaging patterns effectively differentiate individuals with morphological changes from those without pathologies.

The 6 brain MAEs encapsulate various neuroanatomical changes, with Brain 1-3 displaying distinct patterns of brain atrophy, while Brain 4-6 are associated with conserved brain structures, marked by larger brain volumes. Many neurodegenerative diseases were associated with brain atrophy, which could be attributable to the atrophy patterns observed in Brain 1-3. For example, AD was widely known for global atrophy with a focal hub in the medial temporal lobe^79^ and hippocampus^80^. Heterogeneity analysis has unraveled multiple disease subtypes, demonstrating different atrophy patterns. For example, Young et al.^10^ applied the SUSTAIN model and found 3 distinct subtypes and trajectories of AD, which were termed typical, cortical, and subcortical atrophy subtypes. In another study, Yang et al.^9^ applied another AI method called Smile-GAN and identified 4 distinct AD subtypes: *i*) preserved brain volume, exhibits no significant atrophy across the brain compared to healthy controls; *ii*) mild diffuse atrophy, with widespread mild cortical atrophy without pronounced medial temporal lobe atrophy; *iii*) focal medial temporal lobe atrophy, showing localized atrophy in the hippocampus and the anterior-medial temporal cortex with relative sparing elsewhere; and *iv*) global atrophy pattern. Beyond the neuroanatomical heterogeneity, Vogel et al.^33^ also defined 4 distinct tau subtypes in AD, including limbic-predominant and medial temporal lobe-sparing patterns.

Global brain atrophy is less prominent in psychiatric disorders than in neurodegenerative diseases, though some conditions do exhibit brain volume loss, particularly in severe or chronic cases. In a case-control setting, schizophrenia shows widespread atrophy, mainly in the prefrontal cortex, temporal lobes, and hippocampus^81^. Disease subtypes identified by AI further showed that a subset of schizophrenia patients showed conserved brain volume. For example, Chand et al.^36^ applied the HYDRA model to define two subtypes of schizophrenia, with subtype 2 showing increased volume in the basal ganglia and internal capsule. This conserved brain volume subtype was corroborated by Jiang et al.^35^ using an independent dataset, which identified a similar pattern of increased striatal volume in their defined subtype 1. The relatively conserved brain volume patterns observed in several psychiatric disorders, including late-life depression^34^ and autism^82^, may be attributable to the imaging patterns observed in Brain 4-6. In addition, the “medicational compensation effect” concept with conserved brain volume in brain diseases may compensate for disease-induced damage through neuroplasticity or other adaptive mechanisms. This could be observed in specific subtypes/dimensions of brain diseases where, despite widespread atrophy, some brain regions (such as the striatum or hippocampus) maintain or increase their volume. For example, a recent study showed that AI-derived dimension 1 of major depressive disorder, characterized by preserved gray and white matter, showed a significant improvement in depressive symptoms following treatment with SSRI medication (51.1%) but limited changes following placebo (28.6%)^83^.

The impact of normal aging on global brain atrophy, particularly in brain subtypes 1-3, is evident from the strong association with brain BAG (**Fig. 3a**). In a recent study, we applied the Surreal-GAN model to a large and diverse population dataset (*N*=49,482) with brain MRI data and identified five distinct patterns of age-related brain atrophy. Notably, one of these dimensions, referred to as R2, exhibited a pattern reminiscent of AD-related medial temporal lobe atrophy, underscoring the overlap between normal aging and pathological brain changes. Finally, the 6 brain MAEs may also capture morphological changes in the central nervous system beyond the brain, such as alterations in the spinal cord or peripheral neural pathways^84^. These regions may exhibit degeneration or structural adaptations linked to neurodegenerative diseases or systemic conditions affecting the central nervous system.

The three eye MAEs showed distinct morphological changes, with the most prominent macular thickness and optic disk thinning in Eye 2 (**Fig. 1b**), corroborated by the strongest genetic overlaps between Eye 2 and eye BAG (**Fig. 4b**). A pattern of macula thickness thinning and optic disc thinning is often associated with a variety of eye diseases, typically involving degenerative processes or damage to the retina and optic nerve, such as age-related macular degeneration^85^. Eye 1 and Eye 3’s morphological patterns varied. For instance, Eye 1 showed a reduced macular thickness, whereas Eye 3 exhibited macular thickening (ML; **Fig. 1b**). Both Eye 1 and Eye 3 were associated with different forms of glaucoma (**Fig. 2b**; ICD code: H400 & H401)^86^, which manifests typically with retinal nerve fibre layer thinning; however, patients with certain eye diseases showed increased macular thickness. Diseases like diabetic macular edema^87^ can cause a thickening of the macula due to fluid accumulation, inflammation, or abnormal blood vessel growth. This can be supported by the association between Eye 3 and non-insulin-dependent diabetes mellitus in our PWAS analyses (**Fig. 2b**; ICD code: E119). Another unique feature of Eye 3 is the positive association of the left vertical cup-to-disc ratio (VcdrL: https://labs-laboratory.com/medicine/vertical_cup_to_disc_ratio_vcdr_left_f27857_0_0). Previous studies have found this association, most notably in glaucoma, where progressive damage to the optic nerve leads to an expansion of the optic cup relative to the optic disc. This indicates a loss of retinal ganglion cells and optic nerve fibers, resulting in the typical “cupping” phenomenon seen in glaucoma^88^.

The two heart MAEs also demonstrated distinct morphological changes. Overall, Heart 1 was characterized by increased wall thickness of the LV (e.g., WT_global: https://labs-laboratory.com/medicine/lv_mean_myocardial_wall_thickness_global_f24140_2_0), whereas Heart 2 showed increased volume of the LA and the stroke volume of the LA, as well as decreased ejection fraction volume of the LA (LAEF), opposite with Heart 1. Many heart diseases, such as hypertension and aortic stenosis, cause the heart muscle, particularly the left ventricle, to thicken (i.e., LV hypertrophy) as it works harder to pump blood against increased resistance^89^. The opposite direction of associations between Heart 1 and Heart 2 with metrics linked to the LA and RA, normally an indirect marker for the overall function and structure of the LV, may indicate distinct pathological factors related to the two dimensions. For example, a decreased ejection fraction is indicative of impaired left ventricular systolic function and is a hallmark of several heart conditions, including heart failure and myocardial Infarction^90^.

Our AI models analyzed the heterogeneous morphological changes of pan-disease affecting the three organs, identifying potentially distinct pathological and etiological processes associated with the 11 MAEs. This enabled a digitized, dimensional system to prioritize individuals with specific morphological changes.

### Opposite to Brain 5, Brain 1-3 serves as a risk dimension for AD progression, mortality, antipsychotic medications, and Solanezumab

Brain 3 is characterized as a risk dimension associated with AD progression (MCI→AD), mortality, and antipsychotic medication use. The global atrophy pattern of the brain is likely tied to neuroanatomical changes that predispose individuals to more severe cognitive decline^91^, higher mortality risk, and a greater need for antipsychotic interventions. Global cortical atrophy and hippocampus atrophy have been prominent biomarkers for MCI and AD. For example, Leung et al.^91^ used longitudinal data to evidence that hippocampal atrophy rates in MCI subjects accelerate by 0.22%/year^2^ on average. In our PWAS, the brain pattern of structural covariance (PSC; **Method 2a**; data-driven structural covariance network for C128_3: https://www.cbica.upenn.edu/bridgeport/MUSE/Left%20Hippocampus) spatially linked to the left hippocampus was significantly associated with Brain 3 (*β=-*0.077; P-value=1.89×10^−39^). The high relevance of AD with Brain 3 was further strengthened by the link with the CSF t-tau_181p_ (**Extended Data Fig. 1**d). Another study revealed that decreased survival was linked to atrophy in the temporal and frontal lobes, as well as widening of the Sylvian fissure and several ventricular measurements^92^. Finally, various psychiatric disorders exhibit shared global atrophy patterns^93^, suggesting that patients predominantly characterized by Brain 3 may have a more favorable response in future clinical trials and drug development. Crucially, in the initial clinical trial, the authors reported that no significant drug effect was achieved between the placebo and treatment groups for Solanezumab^6^. Our brain MAEs, particularly Brain 2, demonstrated the effectiveness of AI-driven population stratification, dividing treatment groups into two distinct subpopulations with potentially divergent cognitive decline trajectories (**Fig. 6m-o**). This insight is pivotal for future AD clinical trials, as these subgroups may represent different underlying disease mechanisms^94^, underscoring the need for personalized treatment strategies.

In contrast, Brain 5, characterized by conserved brain volume, may indicate a protective dimension for these risks. Brain 5 showed an HR of 0.69 [0.62, 0.78] for MCI→AD progression, indicating lower conversion risks (**Fig. 6e**). However, such a protective association was not observed for the CN→MCI progression. Several factors may explain this pattern. First, our brain MAEs were modeled using Surreal-GAN based on ICD-10 codes derived from hospital inpatient records and other resources, which may reflect a later stage in the progression of brain pan-disease. Consequently, these brain MAEs are less sensitive to early neurobiological changes within the AD continuum. Secondly, various compensatory mechanisms might also be at play. In the initial stages of neurodegenerative diseases like AD, the brain may demonstrate compensatory responses, leading to increased volume due to factors such as edema or neuroinflammation. In individuals with MCI, this increased volume can obscure underlying atrophy. However, as the disease advances, these compensatory effects wane, and brain atrophy becomes more pronounced, resulting in negative correlations with cognitive decline^95^. Similarly, the protective role of Brain 5 for AD was also supported by the positive association between Brain 5 and Aβ_1-42_ (**Extended Data Fig. 1**d), further corroborating a protective or compensatory response in the face of emerging pathology for Brain 5.

Their underlying genetic architectures can also further elucidate the contrast profiles between Brain 3 and Brain 5, showing spark differences in purifying selection signatures and pyrogenicity (**Fig. 4c & d**). The stronger negative purifying selection signature observed in Brain 5 compared to Brain 3, along with Brain 5’s stronger polygenicity, can be attributed to several factors related to evolutionary dynamics, genetic architecture, and the functional significance of the associated traits. First, Brain 5 may be associated with traits under strong evolutionary pressure to maintain function, resulting in more pronounced negative purifying selection. This type of selection removes deleterious alleles from the population, thus preserving critical functions (e.g., conserved brain volume). In contrast, Brain 3 may be less functionally constrained, allowing for a higher tolerance of genetic variation. The negative selection signature is common in complex human traits, as evidenced by Zeng et al. in their original paper on SBayesS^96^. On the other hand, the increased polygenic nature in Brain 5 may also indicate a higher genetic load^97^, where multiple alleles contribute cumulatively to brain function. This complexity can lead to a more robust system that is still subject to the constraints of purifying selection, ensuring that only advantageous variants are retained. To summarize, the distinct evolutionary pressures and genetic architectures of Brain 5 and Brain 3 may also influence their susceptibility to neurodegenerative diseases. The stronger purifying selection in Brain 5 could indicate a protective mechanism that mitigates risks associated with pathological conditions, while the increased polygenicity could make Brain 5 more adaptable to environmental or genetic changes over time.

### Brain 1 identifies the *FLRT2* gene as a novel druggable target for migraine treatment

We identified a multi-layer causal pathway prioritizing the *FLRT2* gene as a novel druggable target for migraine treatment (**Fig. 5e**). Migraine is a common brain disorder with a large genetic component; the *FLRT2* gene is not considered to be associated with migraines ^98^, albeit it does have implications in postnatal central nervous system development^99^, ischemic stroke^100^, and bipolar disorder^101^.

By contrast, our findings show that *FLRT2* does influence migraine disorders through its effects on brain morphology (Brain 1), specifically through the atrophy in the temporal and frontal lobes and the inferior cerebellum. This has significant clinical implications. In particular, this causal association suggests that individuals carrying specific variants of the *FLRT2* gene (e.g., these single SNPs in **Extended Data Fig. 4**b) may be at a higher risk for developing migraines, indicating a potential biomarker for early identification and risk assessment. Furthermore, the role of *FLRT2* in neuronal development points to its potential as a therapeutic target; interventions aimed at modulating FLRT2 pathways may offer new treatment options. The connection between structural brain changes and migraine pathophysiology highlights the importance of understanding how genetic factors contribute to the development of migraines. Integrating genetic and proteomic information with neuroimaging data could refine diagnostic criteria and therapeutic strategies, paving the way for personalized medicine approaches in migraine management. This comprehensive approach emphasizes the significant role of multi-omics data in driving novel drug and therapeutic discoveries. For instance, a recent study^102^ demonstrated the use of spatial proteomics to uncover a new treatment for toxic epidermal necrolysis, illustrating how integrating multi-omics insights can open promising avenues for clinical translation.

Identifying the *cis*- and *trans*-pQTL, on top of the eQTL, for FLRT2 provides direct evidence of genetic variation that modulates the levels or function of this protein, linking specific genetic loci to physiological changes in protein expression and, subsequently, to clinical outcomes^103^. Specifically, this suggests that genetic variations alter the risk of brain atrophy and migraine and affect how much or how efficiently the FLRT2 protein is produced or functions in brain tissue. This enhances our understanding of the causal pathway from gene to disease, as it demonstrates that genetic differences at the *FLRT2* locus lead to quantifiable changes in the protein that may drive or exacerbate the brain atrophy observed in Brain 1. This causal pathway also offers empirical support for the mediational model of the endophenotype hypothesis^7^, highlighting Brain 1 as a promising biomarker for guiding drug development efforts to treat related disorders.

### Eye 1 and Eye 3 show implications for a close eye-cancer relationship

Our results highlight crucial connections between different cancer types and Eye 1 and 3. This highlights the systemic effects of cancer and can be utilized to leverage eye morphology as a potential biomarker for early cancer detection and monitoring.

The eye morphological changes in Eye 1 and Eye 3 have implications in cancer, suggesting that alterations in structures such as the macula or optic disc may reflect ocular health and serve as biomarkers for cancer. For example, Eye 1 and malignant neoplasm of the corpus uteri (ICD code: C541) showed a positive association (β=0.059±0.0016, P-value=7.61×10^−8^; **Fig. 3b**). Our findings highlight potential drug repurposing opportunities for Eye 1 and Eye 3, particularly in the context of cancer therapies. The associations between these eye-derived morphological features and malignant neoplasms, supported by gene-drug-disease network analyses (**Fig. 4e**) and causal links to key proteins (**Fig. 5d**), suggest that eye endophenotypes might be early indicators of oncogenic processes. This relationship opens new avenues for exploring eye health as part of a multi-organ approach to detecting and managing cancer. The underlying hypothesis is that shared molecular and genetic pathways could be at play, influencing both eye morphology and cancer susceptibility, especially in aging. First, metastatic cancer, such as melanoma or breast cancer, can spread to the eye, affecting structures like the retina, choroid, or optic nerve, leading to changes such as thickening, swelling, or tissue atrophy^104,105^. Paraneoplastic syndromes, particularly cancer-associated retinopathy^106^, are another factor, where the immune system’s response to systemic cancer attacks retinal cells, causing significant ocular changes. Additionally, treatments for cancer, including chemotherapy and radiation, can result in complications like macular edema, optic neuropathy, and retinal degeneration, altering the eye’s structural morphology^107^. Moreover, genetic links are also emerging, where mutations in genes involved in both cancer and eye health, such as those controlling retinal development, suggest a molecular connection between eye changes and cancer progression. Finally, these connections may be more likely correlational; potential mechanisms could involve paraneoplastic syndromes, which reflect remote effects of tumors on the eye, or shared microvascular and metabolic pathways that influence both ocular and cancer pathophysiology.

Future research should delve deeper into these connections, investigating how specific eye endophenotypes could be used in cancer risk stratification, monitoring disease progression, and potentially guiding therapeutic interventions, thereby broadening the scope of precision medicine in aging and oncology.

### Heart 1 and Heart 2 are two monitorable dimensions for mortality and various cardiovascular conditions

We identified Heart 1 and Heart 2 as distinct dimensions associated with predicting mortality, multiple cardiovascular conditions, and related medication usage. These findings suggest that these dimensions could be instrumental in patient stratification, potentially guiding future drug development efforts to target specific cardiovascular conditions and optimize treatment strategies.

The two heart MAEs showed the highest reproducible patterns (**Fig. 1e**) compared to the other organs, offering strong evidence that two distinct dimensions exist in heart pan-disease. In particular, Heart 2 and Heart 1 showed higher HR estimates for predicting mortality than the other two organs. Cardiovascular diseases remain the leading cause of death globally^108^. As a result, heart-related MAEs are more likely to be associated with higher HR for mortality and adverse events than those associated with neurodegenerative or ophthalmic diseases. Heart 1 and Heart 2 likely capture systemic cardiovascular changes affecting the entire body, including the brain and eyes. In contrast, the brain and eye MAEs may represent more localized damage that, although severe, may not have the same broad systemic effects as heart conditions. Furthermore, while the brain and eye MAEs are crucial for cognitive and sensory functions, cardiovascular conditions can lead to multi-organ failure^109^ through mechanisms like reduced blood flow, increased inflammation, and oxygen deprivation. This widespread impact could result in more severe clinical outcomes, reflected in higher HRs for heart conditions. From a genetic standpoint, weaker purifying selection signatures of Heart 1 and Heart 2, especially compared to Eye 1 and Eye 3 (**Fig. 4c**), can result in higher genetic variability in heart MAEs. This could lead to more polygenic architectures where many genetic variants with small effects collectively contribute to the phenotypic presentation of heart diseases^110,111,112^ (**Fig. 4d**). The higher HRs observed for Heart 1 and Heart 2 compared to brain and eye MAEs might reflect this polygenic nature, as well as the complex interaction of environmental, lifestyle, and genetic factors in heart disease development.

The distinct clinical implications of the two heart MAEs suggest differential risk profiles for cardiovascular health and drug response (**Fig. 6**). Heart 1, which is more strongly associated with hypertension, hypercholesterolemia, and antihypertensive medications, aligns with traditional cardiovascular risk factors. This suggests that individuals dominated by the Heart 1 MAE may be more predisposed to metabolic and vascular abnormalities, likely benefiting from interventions targeting cholesterol levels and blood pressure management^113^. On the other hand, Heart 2, which is predictive of psychoactive substance use and the use of digoxin (critically not Heart 1) for heart failure treatment (incremental *R*^2^=30%; P-value<=2.05×10^−19^; *N*=4811; **Fig. 6l** & **Supplementary eFigure 11**) highlights a different risk dimension. The link with psychoactive substance use could reflect the broader role of lifestyle and substance exposure in cardiovascular health^114,115^. Clinically, this distinction between the two heart MAEs provides insight into personalized therapeutic approaches, where Heart 1 patients may respond better to antihypertensive and lipid-lowering therapies, whereas Heart 2 patients might benefit from treatments addressing heart failure and lifestyle factors contributing to substance abuse. This highlights the potential for stratifying cardiovascular patients based on their MAE profile for targeted interventions and precision medicine^116^.

### No organ system is an island

In addition to introducing the novel concept of pan-disease to explore disease heterogeneity, our study enhances multi-organ research by incorporating advanced AI techniques to provide a dimensional framework for more precise patient stratification and management.

Previous multi-organ research initially focused on linking IDPs across multiple organ systems. For example, we introduced a novel data-driven brain IDP – patterns of structural covariance^26^ – and established a brain-eye-heart axis^3^. McCracken et al.^117^ established a brain-heart-liver axis using MRI data from these organs using UKBB imaging data. Liu et al.^118^ also integrated genetics with 11 organ traits from abdominal MRI. Later studies focused on deriving individual-level imaging signatures across organ systems via AI. For instance, Tian et al.^22^ developed multi-organ BAGs from 9 organ systems, linking them to clinical traits, while Wen et al.^21^ expanded this by investigating the genetic architecture of these BAGs. More recently, multi-organ BAGs were also derived from proteomics data, further being linked to disease outcomes using UKBB data^119^. In addition, the brain-gut axis^120^ may be another data layer to human aging and disease, and future investigations, especially leveraging emerging microbiome data in the UK Biobank, could expand this work to incorporate gut-related mechanisms.

Our multi-organ MAEs offer a complementary and clinically focused advancement over BAGs^18,121,19,18,25,21,23^. While BAGs summarize the deviation between biological and chronological age and are useful for capturing global aging processes, they are inherently broad, often reflecting the cumulative effects of diverse and overlapping biological pathways. In contrast, MAEs are designed to be disease-specific, data-driven imaging biomarkers that encapsulate organ-level morphological variations more directly tied to individual disease processes. Clinically, this distinction translates to greater specificity. MAEs are more interpretable in the context of particular diseases (e.g., Alzheimer’s, cardiomyopathies, or ocular degeneration), enabling more precise risk stratification and potentially informing targeted interventions. MAEs also improve cross-organ resolution by separately modeling each organ’s structural alterations, rather than aggregating them into a single index as BAGs do. This organ-level granularity can reveal distinct morphometric signatures and interactions between organs that are missed by aging clocks. As such, we envision that aging clocks and disease-specific MAEs are integrated within a unified digital twin framework: BAGs provide a broad systemic health profile, while MAEs supply fine-grained, organ-targeted insights to support more personalized and effective clinical decision-making.

## Limitations & Outlook

carefully evaluated the generalizability of our GWAS findings to non-European groups. Future methodological advances are needed to extend the Surreal-GAN model to non-European populations to overcome this potential bias and ensure model fairness. Second, while we have evaluated the expression of the 6 brain MAEs in independent external datasets, further studies are needed to validate the heart and eye MAEs. We anticipate that future research will explore multi-organ, multi-omics approaches, providing more comprehensive models for studying human aging and disease. This direction promises to unlock deeper insights and refine our patient care and therapeutic development strategies across diverse health conditions. Future work could also focus on developing fully data-driven disease subtypes across multiple organ systems by leveraging shared populations with multi-organ data. This approach may uncover novel, system-level disease mechanisms and enable more precise cross-organ phenotyping.

## Methods

### Method 1: The MULTI consortium

The MULTI consortium is an ongoing initiative to integrate and consolidate multi-organ data, such as brain and heart MRI and eye OCT, with multi-omics data, including imaging, genetics, and proteomics. Building on existing consortia and studies, such as those listed below, MULTI aims to curate and harmonize the data to model human aging and disease across the lifespan. In this study, in total, individual-level data for 129,340 participants were analyzed, including 81,831 for multi-organ imaging data (brain and heart MRI and eye OCT), 83,322 for genetic data (imputed genotype and WGS), and 53,940 for plasma proteomics data (Olink and SomaScan). GWAS summary statistics from FinnGen and PGC were downloaded and harmonized for our post-GWAS analyses. A subset of the population also had cognitive data and neuropathological biomarkers for AD, including Aβ_1-42_ and tau_181p_ from CSF and plasma. RNA-seq and additional protein data were also curated and made available through the HPA and STRING platforms. Refer to **Supplementary eTable 1** for comprehensive information, including the complete list of data analyzed and their respective sample characteristics. Participants provided written informed consent to the corresponding studies. The MULTI consortium is approved by the Institutional Review Board at Columbia University (AAAV6751).

### UK Biobank

UKBB^122^ is a population-based research initiative comprising around 500,000 individuals from the United Kingdom between 2006 and 2010. Ethical approval for the UKBB study has been secured, and information about the ethics committee can be found here: https://www.ukbiobank.ac.uk/learn-more-about-uk-biobank/governance/ethics-advisory-committee.

This study collectively analyzed multi-organ imaging data, including 42,660 brain T1-weighted MRI scans (referred to as the *brain population*), 35,576 heart MRI scans (representing the *heart populations*), and 40,063 eye OCT images (the *eye populations*) at baseline, under Application Numbers: 35148 and 60698. The combined sample size across the three organ populations totaled 81,831. The T1-weighted MRI data underwent processing at the University of Pennsylvania for the brain PSCs, denoting data-driven structural network measurements via the MuSIC atlas and the iSTAGING consortium^26^. To train the Surreal-GAN model, we used the scale (C=128) brain PSCs, comparable to the number of heart and eye IDPs, detailed afterward. The 82 heart IDPs were downloaded directly from the UKBB website and derived from our prior study by Bai et al. ^27^ (Category ID: 157). We included heart IDPs with more than 35,000 participants, resulting in 80 heart IDPs to train the Surreal-GAN model. The 84 eye IDPs, derived from OCT imaging, were directly downloaded from our previous studies^28–30^ by Patel and colleagues via the UKBB Eye and Vision Consortium (Category ID: 100016; Return ID: 1875). Similarly, we constrained the eye IDPs with more than 20,000 participants, resulting in 64 eye IDPs in the Surreal-GAN model.

For the genetic data, we conducted a quality check on the imputed genotype data^122^ for the entire UKBB population (approximately ∼500k individuals). Subsequently, we merged the processed data with the *brain*, *eye & heart population* for all genetic analyses (*N*=81,831). Refer to **Method 6** for details. Our primary focus was on populations of European ancestry, with non-European ancestry populations included in sensitivity check analyses. The preprocessing for the proteomics data is detailed below.

Finally, we also included the Olink proteomics data released by the UK Biobank Pharma Proteomics Project (UKB-PPP). The original data included 2923 Olink plasma proteins for 53,016 participants. Details of the quality check procedure are presented in **Method 5**. In total, Multi-omics data from 125,406 UKBB participants were analyzed in the current study after quality checks.

### Baltimore Longitudinal Study of Aging

The main goal of the BLSA is to understand the normal aging process. Tracking physiological and cognitive changes over time aims to identify risk factors for age-related diseases, study patterns of decline, and discover predictors of healthy aging. BLSA^40,41^ brain MRI, and proteomics data (https://www.blsa.nih.gov/) were used to replicate the brain ProWAS results from the UKBB study. After quality checks in this study, we included 1114 brain MRI scans at baseline and measurements of 7268 plasma proteins from 924 participants quantified with the SomaScan v4.1 platform. Age (years), sex (male/female), race (white/non-white), and education level (years) were defined based on participant self-reports.

### Anti-Amyloid Treatment in Asymptomatic Alzheimer’s

The A4 study^6,42^ (https://atri.usc.edu/study/a4-study/) is a clinical trial study to test a specific way to prevent memory loss associated with AD (clinical trial number: NCT02008357). The A4 study focused on symptom-free adults at higher risk for AD to assess whether an investigational drug (i.e., Solanezumab) could slow memory decline linked to amyloid plaques in the brain. It also examined whether Solanezumab could delay AD progression, measuring related brain changes using imaging, blood biomarkers, and baseline PET scans to assess amyloid levels. This study analyzed 1,055 participants at baseline with brain MRI scans to derive the 6 brain MAEs. Longitudinal outcomes from the clinical trial, with the Preclinical Alzheimer’s Cognitive Composite (PACC) score as the primary measure over 312 weeks, were included. Specifically, there were 516 participants with 5161 longitudinal PACC measures in the treatment group and 591 participants with 5469 longitudinal PACC measures in the placebo group. The drug’s effect between groups (e.g., over-expressed vs. placebo) was evaluated at week 240.

### Alzheimer’s Disease Neuroimaging Initiative

ADNI (https://adni.loni.usc.edu/) includes patients from different stages of the disease progression: cognitively normal individuals (CN), those with MCI, and those with AD. This allows researchers to compare brain structure and function changes across the Alzheimer’s disease continuum. We included baseline brain MRI data of 1765 individuals, 9752 longitudinal follow-up scans (at least >5 scans for longitudinal analyses used in this study), and 1491 whole-genome sequencing data to generalize the GWAS signals from the UKBB data.

### FinnGen

The FinnGen^123^ study is a large-scale genomics initiative that has analyzed over 500,000 Finnish biobank samples and correlated genetic variation with health data to understand disease mechanisms and predispositions. The project is a collaboration between research organizations and biobanks within Finland and international industry partners. For the benefit of research, FinnGen generously made their GWAS findings accessible to the wider scientific community (https://www.finngen.fi/en/access_results). This research utilized the publicly released GWAS summary statistics (version R9), which became available on May 11, 2022, after harmonization by the consortium. No individual data were used in the current study.

FinnGen published the R9 version of GWAS summary statistics via REGENIE software (v2.2.4)^124^, covering 2272 DEs, including 2269 binary traits and 3 quantitative traits. The GWAS model encompassed covariates like age, sex, the initial 10 genetic principal components, and the genotyping batch. Genotype imputation was referenced on the population-specific SISu v4.0 panel. We included GWAS summary statistics for 521 FinnGen DEs in our analyses.

### Psychiatric Genomics Consortium

PGC^125^ is an international collaboration of researchers studying the genetic basis of psychiatric disorders. PGC aims to identify and understand the genetic factors contributing to various psychiatric disorders such as schizophrenia, bipolar disorder, major depressive disorder, and others. The GWAS summary statistics were acquired from the PGC website (https://pgc.unc.edu/for-researchers/download-results/), underwent quality checks, and were harmonized to ensure seamless integration into our analysis. No individual data were used from PGC. Each study detailed its specific GWAS models and methodologies, and the consortium consolidated the release of GWAS summary statistics derived from individual studies. In the current study, we included summary data for 4 brain diseases for which allele frequencies were present.

### Method 2: Multi-organ imaging features used to derive the MAEs via Surreal-GAN

(a) **Patterns of structural covariance of the brain**: Our earlier work^26^ applied the sopNMF method to a large-scale cohort of brain MRIs (*N*=50,699). This resulted in 2003 multi-scale brain PSCs, wherein the scale C ranged from 32 to 1024, expanding exponentially by a factor of 2. To train the Surreal-GAN model, we used the 128 brain PSCs from the scale of C=128.

PSCs represent data-driven structural networks that encapsulate coordinated neuroanatomical changes in brain morphology stemming from a mixture of normal aging, pathology, and unmodeled factors, such as environmental and genetic influences. Mathematically, the sopNMF algorithm is a deep learning-like stochastic approximation constructed and extended based on opNMF^126,127^. Consider a dataset comprising n MR images, each containing *d* voxels. The imaging data are represented as a matrix ***X***, where each column corresponds to a flattened image X = [x, x, …, x],X E Iff^dXn^. The sopNMF algorithm factorizes ***X*** into two low-rank matrices W E Iff^dXr^ and H E Iff^rXn^, subject to the constraints of *i*) non-negativity and *ii*) column-wise orthonormality. More mathematical details are presented in **Supplementary eMethod 2** and the original references^26,126,127^ (**Supplementary eTable 12**).

**(b) Imaging-derived phenotypes of the heart**: The 82 heart IDPs were directly downloaded from the UKBB website (Category ID: 157). Bai et al.^27^ employed a deep learning-based pipeline for analysing UKBB cardiac and aortic MR images, and derived a broad spectrum of structural and functional phenotypes for the heart and aorta. They investigated the correlations of these heart IDPs with factors such as sex, age, critical cardiovascular risk elements, and other non-imaging traits. For the 80 heart IDPs (after quality checks) included in our analyses, we further categorized them into 6 different IDP groups for visualization purposes, as shown in **Supplementary eTable 12**.

**(c) Imaging-derived phenotypes of the eye**: For the 88 eye IDPs, we used the derived OCT measurements from our previous studies^28–30^ (Category ID: 100079) and used the imaging quality scores (Field ID: 28552 and 28553) for quality check. We excluded 12,044 individuals whose scores were lower than 45 (Category ID: 100116).

OCT imaging is an advanced, non-invasive technology that generates three-dimensional images of the macula, crucial for detailed central vision. Abnormalities in macular thickness and morphology captured by OCT imaging are sensitive biomarkers of diabetic retinopathy, age-related macular degeneration, glaucoma, and various neurodegenerative diseases^128,129^. Our prior studies^28–30^ processed the OCT images to derive the 84 eye IDPs used in our analyses, and the results were subsequently returned to UKBB, making them accessible to the community. Ko et al.^29^ and Patel et al.^28^ from the UKBB Eyes and Vision Consortium (https://www.ukbiobankeyeconsortium.org.uk/; Category ID: 100016; Return ID: 1875) analyzed OCT images from over 60,000 individuals in the UKBB. They derived variables related to the thickness of the retinal pigment epithelium (excluding outliers and individuals with diseases affecting macular thickness). A subsequent investigation by Han et al.^30^ extracted optic nerve head morphology measures and performed a GWAS on these additional eye IDPs. For the 64 eye IDPs (after quality checks) included in our analyses, we further categorized them into 11 different IDP groups for visualization purposes (**Supplementary eTable 12**).

### Method 3: Surreal-GAN methodological considerations

**(a) Reference and target population definition**: We employed a weakly-supervised deep representation learning approach called Surreal-GAN^5^ to derive the 11 MAEs. Unlike traditional unsupervised clustering methods, which directly analyze the patient population, weakly-supervised techniques seek the *’1-to-k* mapping’ between the reference domain (CN) and the target domain (pan-disease PT)^15^. This approach effectively captures disease progression along the causal pathway and mitigates confounding factors (e.g., age, sex, or head size) unrelated to neurobiology and etiology.

In each organ-specific population, we defined the CN and PT populations for the Surreal-GAN model training using multiple resources from UKBB (**Fig. 1a**):

- CN: we excluded participants with any ICD codes from hospital inpatient records (Category ID: 2002), any medical conditions recorded in their health and medical history (Category ID: 100044), and certain mental health conditions (e.g., Category ID: 100060);
- PT for the brain pan-disease: We used the ICD-10 codes G (Diseases of the nervous system) and F (Mental and behavioral disorders) to define the brain pan-disease population.
- PT for the eye pan-disease: We used the ICD-10 codes H0-H5 (Diseases of the eye and adnexa) to define the eye pan-disease patient population.
- PT for the brain pan-disease: We used the ICD-10 code I (Diseases of the circulatory system) to define the heart pan-disease patient population.

After defining these populations, we merged them with the organ-specific imaging populations (**Supplementary eTable 1**) to train the 3 Surreal-GAN models.

We employed a grid search to select the optimal hyperparameters of the Surreal-GAN model:

- *k*: the number of pre-defined dimensions of pand-disease of each organ;
- lambda: the regularization term for controlling the orthogonality loss that boosts the separation of captured disease patterns in these dimensions;
- gamma: the regularization term for controlling the change loss that encourages sparsity and distance of transformations.

**(b) Methodological advances in Surreal-GAN**: The Surreal-GAN used in the current study has significant methodological improvements compared to its “*beta*” version proposed in our ICLR paper^4^. The original Surreal-GAN mode assumes that the *k* R-indices (i.e., the MAEs here) are independent, which limits its ability to characterize morphological patterns driven by associated underlying pathologies. If we sample the latent variable ***z***, indicating the transformation directions (i.e., *’1-to-k* mapping’), from a standard multivariate uniform distribution as in the original Surreal-GAN, the covariance of the derived R-indices will be the identity matrix, which leads to bias and decreased model performance when the ground-truth R-indices are correlated with each other. In a follow-up study^5^, we proposed a new version of Surreal-GAN to overcome this limitation, which constructed a parametrized latent distribution for ***z*** using a Gaussian copula, where the learnable parameters θ***_z_*** govern the correlations among the *k* dimensions. **Supplementary eMethod 3** details the methodological considerations and advances for the Sureal-GAN model. **Supplementary eFigure 14** shows the distribution of the 11 MAEs among different ethnic groups. **Supplementary eFigure 15** shows the convergence of the 3 models.

### Method 4: Phenotypic analysis

**(a) Primary phenome-wide associations (PWAS):** We performed different sets of primary PWAS that correlated the 11 MAEs with several sets of phenotypes to validate the derived MAEs: *i*) the 9 multi-organ BAGs derived by our previous study^21^, *ii*) ICD-10-based DEs from UKBB, *iii*) CSF and plasma neuropathological biomarkers of AD from the ADNI and UKBB data, and *iv*) the 8 cognitive scores from UKBB.

We employed a linear regression model for the multi-organ BAG PWAS using UKBB, with each MAE as the dependent variable and the BAG as the independent variable, adjusting for various covariates. For the DE PWAS, we used a logistic regression model, with the DE as the dependent variable and the MAE as an independent variable, adjusting for the same set of covariates. We included common covariates (e.g., age and sex) and organ-specific covariates in these analyses. We included covariates for the CSF/plasma neuropathological biomarker (e.g., tau_181_ and Aβ_1-42_), PWAS, and cognitive PWAS using data from UKBB and ADNI based on their availability in each dataset. For ADNI CSF PWAS, we included the AD diagnosis as an additional covariate in the linear regression model.

For the PWAS related to the brain MAEs in UKBB, we accounted for various covariates, including age (Field ID: 21003), sex (Field ID: 31), brain positioning in the scanner (lateral, transverse, and longitudinal; Field ID: 25756-25758), head motion (Field ID: 25741), intracranial volume, body weight (Field ID: 21002), height (Field ID: 50), waist circumference (Field ID: 48), BMI (Field ID: 23104), assessment center (Field ID: 54), along with the first 40 genetic principal components. For the PWAS of the eye MAEs, we controlled for age (Field ID:21003 at the eye assessment session), sex (Field ID: 31), body weight (Field ID: 21002), height (Field ID: 50), waist circumference (Field ID: 48), and first 40 genetic principal components as covariates. For the PWAS related to the heart MAEs, we included covariates for age (Field ID:21003), sex (Field ID: 31), body weight (Field ID: 21002), assessment center (Field ID: 54), height (Field ID: 50), waist circumference (Field ID: 48), and first 40 genetic principal components, smoking status (Field ID: 20116), diastolic (Field ID: 4079), and systolic (Field ID: 4080) blood pressure, assessment center (Field ID: 54), and BMI (Field ID: 23104).

**(b) Secondary PWAS**: A secondary PWAS was conducted to link the 11 MAEs with the other 117 phenotypes accessible in our downloaded UKBB data. The same linear regression model mentioned above was used. We explicitly excluded the brain, eye, and heart IDPs available at UKBB from this PWAS to prevent circular bias with our MAEs^130^.

### Method 5: Proteomic analyses

We downloaded the original data (Category code: 1838), which was analyzed and made available to the community by the UKB-PPP^131^. The initial quality check was detailed in the original work^32^; we performed additional quality check steps as below. We focused our analysis on the first instance of the proteomics data (“instance”=0). Subsequently, we merged the Olink files containing coding information, batch numbers, assay details, and limit of detection (LOD) data (Category ID: 1839) to match the ID of the proteomics dataset. We eliminated Normalized Protein eXpression (NPX) values below the protein-specific LOD. Furthermore, we restricted our analysis to proteins with sample sizes exceeding 10,000. We used the quality-checked proteomic data for three sets of analyses in this study: *i*) proteome-wide associations (ProWAS), *ii)* proteome- and genome-wide associations (ProGWAS), and *iii*) Mendelian randomization (**Method 6**).

**(a) Proteome-wide associations (ProWAS):** We performed ProWAS by linking the 11 MAEs to 2923 unique plasma proteins from 53,016 participants (10,018<*N*<39,489 per protein after quality check) using the Olink platform.

For the brain ProWAS, we accounted for various covariates, including age (Field ID: 21003), sex (Field ID: 31), brain positioning in the scanner (lateral, transverse, and longitudinal; Field ID: 25756-25758), head motion (Field ID: 25741), intracranial volume, body weight (Field ID: 21002), height (Field ID: 50), waist circumference (Field ID: 48), BMI (Field ID: 23104), assessment center (Field ID: 54), protein batch number (Category ID: 1839), limit of detection (LOD; Category ID: 1839), along with the first 40 genetic principal components. For the eye ProWAS, we controlled for age (Field ID:21003 for eye assessment instance), sex (Field ID: 31), body weight (Field ID: 21002 for eye assessment), height (Field ID: 50 for eye assessment), waist circumference (Field ID: 48 for eye assessment), protein batch number (Category ID: 1839), limit of detection (LOD; Category ID: 1839), and first 40 genetic principal components as covariates. For the heart ProWAS, we included covariates for age (Field ID:21003), sex (Field ID: 31), body weight (Field ID: 21002), height (Field ID: 50), waist circumference (Field ID: 48), and first 40 genetic principal components, smoking status (Field ID: 20116), diastolic (Field ID: 4079), systolic (Field ID: 4080) blood pressure, assessment center (Field ID: 54), and BMI (Field ID: 23104), protein batch number (Category ID: 1839), and LOD (Category ID: 1839). Multiple comparisons were performed using Bonferroni corrections based on the number of proteins and MAEs. We also used the time interval as an alternative covariate in the model for image data from organs that were not collected at the same visit as the proteomics data. The ProWAS signals using UKBB Olink data were then compared with SomaScan data collected from BLSA.

**(b) Tissue/organ-specific map of the expression of the human proteome:** We annotated the tissue/organ-specific expression patterns of significant proteins using the Human Protein Atlas (HPA: https://www.proteinatlas.org/)^132^ platform (version: 23). HPA curated the RNA and protein data from multiple sources, including the GTEx^133^, and FANTOM5^134^ datasets, to comprehensively assess tissue and/or single-cell (81 cell types from 31 datasets) expression profiles for a full set of human proteins. The protein data encompasses 15,303 genes for which antibodies are available. The RNA expression data is obtained from deep sequencing of RNA (RNA-seq), including at single-cell resolutions, across different tissue types. The methodology for determining the expression of the protein is detailed in the original publication^132^. In addition, HPA also performed cross-species analysis for the brain RNA expression using resources from different sources, such as the HPA pig brain and mouse RNA-seq data, GTEx human brain RNA-seq data, and FANTOM5 Human brain CAGE data. Importantly, proteins are often simultaneously over-expressed in various tissues or organs (i.e., lack of organ-specificity). Our main objective was to determine if the tested protein exhibited over-expression in the brain, eye, and heart. If it was not over-expressed in any of these three organs, we highlighted its expression in other tissues with the highest evidence of over-expression.

**(c) Proteome- and genome-wide associations (ProGWAS):** We also performed large-scale ProGWAS to link the 2923 proteins to common SNPs. Refer to **Method 6a** for genetic quality check pipelines and the GWAS model used. This analysis was guided by a previous study^73^ and was meant to use the derived GWAS summary data in our multi-layer Mendelian randomization analysis (**Method 6i**). To validate the ProGWAS results in our study, we demonstrated this by comparing our results using FLRT2 protein with those of Sun et al. (**Supplementary eNote 7 and eFigure 9**), showing a high degree of consistency.

**(d) Protein-protein interaction network analysis:** Using OmicVerse^135^ and the STRING database^136^, we performed protein-protein function and interaction inference, including direct physical binding and indirect interactions like shared biological pathways or cellular processes. STRING^137^ (version 12.0) provides streamlined access to a comprehensive, quality-controlled database of protein-protein associations across various organisms (human sapiens in our analyses). Functional enrichments in the identified PPI network were performed regarding gene sets defined from different resources, including GO terms, the KEGG pathway, etc.

### Method 6: Genetic analyses

We used the imputed genotype data for all genetic analyses. Our quality check pipeline resulted in 33,570 participants for the *brain population*, 30,260 for the *heart population*, and 36,659 for the *eye population* with European ancestry in UKBB (6,477,810 SNPs passing quality checks). In addition, ADNI WGS data resulted in 1555 participants for the brain GWAS, with 24,194,338 SNPs passing quality checks (**Supplementary eNote 4** and **eFigure 13**). We summarize our genetic quality check steps. First, we skipped the step for family relationship inference^138^ because the linear mixed model via fastGWA^139^ inherently addresses population stratification, encompassing additional cryptic population stratification factors. We then removed duplicated variants from all 22 autosomal chromosomes. Individuals whose genetically identified sex did not match their self-acknowledged sex were removed. Other excluding criteria were: *i*) individuals with more than 3% of missing genotypes; *ii*) variants with minor allele frequency (MAF; dosage mode) of less than 1%; *iii*) variants with larger than 3% missing genotyping rate; *iv)* variants that failed the Hardy-Weinberg test at 1×10^−10^. To further adjust for population stratification,^140^ we derived the first 40 genetic principal components using the FlashPCA software^141^. Details of the genetic quality check protocol are described elsewhere^26,23,21,34,94^.

**(a) GWAS:** We applied a linear mixed model regression to the European ancestry populations using fastGWA^139^ implemented in GCTA^57^. For each GWAS, we corrected for common covariates (e.g., age and sex) and organ-specific covariates (e.g., MRI scanner positions for the brain).

**Brain MAE GWAS**: The brain MAE GWAS controlled for confounding factors, including age (Field ID:21003), age-squared, sex (Field ID:31), age x sex interaction, age-squared x sex interaction, the first 40 genetic principal components, and total intracranial volume, the scanner position in lateral, transverse, and longitudinal directions (Field ID:25756-25758), assessment center (Field ID: 54), and BMI (Field ID: 23104), guided by earlier brain imaging GWASs^142^.

**Heart MAE GWAS**: The model included age (Field ID: 21003), age-squared, sex (Field ID: 31), age x sex interaction, age-squared x sex interaction, the first 40 genetic principal components, body weight (Field ID: 21002), height (Field ID: 50), and waist circumference (Field ID: 48), BMI (Field ID: 23104), smoking status (Field ID: 20116), assessment center (Field ID: 54), heart rate (Field ID: 12673), diastolic (Field ID: 12675), and systolic (Field ID: 12674) blood pressure, peripheral pulse pressure (Field ID: 12676), central pulse pressure (Field ID: 12678), body surface area (Field ID: 22427), average heart rate (Field ID: 22426), as covariates.

**Eye MAE GWAS**: The model included age (Field ID: 21003), age-squared, sex (Field ID: 31), age x sex interaction, age-squared x sex interaction, the first 40 genetic principal components, body weight (Field ID: 21002), height (Field ID: 50), assessment center (Field ID: 54), and waist circumference (Field ID: 48) as covariates. For the abovementioned three GWASs, we employed a stringent genome-wide P-value threshold (5×10^−8^/11) using Bonferroni correction based on the number of total MAEs (*N*=11) to ensure stringent statistical rigor.

We scrutinized the robustness of our MAE GWASs through several sensitivity analyses. These checks included: *i*) estimation of LDSC intercept, *ii*) a split-sample GWAS that randomly divided the entire heart population into two groups with no significant differences in sex and age, *iii)* sex-stratified GWAS conducted separately for males and females, and *iv*) a non-European GWAS to gauge the generalizability of GWAS signals identified in populations of European ancestry. We performed one additional sensitivity check for the brain MAE GWAS by generalizing the results to the WGS data from ADNI.

**ProGWAS**: Our ProGWAS followed the protocol of a recent study^73^ investigating plasma proteins’ associations with genetics and health-related traits. We controlled the following covariates in our fastGWA model: age (Field ID: 21003), age-squared, sex (Field ID: 31), age x sex interaction, age-squared x sex interaction, the first 40 genetic principal components, body weight (Field ID: 21002), height (Field ID: 50), assessment center (Field ID: 54), waist circumference (Field ID: 48), diastolic (Field ID: 12675), and systolic (Field ID: 12674) blood pressure. To identify the pQTLs in our ProGWAS, we pinpointed independent signals as defined in FUMA (refer to the section on **Annotation of Genomic Loci**). In line with previous research^32^, we defined *cis*-pQTLs for genomic loci where the top lead SNPs are located within 1 Mb of the gene encoding the corresponding protein. Conversely, loci beyond this range were classified as *trans*-pQTLs (e.g., **Fig. 5e**).

**Annotation of genomic loci**: For the MAE GWASs, genomic loci were annotated using FUMA^143^. For genomic loci annotation, FUMA initially identified lead SNPs (correlation *R*^2^ ≤ 0.1, distance < 250 kilobases) and assigned them to non-overlapping genomic loci. The lead SNP with the lowest P-value (i.e., the top lead SNP) represented the genomic locus. Further details on the definitions of top lead SNP, lead SNP, independent significant SNP, and candidate SNP can be found in **Supplementary eMethod 1**. For visualization purposes in **Fig. 4**, we have mapped the top lead SNP of each locus to the cytogenetic regions based on the GRCh37 cytoband.

**(b) PheWAS:** We used the GWAS Atlas^56^ platform to conduct an online PheWAS look-up analysis for the top lead SNP within each genomic locus; LD was fully considered. To facilitate this, we developed an “in-house web crawler” designed to automate searches on the PheWAS webpage: https://atlas.ctglab.nl/PheWAS. The search threshold was set at a P-value of 5×10^−8^. The GWAS Atlas PheWAS categorized these traits into broad domains/categories that primarily affect respective organ systems. This PheWAS was conducted on September 16, 2023.

**(c) SNP-based heritability**: We calculated the SNP-based heritability (h^2^) using the GCTA software, which employs raw individual genotype data to generate a genetic relationship matrix, addressing the “missing heritability” issue^57^. From our experience, individual-level data approaches (e.g., GCTA) generally obtained higher h^2^ estimates than summary-level methods (e.g., LDSC), a trend observed in our previous findings^21,16^ and those of other research groups^142^. Bonferroni correction based on the number of MAEs was applied to denote statistical significance.

**(d) Selection signature (*S*) and polygenicity estimate (*Pi*)**: We then used SBayesS^59^ to estimate two additional genetic parameters that unveil the genetic architecture of the MAEs. SBayesS is designed to estimate key parameters related to the genetic architecture of complex traits. It uses a Bayesian mixed linear model^96^ and requires only GWAS summary statistics for SNPs and LD information from a reference sample to perform these analyses. These parameters include polygenicity (*Pi*), and the relationship between minor allele frequency (MAF) and effect size (*S*), i.e., the selection signature. We used the software’s pre-computed sparse LD correlation matrix derived from the European ancestry by Zeng et al.^59^, and ran the *gctb* command^96^ using the argument *--sbayes S*. All other arguments were set by default.

**(e) Gene-drug-disease network**: We defined a gene-drug-disease network by examining the enrichment of the gene set linked to the 11 MAEs within specific drug categories from the DrugBank database^62^ using the GREP software^144^. First, we defined the MAE-specific gene set by three different gene mapping approaches: *i*) physical position, *ii*) eQTL, and *iii*) chromatin interactions (default parameters in FUMA). We then used these gene sets as input for GREP and conducted Fisher’s exact tests to determine whether these gene sets were enriched in gene sets defined by drugs within clinical indication categories for specific diseases or conditions (based on the ICD code).

**(f) Cell- and Tissue-specific partitioned heritability estimate**: We investigated different tissue types contributing to the heritability of the MAEs. To achieve this, the partitioned heritability analysis through stratified LD score regression assesses the extent of heritability enrichment attributed to predefined and annotated genome regions and categories^68^. This analysis considers three sets of analyses considering different tissue types: *i*) three cell types from Cahoy et al.^145^, *ii*) 498 multi-tissue chromatin-based annotations from peaks from six epigenetic marks using data from ROADMAP^146^ and ENTEx^147^, and *iii*) 205 multi-tissue gene expression data estimates using data from GTEx V8^148^ and “Franke lab” dataset. Detailed methodologies for the stratified LD score regression are outlined in the original work^68^. LD scores and allele frequencies for European ancestry were acquired from a predefined version based on data from the 1000 Genomes project.

**(g) Genetic correlation**: We estimated the genetic correlation (*g_c_*) between each MAE pair using the LDSC software. Precomputed LD scores from the 1000 Genomes of European ancestry were employed, maintaining default settings for other parameters in LDSC. It’s worth noting that LDSC corrects for sample overlap, ensuring an unbiased genetic correlation estimate^149^. Statistical significance was determined using Bonferroni correction. Three sets of analyses were performed: *i*) pair-wise MAEs, *ii*) the 11 MAEs vs. the 9 multi-organ BAGs, and *iii*) the 11 MAEs vs. the 521 FinnGen DEs and 4 PGC DEs.

**(h) Two-sample bidirectional Mendelian randomization**: We hypothesized that proteins and MAEs, acting as intermediate phenotypes (i.e., the endophenotype hypothesis^150^; refer to **Supplementary eFigure 8** for a schematic illustration of our hypothesis), reside along the causal pathway from underlying genetics to chronic DEs. We constructed a multi-layer causal network using multi-omics data (e.g., imaging, genetics, and proteomics).

To this end, we constructed 12 bi-directional causal networks: 1) *Brain2Protein*, 2) *Protein2Brain*, 3) *Eye2Protein*, 4) *Protein2Eye*, 5) *Heart2Protein,* 6) *Protein2Heart,* 7) *Brain2DE*, 8) *DE2Brain,* 9) *Eye2DE*, 10) *DE2Eye,* 11) *Heart2DE*, 8) *DE2Heart.* These networks used summary statistics from our MAE GWAS analyses in the UKBB, the FinnGen^123^, and the PGC^125^ study for the 11 MAEs and 525 DEs. For example, the *Brain2DE* causal network employed the 11 MAEs from UKBB as exposure variables and the 525 DEs from FinnGen and PGC as outcome variables. The systematic quality-checking procedures to ensure unbiased exposure/outcome variable and instrumental variable (IVs) selection are detailed below.

We used a two-sample Mendelian randomization approach implemented in the *TwoSampleMR* package^72^ to infer the causal relationships within these networks. We employed five distinct Mendelian randomization methods, presenting the results of the inverse variance weighted (IVW) method in the main text and the outcomes of the other four methods (Egger, weighted median, simple mode, and weighted mode estimators) in the supplement. The STROBE-MR Statement^151^ guided our analyses to increase transparency and reproducibility, encompassing the selection of exposure and outcome variables, reporting statistics, and implementing sensitivity checks to identify potential violations of underlying assumptions. First, we performed an unbiased quality check on the GWAS summary statistics. Notably, the absence of population overlapping bias^152^ was confirmed, given that FinnGen and UKBB participants largely represent populations of European ancestry without explicit overlap. PGC GWAS summary data were ensured to exclude UKBB participants. For networks between the proteins and MAEs from UKBB, we reran the ProGWAS and ensured no overlapping populations. Furthermore, all consortia’s GWAS summary statistics were based on or lifted to GRCh37. Subsequently, we selected the effective exposure variables by assessing the statistical power of the exposure GWAS summary statistics in terms of instrumental variables (IVs), ensuring that the number of IVs exceeded 8 before harmonizing the data. Crucially, the function “*clump_data*” was applied to the exposure GWAS data, considering LD. The function “*harmonise_data*” was then used to harmonize the GWAS summary statistics of the exposure and outcome variables. This yielded fewer IVs than the original included MAEs/DEs/proteins. Bonferroni correction was applied to all tested traits based on the number of effective DEs/MAEs/proteins or diseases, whichever was larger.

Finally, we conducted multiple sensitivity analyses. First, we conducted a heterogeneity test to scrutinize potential violations of the IV’s assumptions. To assess horizontal pleiotropy, which indicates the IV’s exclusivity assumption^153^, we utilized a funnel plot, single-SNP Mendelian randomization methods, and the Egger estimator. Furthermore, we performed a leave-one-out analysis, systematically excluding one instrument (SNP/IV) at a time, to gauge the sensitivity of the results to individual SNPs.

**(i) PRS calculation**: The PRS was computed using split-sample sensitivity GWASs for the 11 MAE GWASs. The PRS weights were established using split1/discovery GWAS data as the base/training set, while the split2/replication GWAS summary statistics served as the target/testing data. Both base and target data underwent rigorous quality control procedures involving several steps: *i*) excluding duplicated and ambiguous SNPs in the base data; *ii*) excluding high heterozygosity samples in the target data; and *iii*) eliminating duplicated, mismatching, and ambiguous SNPs in the target data.

After completing the QC procedures, PRS for the split2 group was calculated using the PRS-CS^154^ method. PRC-CS applies a continuous shrinkage prior, which adjusts the SNP effect sizes based on their LD structure. SNPs with weaker evidence are “shrunk” toward zero, while those with stronger evidence retain larger effect sizes. This avoids overfitting and improves prediction performance; no clumping was performed because the method takes LD into account. The shrinkage parameter was not set, and the algorithm learned it via a fully Bayesian approach.

### Method 7: Prediction and association analyses for the risk of DEs, AD progression, mortality, history of medication use, and preclinical AD drug effects

We investigated the clinical promise of the proposed MAEs in three sets of prediction analyses: *i)* survival analysis for the risk of DEs based on the ICD-10 code, ensuring each DE had at least 20 patients in each organ-specific population, *ii*) survival analysis for predicting the progression of AD progression: CN → MCI → AD, *iii*) survival analysis for predicting the risk of mortality, and *iv*) logistic regression to quantify the additional power (i.e., incremental *R*^2^).

**(a) Survival analysis for the incidence of DEs**: Using UKBB ICD-based DEs, we employed a Cox proportional hazard model while adjusting for covariates(i.e., age and sex) to test the associations. The covariates were included as additional right-side variables in the model. The hazard ratio (HR), exp(β*_R_*), was calculated and reported as the effect size measure that indicates the influence of each MAE on the risk of DEs. To train the model, the “time” variable was determined by calculating the difference between the date of the first in-patient diagnosis (Field ID: 41280) for a specific DE for cases (or the censoring date for non-cases) and the date attending the assessment center (Field ID: 53). We excluded the participants whose “time” variable were negative (i.e., diagnosed with the disease before entering the study). After entering the study, participants who received an ICD-based disease diagnosis were classified as cases.

**(b) Survival analysis for AD progression**: Using 9752 longitudinal brain MRI scans from ADNI (547 CN, 875 MCI, and 343 AD at baseline) and the same Cox model, we evaluated the risk of AD progression for the 6 brain MAEs. Further, to quantitatively assess prognostic performances with the 6 brain MAEs as features, we progressively added the most predictive brain MAE, on top of age and sex, to the Cox model to understand its optimal performance. The concordance index (C-index) was utilized to quantify the performance of risk prediction in a 100-repetition & 20% holdout cross-validation. For the progression of MCI → AD, the “time” variable was determined by calculating the difference between the age at AD diagnosis during follow-up for cases (or the censoring age for non-cases) and the age at baseline. Participants who received an AD diagnosis during the follow-up period were classified as cases. For the cumulative analyses, 5-fold cross-validation was run 100 times to derive the concordance index on validation sets.

**(c) Survival analysis for mortality risk**: Using the same Cox model and UKBB data, we also predicted the mortality risk of the 11 MAEs. To train the model, the “time” variable was determined by calculating the difference between the date of death (Field ID: 40000) for cases (or the censoring date for non-cases) and the date attending the assessment center (Field ID: 53). Participants who passed away after enrolling in the study were classified as cases.

**(d) Logistic regression for the pseudo incremental *R***^2^: We evaluated the predictive power of 11 MAEs on medication status at two levels of analysis, defining medication status. First, using a logistic regression model and UKBB data, we associated the medication status for four drug categories: *i*) antipsychotic medications (Field ID: 20466), *ii*) blood pressure medications (Field ID: 6177 for males & 6153 for females), *iii*) cholesterol-lowering medication (Field ID: 6177 for males & 6153 for females), and insulin (Field ID: 6177 for males & 6153 for females). For participants not on blood pressure, cholesterol-lowering, and insulin medications, we defined non-cases (CN) as individuals who had never taken the following additional drug: hormone replacement therapy. Additionally, we excluded participants who responded with ‘None of the above’ or ‘Prefer not to answer’ during the touchscreen questionnaire. We selected participants who reported taking any of these medications for each drug case during any study session. Secondly, to further validate the predictive capacity of the 11 MAEs for drug status, we investigated their association with 164 individual medications selected from UKBB (Field ID: 20003) based on a minimum of 1000 cases. The non-cases (CN) comprised individuals with no recorded history of exposure to the 6745 available drugs.

To derive the incremental *R*^2^, we built a null and an alternative logistic regression using the *statsmodels* package. The null model predicted medication status based on age and sex as the features, while the alternative model included each MAE as an additional feature. The difference between the *R*^2^ pseudo values for the alternative and null models reflected the incremental *R*^2^ explained by the MAE. Bonferroni correction was performed based on the number of MAEs for each drug.

**e) Natural cubic splines for modeling drug effects on the under-expressed, over-expressed patient groups in Brain 1-3 and the placebo group**: In our survival analysis of MCI → AD progression, we observed a significantly higher risk in Brain 1-3. Therefore, we used clinical trial data from the A4 study^6^ to investigate whether our proposed brain MAE could serve as a novel tool for population selection in future AD clinical trials. The original trial did not demonstrate cognitive decline slowing compared to the placebo over 240 weeks, with the treatment group even showing a slight worsening in the PACC score. We hypothesized that significant heterogeneity exists in drug responses within the treatment group. To test this, we conducted three comparisons using natural cubic spline modeling: i) under-expressed (e.g., < Brain 1 median) vs. over-expressed (e.g., > Brain 1 median), to assess differential drug effects; ii) under-expressed vs. placebo, to evaluate whether patients with lower brain MAE expression responded better (i.e., beneficial effect) to the drug than the placebo group; and iii) over-expressed vs. placebo, to determine if patients with higher brain MAE expression exhibited worsened cognition (i.e., detrimental effect) compared to placebo (also stratified by the brain MAE for fair comparison).

From a statistical perspective, this is to model repeated measures (i.e., PACC as the primary trial outcome for global cognition) as a continuous outcome in a mixed-effect model. We used the same method proposed by the original work from Donohue et al.^155^, in which the authors proposed a constrained longitudinal data analysis with natural cubic splines that treated time as continuous and used test version effects to model the mean over time for PACC. Fixed effects included the following terms: i) spline basis expansion terms (two terms), ii) interaction of the spline basis expansion terms with treatment (two terms), iii) the version of the PACC test implemented, iv) baseline age, v) education, vi) APOE4 carrier status (yes/no), and vii) baseline florbetapir cortical SUVr value. Following the guidance of the original method, several variance-covariance structures were sequentially assumed until the model converged with the heterogeneous Toeplitz. At week 240, we compared the mean PACC values between each pair of the two groups.

## Data Availability

The GWAS summary statistics and pre-trained AI models from this study are publicly accessible via the MEDICINE Knowledge Portal (https://labs-laboratory.com/medicine/) and Synapse (https://www.synapse.org/Synapse:syn64923248/wiki/630992). Our study used data generated by the TCGA Research Network (https://www.cancer.gov/tcga), the human protein atlas (HPA: https://www.proteinatlas.org), and the STRING data (https://string-db.org/). The two platforms curated and consolidated publicly available (single-cell) RNA-seq and protein data, including the GTEx project (https://gtexportal.org/home/). Genomic loci annotation used data from FUMA (https://fuma.ctglab.nl/). PheWAS used data from the GWAS Atlas platform (https://atlas.ctglab.nl/PheWAS). GWAS summary data for the DEs were downloaded from the official websites of FinnGen (R9: https://www.finngen.fi/en/access_results) and PGC (https://pgc.unc.edu/for-researchers/download-results/). Individual data from UKBB can be requested with proper registration at https://www.ukbiobank.ac.uk/. Data from ADNI can be requested with proper registration at https://adni.loni.usc.edu/. The gene-drug-disease network used data from the DrugBank database (v.5.1.9; https://go.drugbank.com/). The analysis for partitioned heritability estimates used data from ROADMAP (https://egg2.wustl.edu/roadmap/web_portal/) and ENTEx (https://www.encodeproject.org/). All unrestricted data supporting the findings are also available from the corresponding author upon request.

## Code Availability

The software and resources used in this study are all publicly available:

- Surreal-GAN: https://github.com/zhijian-yang/SurrealGAN, Disease heterogeneity analysis;
- MUSE: https://github.com/CBICA/MUSE, RAVENS map extraction;
- SOPNMF: https://github.com/anbai106/SOPNMF, Brain PSC extraction;
- ukbb_cardiac: https://github.com/baiwenjia/ukbb_cardiac, Heart IDP extraction;
- FUMA: https://fuma.ctglab.nl/, Gene mapping, genomic locus annotation;
- GCTA: https://yanglab.westlake.edu.cn/software/gcta/#Overview, Heritability estimates & fastGWA;
- LDSC: https://github.com/bulik/ldsc, genetic correlation
- TwoSampleMR: https://mrcieu.github.io/TwoSampleMR/index.html, Mendelian randomization;
- PRS-CS: https://github.com/getian107/PRScs, PRS calculation;
- SBayesS: https://cnsgenomics.com/software/gctb/#Overview, the three key parameter estimates;
- Lifelines: https://lifelines.readthedocs.io/en/latest/, Survival analysis;
- Statsmodels: https://www.statsmodels.org/stable/index.html, Medication prediction, BWAS, PWAS, and ProWAS;
- OmicVerse: https://omicverse.readthedocs.io/en/latest/; Protein-protein interaction.

## Authors’ contributions

Dr. Wen has full access to all the study data and is responsible for its integrity and accuracy.

*Study concept and design*: W.J.

*Drafting of the manuscript*: W.J.

*Critical revision of the manuscript for important intellectual content*: All authors

*Statistical analysis*: W.J. ran all main analyses; P.A and A.F. helped with analyses related to FinnGen and PGC using GWAS summary statistics; D.M. ran the ProWAS for the SomaScan proteomic data in the BLSA sample.

*Detailed contributions*: W.J. leads the MULTI consortium; D.C. leads the iSTAGING consortium; E.G. and S.D. conducted the image preprocessing pipelines for the brain MRI from the A4 study; B.W. derived the original heart IDPs; P.J.P. derived the original eye IDPs; W.A.K. represents the BLSA study; Z.A. and T.Y. derived the nine multi-organ BAGs; Y.Z. developed the Surreal-GAN model.

## Acknowledgments

This research is supported through funding from the NIH-supported MULTI Consortium (W.J.; grant number: RF1AG092412). The MULTI consortium (J.W.; UK Biobank Application Number: 647044) aims to integrate multi-organ imaging and multi-omics data to advance our understanding of human aging and disease mechanisms. This study used the UK Biobank resource under Application Numbers 647044 and 35148 (D.C.; grant number: RF1AG054409) and Application Number: 60698 (Z.A.). D.M., J.C., and W.K. were supported in part by the National Institute on Aging (NIA) Intramural Research Program (IRP) of the NIH. We want to express our sincere gratitude to the UK Biobank team for their invaluable contribution to advancing clinical research in our field (https://www.ukbiobank.ac.uk/). We also acknowledge the data sharing from the UKBB Eye and Vision Consortium (https://www.ukbiobank.ac.uk/enable-your-research/approved-research/genetic-contribution-to-vision-loss-and-disability-the-uk-biobank-eye-vision-consortium; Return ID: 1875) and UKB-PPP consortium (https://registry.opendata.aws/ukbppp/; Category code: 1838) to share the returned data with the community. We thank the BLSA participants and staff for their participation and continued dedication. The BLSA protocol was approved by the Institutional Review Board of the National Institute of Environmental Health Science, National Institutes of Health (03AG0325). Part of the data used was obtained from the ADNI database. The investigators within the ADNI contributed to the design and implementation of ADNI and/or provided data but did not participate in the analysis or writing of this report. A complete listing of ADNI investigators can be found at http://adni.loni.usc.edu/wpcontent/uploads/how_to_apply/ADNI_Acknowledgement_List.pdf. The A4 Study was a secondary prevention trial in preclinical Alzheimer’s disease, aiming to slow cognitive decline associated with brain amyloid accumulation in clinically normal older individuals. The A4 Study was funded by a public-private-philanthropic partnership, including funding from the National Institutes of Health-National Institute on Aging, Eli Lilly and Company, Alzheimer’s Association, Accelerating Medicines Partnership, GHR Foundation, an anonymous foundation, and additional private donors, with in-kind support from Avid Radiopharmaceuticals, Cogstate, Albert Einstein College of Medicine and the Foundation for Neurologic Diseases. The companion observational Longitudinal Evaluation of Amyloid Risk and Neurodegeneration (LEARN) Study was funded by the Alzheimer’s Association and GHR Foundation. The A4 and LEARN Studies were led by Dr. Reisa Sperling at Brigham and Women’s Hospital, Harvard Medical School, and Dr. Paul Aisen at the Alzheimer’s Therapeutic Research Institute (ATRI) at the University of Southern California. The A4 and LEARN Studies were coordinated by ATRI at the University of Southern California, and the data are made available under the auspices of the Alzheimer’s Clinical Trial Consortium through the Global Research & Imaging Platform (GRIP). The complete A4 Study Team list is available at: https://www.actcinfo.org/a4-study-team-lists/. We want to acknowledge the dedication of the study participants and their partners who made the A4 and LEARN Studies possible. We want to acknowledge the participants and investigators of the FinnGen study and the PGC consortium, and we thank FinnGen (https://www.finngen.fi/en) and PGC (https://pgc.unc.edu/) for their generosity in sharing the GWAS summary statistics with the scientific community.

## Inclusion & Ethics Statement

Our analyses considered both inclusion and ethnic diversity. All analyses included female and male participants, with models run jointly and/or separately by sex. Primary GWASs were conducted in European populations; however, we also examined the generalizability of these signals in non-European populations. Additionally, different disease groups were analyzed.

## Extended Data Figure

**Extended Data Fig. 1:**
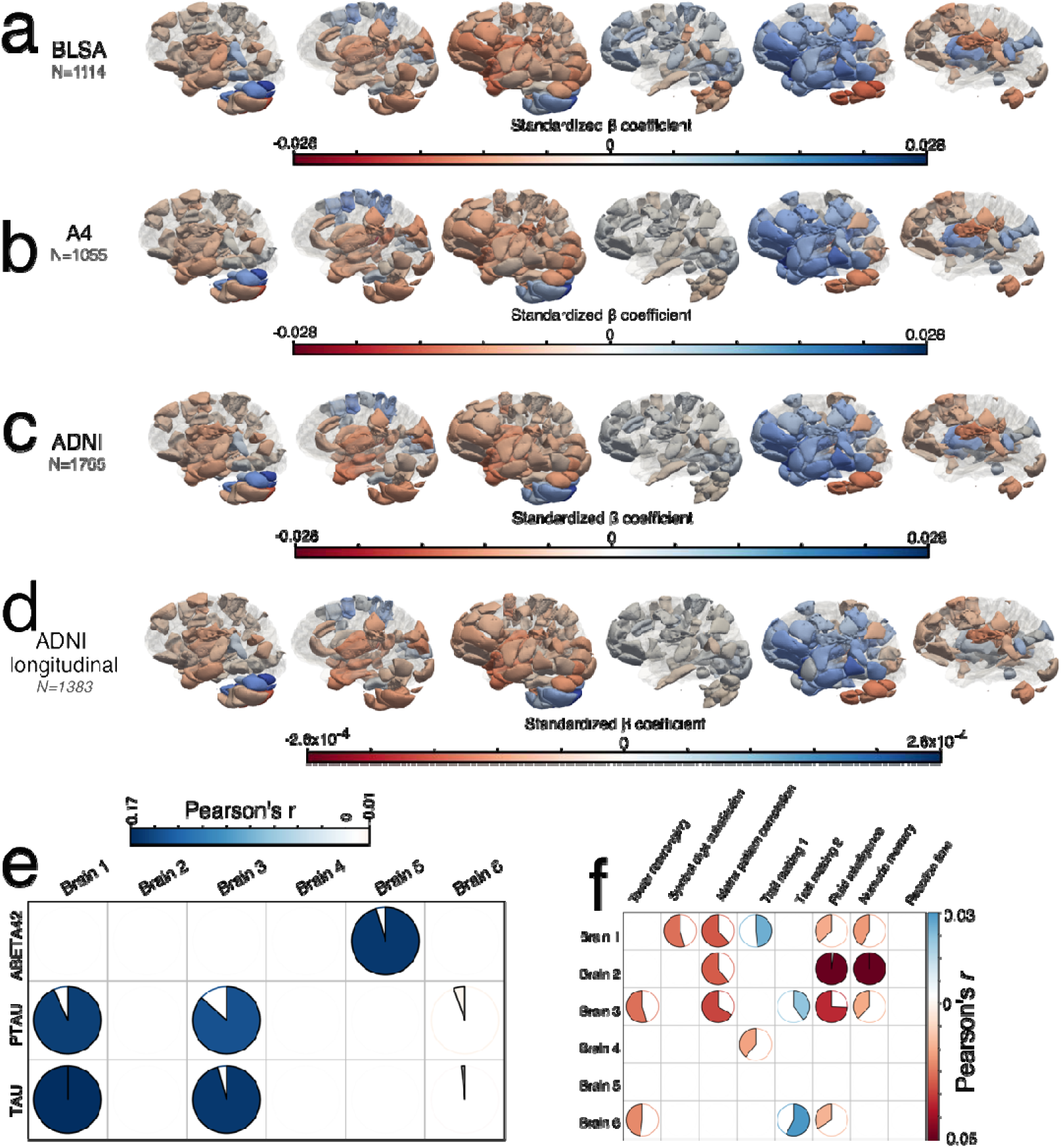
Manifestation of the 6 brain MAEs in independent, harmonized datasets covering the AD continuum and their longitudinal follow-ups. **a**) The imaging patterns of the 6 brain MAEs manifest in an aging cohort (BLSA) using the baseline scans. The same brain regions that passed the Bonferroni correction in the training data are presented with the same effect size magnitude (−0.028<standardized *β* coefficient<0.028). **b**) The imaging patterns of the 6 brain MAEs manifest in a preclinical AD cohort (A4) using the baseline scans. The same brain regions that passed the Bonferroni correction in the training data are presented with the same effect size magnitude (−0.028<standardized *β* coefficient<0.028). **c**) The imaging patterns of the 6 brain MAEs manifest in an AD clinical cohort (ADNI) using the baseline scans. The same brain regions that passed the Bonferroni correction in the training data are presented with the same effect size magnitude (−0.028<standardized *β* coefficient<0.028). **d**) The imaging patterns of the 6 brain MAEs manifest longitudinally in the ADNI data (*N*=1383 at baseline for at least 5 follow-up MRI scans). **e**) The ADNI data shows the associations between the 6 brain MAEs and CSF levels of Aβ_1-42_, tau_181p_, and p-tau_181p_. Statistical significance was assessed using a linear regression model that adjusted for covariates, with the Bonferroni correction applied. Pearson’s *r* was reported for the significant associations. **f**) The associations between the 6 brain MAEs and 8 cognitive scores in the UKBB data. Statistical significance was assessed using a linear regression model that adjusted for covariates, with the Bonferroni correction applied. Pearson’s *r* was reported for the significant associations. The scatter plot shows a linear regression line for visualization despite a weak linear effect.

**Extended Data Fig. 2:**
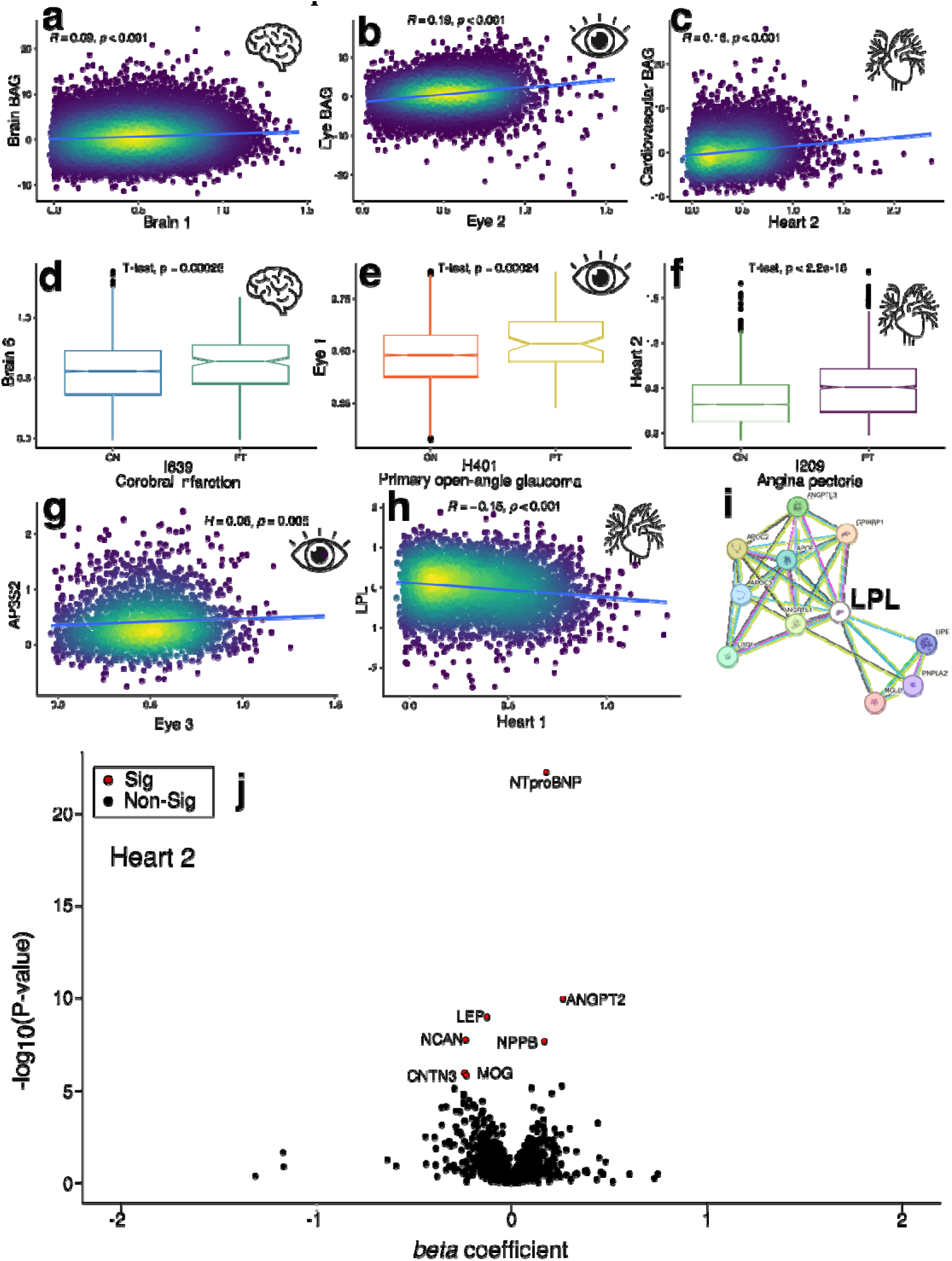
Representative association results for our PWAS and ProWAS across the brain, eye, and heart pan-disease. **a**) The scatter plot between Brain 1 and brain BAG. **b**) The scatter plot between Eye 2 and eye BAG. **c**) The scatter plot between Heart 2 and cardiovascular BAG. The brightness of the dots reflects their density, and the plot includes Pearson’s correlation coefficient (*R*) and the corresponding P-value, with a fitted linear regression line overlaid. Full sets of covariate-corrected statistics from a linear regression model are reported in the **Supplementary Material**. **d**) The box plot for Brain 6 between healthy controls (CN) and patients (PT) of cerebral infarction (ICD-10 code: I639). **e**) The box plot for Eye 1 between healthy controls (CN) and patients (PT) of primary open-angle glaucoma (ICD-10 code: H401). **f**) The box plot for Heart 2 between healthy controls (CN) and patients (PT) of angina pectoris (ICD-10 code: I209). The mean value of the MAE and the P-value of the two-sample t-test are shown, along with outlier data points. **g**) The scatter plot between Eye 3 and the AP3S2 protein. **h**) The scatter plot between Heart 1 and the LPL protein. **i**) The protein-protein-interaction (PPI) network for the LPL protein is presented using the STRING database. **j**) The volcano plots for Heart 2 in our ProWAS analyses. Significant signals are annotated by red dots and protein symbols.

**Extended Data Fig. 3:**
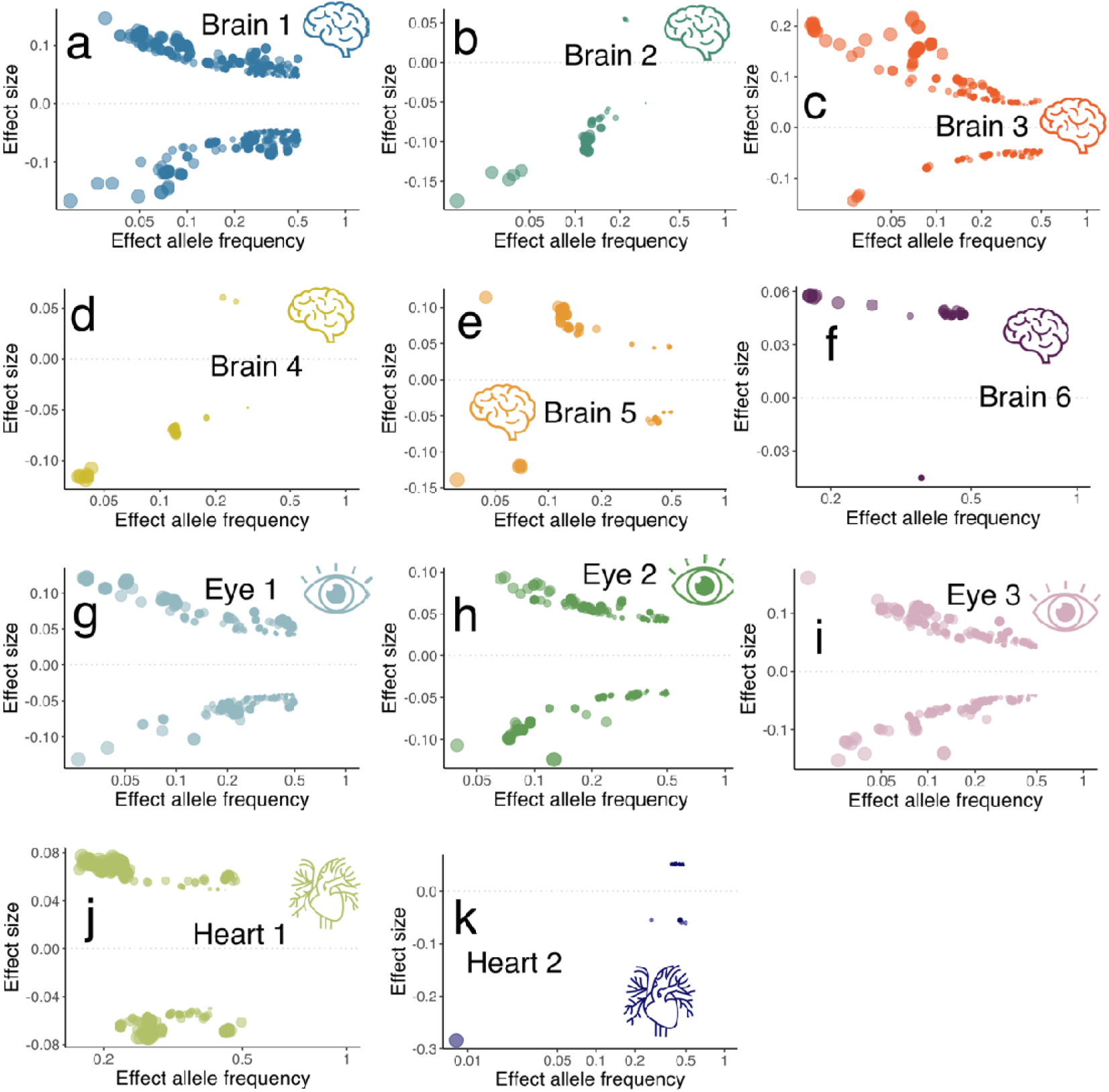
Trumpet plots of the effect (alternative) allele frequency vs. the *β* coefficient of the 11 MAE GWASs. The trumpet plots display the inverse relationship between the alternative (effect) allele frequency and the effect size (*β* coefficient) for the 11 MAEs. For visualization purposes, we showed the SNPs that passed the genome-wide P-value threshold (LD was considered in all other analyses). The dot size corresponds to the effect size, while the transparency of the dot is proportional to its statistical significance.

**Extended Data Fig. 4:**
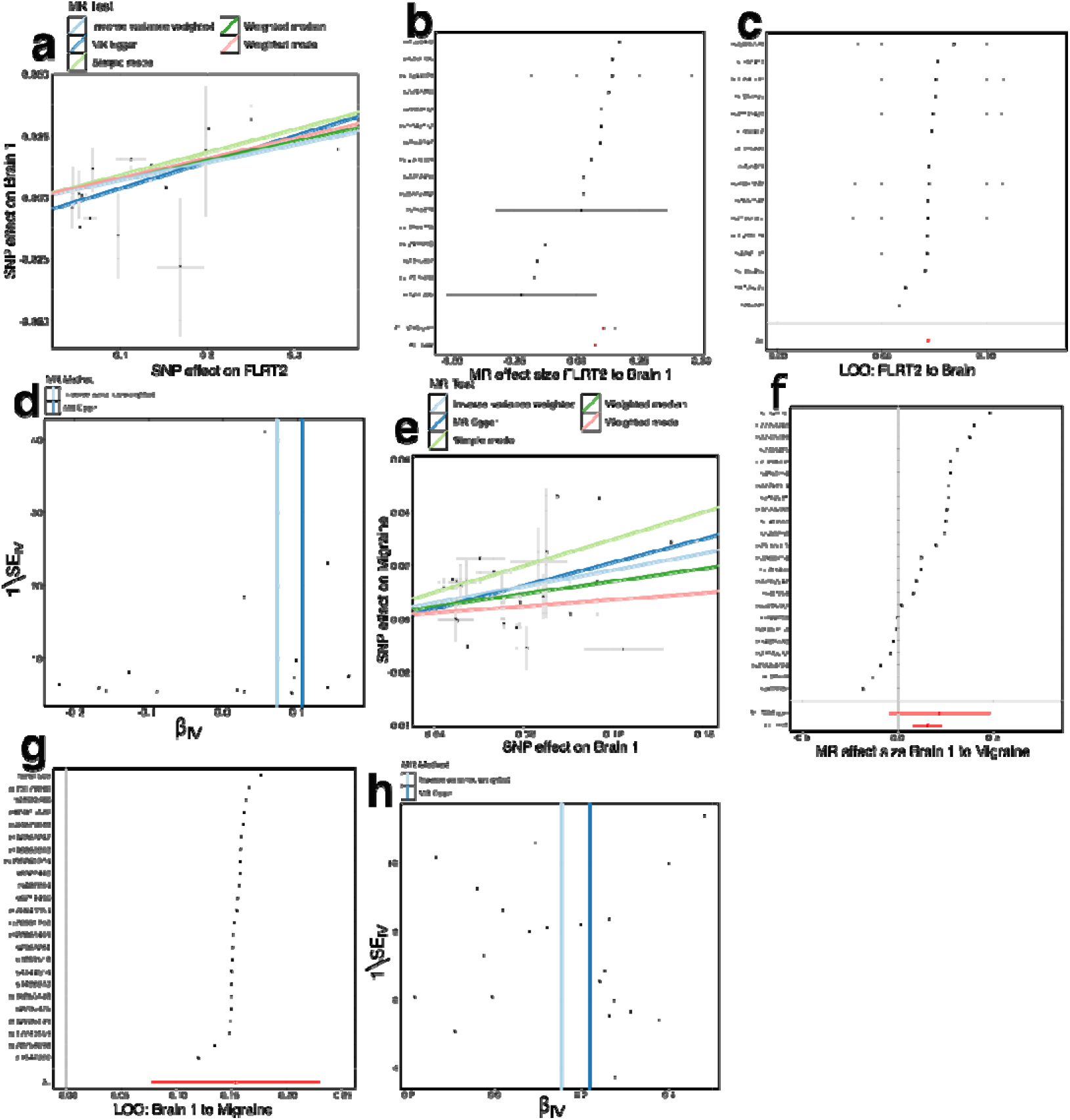
Sensitivity check analyses for the causal pathway of “FLRT2 → Brain 1 → migraine”. **a**) Scatter plot for the MR effect sizes of the SNP-FLRT2 association (*x*-axis, log OR) and the SNP-Brain 1 associations (*y*-axis, log OR) with standard error bars. The slopes of the five lines correspond to the causal effect sizes estimated by the five MR estimators, respectively. **b**) Forest plot for the single-SNP MR results. Each dot represents the MR effect (log (OR)), and the error bar displays the 95% CI for FLRT2 on Brain 1 using only one SNP; the red line shows the MR effect using all SNPs together for IVW and MR Egger estimators. **c**) Leave-one-SNP-out analysis of FLRT2 on Brain 1. Each dot represents the MR effect (log OR), and the error bar displays the 95% CI by excluding that SNP from the analysis. The red line depicts the IVW estimator using all SNPs. **d**) Funnel plot for the relationship between the causal effect of FLRT2 on Brain 1. Each dot represents MR effect sizes estimated using each SNP as a separate instrument against the inverse of the standard error of the causal estimate. **e**) Scatter plot for the MR effect sizes of the SNP-Brain 1 association (*x*-axis, log OR) and the SNP-migraine associations (*y*-axis, SD units) with standard error bars. The slopes of the five lines correspond to the causal effect sizes estimated by the five MR estimators, respectively. **f**) Forest plot for the single-SNP MR results. Each dot represents the MR effect (log OR), and the error bar displays the 95% CI for Brain 1 on migraine using only one SNP; the red line shows the MR effect using all SNPs together for IVW and MR Egger estimators. **g**) Leave-one-SNP-out analysis of Brain 1 on migraine. Each dot represents the MR effect (log OR), and the error bar displays the 95% CI by excluding that SNP from the analysis. The red line depicts the IVW estimator using all SNPs. **h**) Funnel plot for the relationship between the causal effect of Brain 1 on migraine. Each dot represents MR effect sizes estimated using each SNP as a separate instrument against the inverse of the standard error of the causal estimate.

**Extended Data Fig. 5:**
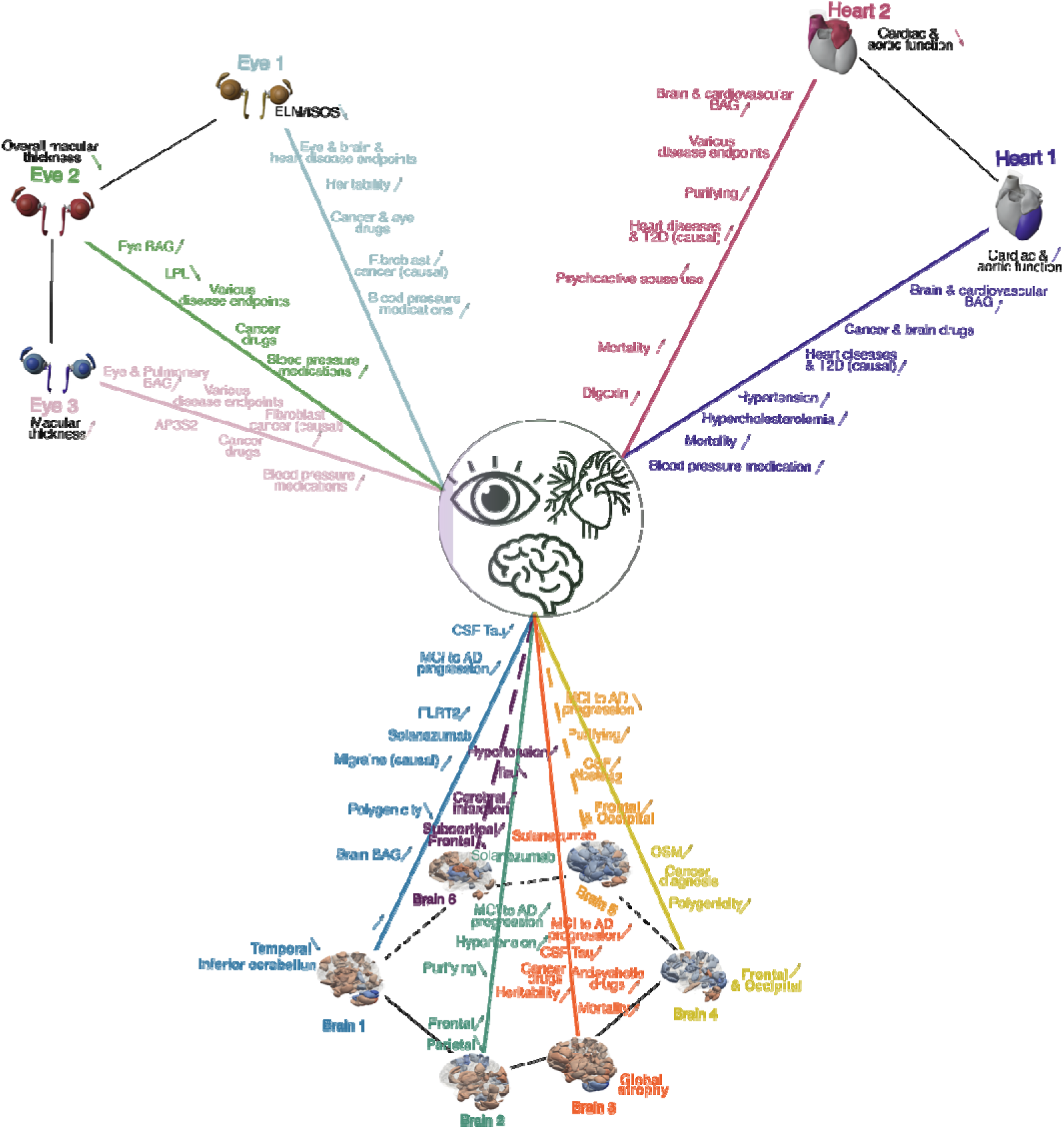
A dimensional representation of the 11 Multi-organ AI Endophenotypes (MAEs) for pan-disease of the brain, eye, and heart. We summarized the key characteristics of the 11 MAEs to understand the heterogeneity and commonalities across pan-disease manifestations in the brain, eye, and heart. Each MAE is represented by a distinct color, while the black-italicized text describes the imaging patterns specific to each MAE. Directional arrows indicate trends in characteristics such as atrophy/enlarged volume or thinning/thickening. If no directional trend was observed (e.g., in gene-drug-disease networks), arrows were omitted from the illustration.

## The MULTI consortium

Junhao Wen^1,8,9,110,11,12^*, Christos Davatziko^2^, Ye Ella Tian^3^, Wenjia Bai^4^, Michael S. Rafii^5^, Paul Aisen^5^, Keenan A. Walker^6^, Andrew Zalesky^3^, Luigi Ferrucci^6^, Jian Zeng^7^

^1^Laboratory of AI and Biomedical Science (LABS), Columbia University, New York, NY, USA

^2^Artificial Intelligence in Biomedical Imaging Laboratory (AIBIL), Center for AI and Data Science for Integrated Diagnostics (AI^2^D), Perelman School of Medicine, University of Pennsylvania, Philadelphia, PA, USA

^3^Systems Lab, Department of Psychiatry, Melbourne Medical School, The University of Melbourne, Melbourne, Victoria, Australia

^4^Department of Brain Sciences and Department of Computing, Imperial College London, London, UK

^5^Alzheimer’s Therapeutic Research Institute, Keck School of Medicine of the University of Southern California, San Diego, CA 92121, USA

^6^National Institute on Aging, National Institutes of Health, Baltimore, MD, USA

^7^The Institute for Molecular Bioscience, University of Queensland, Brisbane, QLD 4072, Australia

^8^Department of Radiology, Columbia University, New York, NY, USA

^9^Department of Biomedical Engineering, Columbia University, New York, NY, USA

^10^New York Genome Center (NYGC), New York, NY, USA

^11^Center for Innovation in Imaging Biomarkers and Integrated Diagnostics (CIMBID), Department of Radiology,

^12^Data Science Institute (DSI), Columbia University, New York, NY, USA Columbia University, New York, NY, USA

