## supplement for "Multi-organ AI Endophenotypes Chart the Heterogeneity of Pan-disease in the Brain, Eye, and Heart"

### Online Supplementary Materials

**eMethod 1: The definition of genomic loci, independent significant SNP, lead SNP, candidate SNP**
**eMethod 2: Structural covariance patterns via stochastic orthogonally projective non-negative matrix factorization**
**eMethod 3: Methodological advances in Surreal-GAN from our previous work**
**eNote 1: Strategies to combat potential domain shifts in our brain MAEs generalization across studies, populations, and age ranges**
**eNote 2: Secondary PWAS to link the 11 MAEs with clinical traits beyond the imaging-derived phenotypes of the brain, eye, and heart in UKBB**
**eNote 3: ProWAS results for the brain MAE using UKBB Olink data and its comparison with BLSA SomaScan data**
**eNote 4: Sensitivity check analyses for the primary GWAS of the 11 MAEs**
**eNote 5: PheWAS for the top lead SNP of the genomic loci linked to the 11 MAEs using the GWAS Atlas platform**
**eNote 6: Sensitivity check analyses for the Mendelian randomization analysis results**
**eNote 7: Comparisons of cis- and trans-pQTLs identified by the current study and from Sun et al. using the UKBB data for the FLRT2 protein**
**eFigure 1: The expression of the 11 MAEs in the UKBB test dataset**
**eFigure 2: The expression of the 6 MAEs in the UKBB test dataset in the image space**
**eFigure 3: Associations between 11 MAEs in UKBB and clinical features from other organs**
**eFigure 4: RNA and protein expression levels of the AP3S2 protein in different tissues using the HPA, GTEx data**
**eFigure 5: PheWAS results of the top lead SNPs in the GWAS Atlas platform**
**eFigure 6a: Manhattan and QQ plots for the brain MAE GWASs**
**eFigure 6b: Manhattan and QQ plots for the eye and heart MAE GWASs**
**eFigure 7: The MAPT protein's cancer-type specificity and single-cell specific expression patterns in the brain and breast tissues**
**eFigure 8: Potential conceptualized causal pathways linking genetics, proteomics, MAEs, and DEs**
**eFigure 9: The pQTL identified in the current study vs. these by Sun et al. for the FLRT2 protein**
**eFigure 10: The FLRT2 protein's RNA and single-cell expression profiles**
**eFigure 11: Group differences for the medication status of Digoxin for Heart 2**
**eFigure 12: Comparisons between the ProWAS results between UKBB Olink and BLSA SomaScan proteins with Brain 4**
**eFigure 13: Beta coefficients of the significant SNPs of the GWAS for Brain 1 in UKBB vs. ADNI**
**eFigure 14: MAE differences in different ethnic groups compared to European**
**eFigure 15: Model convergence of the 3 Surreal-GAN models based on the R-indices correlation along the training epochs**
**eTable 1: The characteristics of the MULTI consortium**
**eTable 2: The associations between the 6 brain MAEs and CSF and plasma neuropathological biomarkers using ADNI and UKBB**
**eTable 3: The associations between the 6 brain MAEs and 8 cognitive scores using UKBB**

**eTable 4: The associations between the 11 MAEs and 9 BAGs using UKBB**
**eTable 5: The SNP-based heritability estimates from three methods**
**eTable 6: The polygenicity and nature selection signatures of the 11 MAEs**
**eTable 7: The phenotypic and genetic associations between the 11 MAEs**
**eTable 8: The genetic correlation between the 11 MAEs and the 9 BAGs**
**eTable 9: The results of survival analyses for predicting AD progression**
**eTable 10: The results of survival analyses for predicting mortality**
**eTable 11: Preclinical AD drug outcome (PACC) is linked to Brain 1-3**
**eTable 12: The brain PSCs, heart IDPs, and eye IDPs included to derive the 11 MAEs via Surreal-GAN**
**eFile 1-19: The Online Supplementary files contain large tables**
**eFolder 1: Sensitivity analyses for the MR analyses**
**GWAS summary statistics**

**eMethod 1: The definition of genomic loci, independent significant SNP, lead SNP, candidate SNP**

FUMA defined the significant independent SNPs, lead SNPs, candidate SNPs, and genomic risk loci as follows (https://fuma.ctglab.nl/tutorial#snp2gene):

*Independent significant SNPs*

They are defined as SNPs with $P \leq 5\times10^{-8}$ that are independent of each other at the user-defined $r^2$ (set to 0.6 in the current study). We further describe *candidate SNPs* as those in linkage disequilibrium (LD) with independent significant SNPs. FUMA then queries each candidate SNP in the GWAS Catalog to check whether any clinical traits have been reported to be associated with previous GWAS studies.

*Lead SNPs*

Lead SNPs are defined as independent significant SNPs that are also independent of each other at $r^2<0.1$. If multiple independent significant SNPs are correlated at $r^2 \geq 0.1$, then the one with the lowest individual *P*-value becomes the lead SNP. If $r^2$ threshold is set to 0.1 for the independent significant SNPs, then they would constitute the identical set as the lead SNPs. FUMA thus advises setting $r^2$ to be 0.6 or higher.

*Genomic risk loci*

FUMA defines genomic risk loci to include all independent signals physically close or overlapping in a single locus. First, independent significant SNPs dependent on each other at $r^2 \geq 0.1$ are assigned to the same genomic risk locus. Then, independent significant SNPs with less than the user-defined distance (250 kilobases by default) away from one another are merged into the same genomic risk locus - the distance between two LD blocks of two independent significant SNPs is the distance between the closest points from each LD block. Each locus is represented by the SNP within the locus with the lowest *P*-value.

**eMethod 2: Structural covariance patterns via stochastic orthogonally projective non-negative matrix factorization**

The sopNMF algorithm is a stochastic approximation built and extended based on opNMF[1,2]. We consider a dataset of $n$ MR images and $d$ voxels per image. We represent the data as a matrix $\boldsymbol{X}$ where each column corresponds to a flattened image: $\boldsymbol{X} = [\boldsymbol{x}_1, \boldsymbol{x}_2, \dots, \boldsymbol{x}_n], \boldsymbol{X} \in \mathbb{R}_{\geq 0}^{d \times n}$. The sopNMF algorithm factorizes $\boldsymbol{X}$ into two low-rank ($r$) matrices $\boldsymbol{W} \in \mathbb{R}_{\geq 0}^{d \times r}$ and $\boldsymbol{H} \in \mathbb{R}_{\geq 0}^{r \times n}$ under the constraints of non-negativity and column-orthonormality. Using the Frobenius norm, the loss of this factorization problem can be formulated as

$$\min_{\boldsymbol{W},\boldsymbol{H}} \|\boldsymbol{X} - \boldsymbol{W}\boldsymbol{H}\|_F^2$$

$$\text{subject to } \boldsymbol{H} = \boldsymbol{W}^T\boldsymbol{X}, \boldsymbol{W} \geq 0 \text{ and } \boldsymbol{W}^T\boldsymbol{W} = \boldsymbol{I}, (1)$$

where $\boldsymbol{I}$ stands for the identity matrix. The columns $\boldsymbol{w}_i \in \mathbb{R}^d, \|\boldsymbol{w}_i\|_2 = 1, \forall i \in \{1..r\}$ of the so-called component matrix $\boldsymbol{W} = [\boldsymbol{w}_1, \boldsymbol{w}_2, \dots, \boldsymbol{w}_r]$ are part-based representations promoting sparsity in data in this lower-dimensional subspace. From this perspective, the loading coefficient matrix $\boldsymbol{H}$ represents the importance (weights) of each feature above for a given image. Instead of optimizing the non-convex problem in a batch learning paradigm (i.e., reading all images into memory) as opNMF,[1] sopNMF subsamples the number of images at each iteration, thereby significantly reducing its memory demand. This is done by randomly drawing data batches $\boldsymbol{X}_{B_i} = \boldsymbol{X}_{\cdot,B_i} \in \mathbb{R}_{\geq 0}^{d \times b}$ of $b \leq n$ images ($b$ is the batch size; $b$=32 was used in the current analyses). Here, $B_i \subset \{1..n\}$ denotes a random subset of $|B_i| \approx b$ indices. The sorting is done without replacement, $B_i \cap B_j = \emptyset$, so that all data goes through the model once ($i \in \{1..\lceil n/b \rceil\}$).

In our formulation, the internal updating rule can be rewritten as

$$\boldsymbol{W}_{t+1} = \boldsymbol{W}_t \frac{\boldsymbol{X}_{B_t}\boldsymbol{X}_{B_t}^T\boldsymbol{W}_t}{\boldsymbol{W}_t\boldsymbol{W}_t^T\boldsymbol{X}_{B_t}\boldsymbol{X}_{B_t}^T\boldsymbol{W}_t} \ (2)$$

After running through all the image data ($\lceil n/b \rceil$ such updates), we reach the end of the epoch and calculate the loss on the entire dataset (i.e., the loss is incremental across all batches) as

$$\theta = \sum_{i=1}^{\lceil n/b \rceil} \left\|\boldsymbol{X}_{B_i} - \boldsymbol{W}\boldsymbol{W}^T\boldsymbol{X}_{B_i}\right\|_F^2 \ (3)$$

where we call $\boldsymbol{W} = \boldsymbol{W}_{\lceil n/b \rceil}$ for simplicity.

We evaluated the training loss and the sparsity of $\boldsymbol{W}$ at the end of each epoch. Moreover, early stopping was implemented to improve training efficiency and alleviate overfitting. We summarize the sopNMF algorithm in **SI Algorithm 1**. An empirical comparison between sopNMF and opNMF is detailed in **SI eMethod 1**.

We applied sopNMF to the training population ($N$=4000). The component matrix $\boldsymbol{W}$ was sparse after the algorithm converged with a pre-defined maximum number of epochs (100 by default) with an early stopping criterion. To build the MuSIC atlas, we clustered each voxel (row-wise) into one of the $r$ features/PSCs as per

$$\boldsymbol{m}_j = \text{argmax}_{k \in \{1..r\}}(\boldsymbol{W}_{j,k}) \ (4)$$

where $\boldsymbol{m} \in \mathbb{R}_{\geq 0}^d$ is a $d$-dimensional vector and $j \in \{1..d\}$. The $j$-th element of $\boldsymbol{m}$ equals $k$ if $\boldsymbol{W}_{j,k}$ is the maximum value of the $j$-th row. In this way, $\boldsymbol{m}$ assigns a PSC to each voxel. We finally projected the vector $\boldsymbol{m}$ back to the original image space to visualize each PSC of the MuSIC atlas (as shown in **Fig. 2a**). We present the algorithm below:

**Algorithm 1:** Algorithm for sopNMF.

The source code of the Python implementation of sopNMF is available here: https://github.com/anbai106/SOPNMF

---

**Algorithm 1:** sopNMF

---

**Data:** Image data $\mathbf{X} \in \mathbb{R}^{d \times n}$, batch size $b$, desired rank $r$, early-stopping criterium $\mathrm{stop}(\cdot, \cdot)$ based on loss evolution, maximum number of epochs $p$

**Result:** $\mathbf{W} \in \mathbb{R}^{d \times r}$, $\mathbf{H} \in \mathbb{R}^{r \times n}$

Initialize $\mathbf{W}_0 = \mathbf{1}$; $\theta_1 = \infty$; $\theta_0 = 0$;

**while** epoch $< p$ **and not** $\mathrm{stop}(\theta_1, \theta_0)$ **do**

  **for** $i \leftarrow 1$ **to** $\lceil n/b \rceil$ **by** $1$ **do**

    Update $\mathbf{W}_i$ from $\mathbf{W}_{i-1}$ via (2) using the image batch $\mathbf{X}_{B_i}$;

  **end**

  $\theta_0 \leftarrow \theta_1$;

  Compute the new loss $\theta_1$ via (3);

  Randomize the columns of $\mathbf{X}$;

  $\mathbf{W}_0 \leftarrow \mathbf{W}_{\lceil n/b \rceil}$;

  epoch$+ = 1$;

**end**

$\mathbf{W} \leftarrow \mathbf{W}_{\lceil n/b \rceil}$;

$\mathbf{H} \leftarrow \mathbf{W}^\intercal \mathbf{X}$;

---

**eMethod 3: Methodological advances in Surreal-GAN from our previous work**

Surreal-GAN is an advanced deep representation learning approach proposed for dissecting disease-related heterogeneity under the principle of weakly-supervised learning. The key innovation of Surreal-GAN lies in modeling phenotypic heterogeneity by considering both spatial and temporal (i.e., disease severity) variation using only baseline data. This enables the derivation of low-dimensional R-indices that directly reflect the severity of distinct phenotypic change patterns. To capture these changes due to disease effects, Surreal-GAN leverages the generative adversarial network (GAN) to learn multiple transformations from a reference (REF) group (e.g., CN) to a target (TAR) group (e.g., PT)). Specifically, the method learns a function f, which maps the REF data x to synthesized TAR data $\mathrm{y}' = f(\mathrm{x}, \mathrm{z})$, where z is a latent variable indicating the transformation directions. As typical in GAN frameworks, an adversarial discriminator *D* distinguishes between real TAR data **y** and synthesized TAR data $\mathrm{y}'$, ensuring the generated data are indistinguishable from real patient data.

Beyond that, an inverse mapping, g, is introduced to re-estimate the latent variables z from the generated data $\mathrm{y}'$, ensuring that the latent variables capture distinct and recognizable phenotypic signatures. Multiple other regularizations were employed to further guide the transformation function f to approximate the disease effect while promoting the positive association between different dimensions of z and the severity of changes of various features.

During the model inference stage, the inverse function, g, is utilized to derive the latent variables (referred to as R-indices) for real TAR data after the training process. Through GAN and regularizations during training, the learned transformation function f was considered a good approximation of the underlying pathological process, denoted by function h, such that $\mathrm{f}(\mathrm{x}, \mathrm{z}) \approx \mathrm{h}(\mathrm{x}, \sigma(\mathrm{z}))$, where $\sigma \in \Omega$ represents a class of permutation functions that reorder the elements in the latent variables z. Since the orders of indices in z are unimportant and can be rearranged for optimal matching, we simplify this to $\mathrm{f}(\mathrm{x}, \mathrm{z}) \approx \mathrm{h}(\mathrm{x}, \mathrm{z})$ without loss of generality. For any real TAR data, $\bar{\mathrm{y}} = \mathrm{h}(\bar{\mathrm{x}}, \mathrm{r}) \sim \mathrm{p}_{\mathrm{tar}}(\mathrm{y})$, we estimate its ground truth R-indices, denoted as r, through $\mathrm{g}(\bar{\mathrm{y}}) = \mathrm{g}\big(\mathrm{h}(\bar{\mathrm{x}}, \mathrm{r})\big) \approx \mathrm{g}\big(\mathrm{f}(\bar{\mathrm{x}}, \mathrm{r})\big) \approx \mathrm{r}$. More methodological details can be found in the initial paper by Yang et al[3].

Recently, the Surreal-GAN model has been improved to capture correlations in its latent space, enabling the derivation of correlated R-indices that reflect associated underlying pathologies[4]. In contrast, the vanilla Surreal-GAN models predominantly derive independent R-indices due to the fixed sampling distribution (i.e., a standard multivariate uniform distribution) for latent variables during training. Specifically, under the assumption of inverse consistency, $\mathrm{g}\big(\mathrm{f}(\mathrm{x}, \mathrm{z})\big) \approx \mathrm{z}$, and equality in distributions, $\mathrm{p}_{\mathrm{syn}}(\mathrm{f}(\mathrm{x}, \mathrm{z})) \approx \mathrm{p}_{\mathrm{tar}}(\mathrm{y})$, the distribution of R-indices for TAR participants satisfies $\mathrm{p}(\mathrm{g}(\mathrm{y})) \approx \mathrm{p}(\mathrm{g}(\mathrm{f}(\mathrm{x}, \mathrm{z}))) \approx \mathrm{p}(\mathrm{z})$. As a result, sampling z from a standard multivariate uniform distribution, as done in the vanilla Surreal-GAN, leads to a covariance of derived R-indices that equals the identity matrix, which can introduce bias and reduce model performances when the ground-truth R-indices are correlated.

The improved version constructed a parametrized latent distribution for z using gaussian copula, denoted as $\mathrm{p}_{\theta_\mathrm{z}}(\mathrm{z}) = \mathrm{C}_{\theta_\mathrm{z}}^{\mathrm{Gauss}}(\mathrm{z})$, where the learnable parameters $\theta_\mathrm{z}$ control the correlations among dimensions:

$$\mathrm{p}_{\theta_\mathrm{z}}(\mathrm{z}) = \mathrm{C}_{\theta_\mathrm{z}}^{\mathrm{Gauss}}(\mathrm{z}) = \Phi_{\theta_\mathrm{z}}(\Phi^{-1}(\mathrm{z}_1), \Phi^{-1}(\mathrm{z}_2), \ldots, \Phi^{-1}(\mathrm{z}_\mathrm{M})) \quad (1)$$

where $\Phi^{-1}$ is the inverse cumulative distribution function of a standard normal and $\Phi_{\theta_\mathrm{z}}$ is the joint cumulative distribution function of a multivariate normal distribution with the mean vector zero and the covariance matrix equal to a correlation matrix $\theta_\mathrm{z}$. Further modifications were made to the original GAN loss function to optimize this parameterized latent distribution for approximating the ground-truth correlations:

$$L_{GAN}(\theta_D, \theta_f, \theta_z) = E_{y \sim p_{tar}(y)}\left[\log\big(D(y)\big)\right] + E_{y' \sim p_{syn}(y')}\left[1 - \log\big(D(y')\big)\right] \quad (2)$$

$$= E_{z \sim p_{\theta_z}(z), y \sim p_{tar}(y)}\left[\log\big(D(y)\big)\right] + E_{z \sim p_{\theta_z}(z), x \sim p_{ref}(x)}\left[1 - \log\Big(D\big(f(x, z)\big)\Big)\right] \quad (3)$$

$$= \int_D p_{\theta_z}(z) E_{y \sim p_{tar}(y)}\left[\log\big(D(y)\big)\right] d^M z + \int_D p_{\theta_z}(z) E_{x \sim p_{ref}(x)}\left[1 - \log\Big(D\big(f(x, z)\big)\Big)\right] d^M z \quad (4)$$

$$= \int_D p_U(z) p_{\theta_z}(z) E_{y \sim p_{tar}(y)}\left[\log\big(D(y)\big)\right] d^M z + \int_D p_U(z)\, p_{\theta_z}(z) E_{x \sim p_{ref}(x)}\left[1 - \log\Big(D\big(f(x, z)\big)\Big)\right] d^M z \quad (5)$$

$$= E_{y \sim p_{tar}(y), z \sim p_U(z)}\left[p_{\theta_z}(z) \log\big(D(y)\big)\right] + E_{z \sim p_U(z), x \sim p_{ref}(x)}\left[p_{\theta_z}(z)\left(1 - \log\Big(D\big(f(x, z)\big)\Big)\right)\right] \quad (6)$$

The revised loss function allows sampling z from a uniform distribution, $p_U(z)$, while penalizing the losses with their probability under the distribution $p_{\theta_z}(z)$. Additionally, to prevent $p_{\theta_z}$ from converging to an extreme distribution (e.g., two latent variables become completely correlated), a regularization term is introduced to control the divergence between $p_{\theta_z}(z)$ and $p_U(z)$. As a result, the final modified GAN loss function equals:

$$L_{GAN}(\theta_D, \theta_f, \theta_{z_2}) = E_{y \sim p_{tar}(y), z \sim p_U(z)}\left[p_{\theta_z}(z) \log\big(D(y)\big)\right] + E_{z \sim p_U(z), x \sim p_{ref}(x)}\left[p_{\theta_z}(z)\left(1 - \log\Big(D\big(f(x, z)\big)\Big)\right)\right] + \alpha D_{KL}\left(p_U(z) \middle| p_{\theta_z}(z)\right) \quad (7)$$

This improvement enables the identification of associated R-indices, significantly boosting the model's performance and enhancing the method's applicability.

**eNote 1: Strategies to combat potential domain shifts in our brain MAEs generalization across studies, populations, and age ranges**

We carefully employed strategies for the 6 brain MAEs to mitigate potential domain shifts when applying the trained Surreal-GAN model on UKBB data to other datasets, including the BLSA, A4, and ADNI. Domain shifts are a common issue in ML/AI, especially when integrating data from multiple sources, and we accounted for this challenge in our approach to ensure consistent performance across studies.

We have thoroughly considered this while developing and applying the models to the UKBB data. First, the iSTAGING consortium systematically and statistically consolidated and harmonized the brain MRI data for more than 500k individuals worldwide, with different types of scanners, demographics, ethnicities, etc. We developed the Combat-GAM model to alleviate adverse effects induced by the site-/study-specific effect. Our previous study[5] showed that the harmonized imaging data, using our developed statistical harmonization method, showed improved data normality. Secondly, the Surreal-GAN model is a weakly-(semi-) supervised representation learning method, which leverages the healthy control (CN) population as a reference domain to model distinct dimensions ("*1-to-k*" mapping). Therefore, when we applied the pre-trained model to non-UKBB data, the data were corrected (linearly) and contrasted with the healthy control populations. Finally, empirical results showed that the expression of these imaging patterns of the 6 brain MAEs could be manifested in different studies (general populations vs. disease-specific clinical cohorts), age ranges (young vs. old), and longitudinal settings, albeit these cannot entirely exclude potential domain shifts. Therefore, caution must be taken when interpreting the results presented in **Extended Data Fig. 1**.

**eNote 2: Secondary PWAS to link the 11 MAEs with clinical traits beyond the imaging-derived phenotypes of the brain, eye, and heart in UKBB**

We conducted a secondary PWAS to link the 11 MAEs with clinical traits beyond the IDPs from the three organs. Between the 106 traits and 11 MAEs, we identified 190 significant MAE-trait associations (P-value<0.05/106/11) (**Supplementary eFigure 3**).

We have observed cross-organ interactions as we have explicitly excluded the imaging-derived phenotypes from the three organs. Most associations linked mental traits and the MAEs (90 trait-MAE pairs), such as between "ever manic/hyper for 2 days (Field ID=4642)" and Eye 1 ($\beta$=0.006±0.0009; P-value=1.70x$10^{-10}$). Lifestyle and environment-related traits were linked to MAEs from all three organs (15 trait-MAE pairs), including Brain 3 and "nap taken during the day (Field ID=1190)" ($\beta$=-0.038±0.009; P-value=2.60x$10^{-5}$), Eye 2 and "smoking status (Field ID=20116)" ($\beta$=0.036±0.007; P-value=8.63x$10^{-7}$), and Heart 2 and "alcohol drinker status (Field ID=20117)" ($\beta$=-0.01±0.001; P-value=4.00x$10^{-31}$). In addition, immune traits were also linked with multiple MAEs (19 trait-MAE pairs), including Brain 1 and "neutrophil count (Field ID=30140)" ($\beta$=0.017±0.004; P-value=3.20x$10^{-5}$), Eye 1 and "mean corpuscular volume (Filed ID=30040)" ($\beta$=0.006±0.001; P-value=2.61x$10^{-6}$), and Heart 2 and "hematocrit percentage (Field ID=30030)" ($\beta$=-0.011±0.002; P-value=1.13x$10^{-8}$). Finally, musculoskeletal traits were also largely linked to multiple MAEs (28 trait-MAE pairs), including Eye 1 and "waist circumference (Field ID=48)" ($\beta$=0.005±0.0004; P-value=1.74x$10^{-34}$). The detailed statistical results are presented in **Supplementary eFile 6**.

**eNote 3: ProWAS results for the brain MAE using UKBB Olink data and its comparison with BLSA SomaScan data**

We systematically compared the ProWAS signals using the UKBB Olink and BLSA SomaScan proteins. First, we ensured that the imaging patterns of the 6 MAEs manifested in the BLSA sample (**Extended Data Fig. 1a**). This comparative analysis yielded three main observations.

First, BLSA SomaScan proteins were more sensitive and linked with the brain MAEs, albeit the BLSA SomaScan sample size (*N*=924) was much smaller than that of the UKBB Olink proteins (*N*~4000). With a P-value threshold of 0.05/2139 common proteins presented in both platforms, we observed 60 MAE-protein significant signals for SomaScan. In contrast, none survived the multiple comparisons in the UKBB Olink data. In addition, the $\beta$ coefficient estimates of the two platforms showed a moderate correlation (Pearson's $r$ =0.26; P-value=0.026; **eFigure 12**). This was consistent with previous literature, where Eldjarn et al.[6] found the median Spearman correlation between plasma levels of 1,848 proteins measured with matching Olink and SomaScan assays was 0.33 in the Icelandic set of 1,514 individuals. In addition, they observed a higher coefficient of variation (CV) in Olink than in SomaScan. In proteomics analyses, CV assesses the variability in protein measurements across samples. It helps evaluate the consistency and reliability of the data, especially when comparing different platforms like SomaScan and Olink. A lower CV indicates higher precision and less variation between repeated measures of the same protein, which is crucial when determining associations between proteins and diseases. Furthermore, SomaScan is often noted for its broader proteomic coverage and higher sensitivity, which can lead to more significant associations with disease phenotypes, even with smaller sample sizes. This may result in lower CVs for SomaScan proteins compared to Olink, making the former more reliable for detecting subtle biological differences. Olink, while more targeted, generally has a higher CV due to its narrower panel of proteins, which could explain why it shows fewer significant associations than SomaScan.

Secondly, we observed that the $\beta$ coefficients showed opposite directions for certain proteins. As annotated in **eFigure 12**, for example, the CPLX2 protein was positively associated with Brain 4 in SomaScan ($\beta$=0.20±0.05; P-value=2x10$^{-5}$; *N*=924), whereas a negative association was found in Olink ($\beta$=-0.32; *N*=45). However, the sample size for the Olink data was very small, so the model could not converge to estimate the $\beta$ SE and P-value. Another example is the KLKB1-Brain 4 associations: Olink data showed one positive association ($\beta$=0.04±0.07; P-value=0.61; *N*=2928), and SomaScan showed a negative association ($\beta$=-0.37±0.08; P-value=1x10$^{-5}$; *N*=924), although the Olink association was not significant, and the $\beta$ SE was larger than the $\beta$ itself. Overall, this discrepancy may be attributed to several factors. First, SomaScan and Olink rely on different technologies to detect proteins. SomaScan uses aptamer-based binding, while Olink utilizes proximity extension assays. These methodological differences can result in variability in how the proteins are quantified and the strength or direction of associations with disease endpoints, such as brain MAEs. Secondly, SomaScan generally covers a broader range of proteins and is known for its sensitivity, which may influence the beta coefficients in different contexts. If a protein is detected with greater sensitivity on one platform, the observed associations might reflect a stronger or weaker effect, depending on sample characteristics or disease stage. Of note, our $\beta$ coefficient correlation was computed between two different populations with clear demographic discrepancies (i.e., UKBB vs. BLSA). Thirdly, some proteins may have context-specific roles in different diseases or phenotypes, and this can be captured differently by the platforms. For instance, one platform may better detect a protein's role in inflammation, while another captures its role in neurodegeneration, leading to conflicting results. Finally, biological complexity and technical artifacts can also contribute to these differences. Further validation is needed to discern whether these opposite beta coefficients are due to true biological differences or platform-specific variation.

Thirdly, we observed that high consistency was obtained in SomaScan for the MAE-protein associations using different SeqIDs. For example, in **eFigure 12**, we found a robust negative association between the IGFRB2 protein and Brain 4 using four different SeqIDs, including 8469-41, 2570-72, 8819-3, and 22985-160 ($0.17>\beta>0.14$; P-value$<1x10^{-5}$). In the SomaScan platform, certain proteins may be quantified under different SeqIDs, each representing distinct aptamers or probes specific to that protein. Each SeqID might, therefore, capture a different aspect of the protein's structure, post-translational modifications, or interactome, which can lead to variations in quantitative results. Using multiple SeqIDs for the same protein can enhance sensitivity and specificity, but it also necessitates careful interpretation to ensure that the measured associations accurately represent the biological effects rather than assay-specific variability. Cross-validation with alternative platforms like Olink or independent biomarkers can help verify the clinical relevance of these results.

In summary, our systematic comparisons show that SomaScan is often more powerful in detecting associations due to its broader coverage and sensitivity. However, Olink's results are usually more specific and may provide more actionable insights for targeted research, depending on the clinical or biological focus. Therefore, the platform choice should align with the study's goals, whether broad discovery (SomaScan) or targeted hypothesis testing (Olink).

**eNote 4: Sensitivity check analyses for the primary GWAS of the 11 MAEs**

We conducted four sensitivity checks to examine the reliability of our primary GWAS outcomes thoroughly – these assessments aimed to gauge the consistency of genetic signals across various conditions. For instance, in the split-sample GWAS, where both splits were randomly generated to maintain balanced age and sex distributions, split1 functioned as the discovery GWAS, while split2 was the replication GWAS. We identified *N* SNPs in split1 GWAS that surpassed the genome-wide significance threshold (P-value<5x10$^{-8}$). Subsequently, we assessed $N_n$ SNPs that exceeded the nominal P-value threshold (0.05) and $N_b$ SNPs that surpassed the Bonferroni-corrected P-value threshold (0.05/*N*). Regarding $\beta$ values, we evaluated the number of SNPs ($N_{beta}$), where the $\beta$ values aligned with those derived from the *N* SNPs in split1 GWAS.

- The concordance rate of the P-value is defined as:
  - At the nominal threshold: CR-$P_n$=$N_n/N$
  - At the Boneforroni corrected threshold: CR-$P_b$=$N_b/N$
- The concordance rate of the $\beta$ values:
  - At the nominal threshold: CR-$\beta$=$N_{beta}/N_n$
  - *r-$\beta$*=Pearson's *r* of the *$\beta$-discovery* and *$\beta$-replication SNPs from the $N_n$ SNPs.*

**Brain MAE GWAS:**

**Genomic inflation**

We estimated the LDSC intercept for the 6 brain MAEs: 1.0117 [1.0062, 1.0185]. Genomic inflation was negligible in our primary GWAS, with intercepts approaching 1.

**Split-sample GWAS**

*P-values:*

In the split1 GWAS, we found 175, 1, 128, 0, 12, and 0 MAE-SNP associations for Brain 1-6 (P-value < 5x10$^{-8}$). In the split2 GWAS, the concordance rates of the P-value were CR-$P_n$=0.97 and 0.98 for Brain 1 and Brain 3, using the nominal P-value and CR-$P_b$=0.94 and 0.55 for the Bonferroni-corrected threshold (<0.05/*N*).

*$\beta$ values:*

Compared to the $N_n$ SNPs from the split1 GWAS, the split2 GWAS showed a mean concordance rate of CR-$\beta$=1 and *r-$\beta$*=0.95. Detailed results of these statistics for each MAE are presented in **Supplementary eFile 16**.

**Sex-stratified GWAS**

*P-values:*

In the female GWAS, we found 175, 1, 128, 0, 12, and 0 MAE-SNP associations for Brain 1-6 (P-value < 5x10$^{-8}$). In the male GWAS, the mean concordance rate of the P-value was CR-$P_n$=0.99, 1, 0.99 for Brain 1-3, using the nominal P-value and CR-$P_b$=0.89, 0.88, 0.30 for the Bonferroni-corrected threshold (<0.05/*N*).

*$\beta$ values:*

Compared to the $N_n$ SNPs from the female GWAS, the male GWAS showed a mean concordance rate of CR-$\beta$=1 and *r-$\beta$*=0.99. Detailed results of these statistics for each MAE are presented in **Supplementary eFile 16**.

**Non-European GWAS**

*P-values:*
In the fastGWA European GWAS, we found 175, 1, 128, 0, 12, and 0 MAE -SNP associations for Brain 1, 2, 3, 5, and 6 (P-value < $5x10^{-8}$); The fastGWAS analysis for non-European populations for Brain 4 failed to converge due to the small sample size ($N$=4663). In the fastGWA non-European GWAS, the mean concordance rate of the P-value was CR-$P_n$=0.47 using the nominal P-value, indicating low replication across ethnicity groups based on P-values.

*$\beta$ values:*
Compared to the $N_n$ SNPs from the European GWAS, the non-European GWAS showed a mean concordance rate of CR-$\beta$=1. The $\beta$ values of the two sets were highly correlated ($r$-$\beta$=0.85). Detailed results of these statistics for each MAE are presented in **Supplementary eFile 16**. Based on the low replication for P-values and the high correlation between European and non-European ancestry $\beta$ values, our findings underscore the necessity of incorporating data from underrepresented ethnic groups in future studies to enhance generalizability and capture population-specific effects.

**ADNI WGS GWAS**
For the ADNI WGS data, we performed the quality check as below. We first convert the VCF files into *plink* binary format. We excluded related individuals (up to $2^{nd}$-degree) using the KING software for family relationship inference.[7] Further QC steps are: excluding criteria were: i) individuals with more than 2% of missing genotypes; ii) variants with minor allele frequency (MAF) of less than 0.1%; iii) variants with larger than 5% missing genotyping rate; iv) variants that failed the Hardy-Weinberg test at $1x10^{-5}$. We then removed duplicated variants from all 22 autosomal chromosomes. We also excluded individuals for whom either imaging or genetic data were not available. To adjust for population stratification,[8] we derived the first 40 genetic principal components (PC) using the SmartPCA software[9]. For the ADNI GWAS, due to the smaller sample size ($N$~1000), we fit a linear model using PLINK because fastGWA was designed for larger sample-sized GWAS to evaluate the genetic relationship matrix precisely.

*P-values:*
In the fastGWA UKBB GWAS, we 1111, 71, 730, 32, 104, and 43 MAE -SNP associations for Brain 1, 2, 3, 5, and 6 (P-value < $5x10^{-8}$); in the PLINK ADNI GWAS, the mean concordance rate of the P-value was CR-$P_n$<0.5 using the nominal P-value, indicating low replication using P-value threshold due to the limited sample sizes ($N$~1000).

*$\beta$ values:*
Effect sizes like $\beta$ value are independent of sample sizes. Compared to the $N_n$ SNPs from the UKBB GWAS, the ADNI GWAS showed a mean concordance rate of CR-$\beta$=0.93806, 0.46762, 0.97351, 0.99859, 0.74500 for Brain 1, 2, 3, 5, and 6; Brain 4 did not have any overlap SNPs between UKBB and ADNI for the significant signals. Detailed results of these statistics for each MAE are presented in **Supplementary eFile 16**.

In **eFigure 13**, we show the scatter plot for the $\beta$ values between the two datasets for Brain 1, which showed a high correlation between the two sets of $\beta$ values ($r$=0.94). The two sets of estimation showed concordant directions for most SNPs, with exceptions for the annotated three SNPs in high LD on chromosome 5. This emphasizes the importance of including more diverse genetic data, including disease-specific populations.

**Eye MAE GWAS:**

**Genomic inflation**

We estimated the LDSC intercept for the 3 eye MAEs: 1.0129 [1.0088, 1.015]. Genomic inflation was negligible in our primary GWAS, with intercepts approaching 1.

**Split-sample GWAS**

*P-values:*

In the split1 GWAS, we found 64, 30, and 336 MAE-SNP associations for the 3 eye MAEs (P-value < $5 \times 10^{-8}$). In the split2 GWAS, the mean concordance rate of the P-value was CR-$P_n$=0.99 using the nominal P-value and CR-$P_b$=0.79 for the Bonferroni-corrected threshold (<0.05/*N*).

*$\beta$ values:*

Compared to the $N_n$ SNPs from the split1 GWAS, the split2 GWAS showed a mean concordance rate of CR-$\beta$=1 and $r$-$\beta$=0.97. Detailed results of these statistics for each MAE are presented in **Supplementary eFile 17**.

**Sex-stratified GWAS**

*P-values:*

In the female GWAS, we found 128, 53, and 470 MAE-SNP associations for the 3eye MAEs (P-value < $5 \times 10^{-8}$). In the male GWAS, the mean concordance rate of the P-value was CR-$P_n$=0.96 using the nominal P-value and CR-$P_b$=0.75 for the Bonferroni-corrected threshold (<0.05/*N*).

*$\beta$ values:*

Compared to the $N_n$ SNPs from the female GWAS, the male GWAS showed a mean concordance rate of CR-$\beta$=1 and $r$-$\beta$=0.97. Detailed results of these statistics for each MAE are presented in **Supplementary eFile 17**.

**Non-European GWAS**

*P-values:*

In the fastGWA European GWAS, we found 1525, 1008, and 1590 MAE-SNP associations for the 3 eye MAEs (P-value < $5 \times 10^{-8}$). In the fastGWA non-European GWAS, the mean concordance rate of the P-value was CR-$P_n$=0.31 using the nominal P-value.

*$\beta$ values:*

Compared to the $N_n$ SNPs from the European GWAS, the non-European GWAS showed a mean concordance rate of CR-$\beta$=1. The $\beta$ values of the two sets were highly correlated ($r$-$\beta$=0.99). Detailed results of these statistics for each MAE are presented in **Supplementary eFile 17**.

**Heart MAE GWAS:**

**Genomic inflation**

We estimated the LDSC intercept for the 2 heart MAEs: 1.0059 and 1.0152. Genomic inflation was negligible in our primary GWAS, with intercepts approaching 1.

**Split-sample GWAS**

*P-values:*

In the split1 GWAS, we found 5 and 47 MAE-SNP associations for Heart 1 and Heart 2 (P-value < $5x10^{-8}$). In the split2 GWAS, the mean concordance rate of the P-value was CR-$P_n$=0.85 using the nominal P-value for Heart 2.

*$\beta$ values:*
Compared to the $N_n$ SNPs from the split1 GWAS, the split2 GWAS showed a mean concordance rate of CR-$\beta$=1 and $r$-$\beta$=0.99. Detailed results of these statistics for each MAE are presented in **Supplementary eFile 18**.

**Sex-stratified GWAS**
*P-values:*
In the female GWAS, we found 1 and 3 MAE-SNP associations for Heart 1 and Heart 2 (P-value < $5x10^{-8}$). In the male GWAS, the mean concordance rate of the P-value was CR-$P_n$=0 due to limited significant signals. Detailed results of these statistics for each MAE are presented in **Supplementary eFile 18**.

**Non-European GWAS**
*P-values:*
In the fastGWA European GWAS, we found 795 and 85 MAE-SNP associations for Heart 1 and Heart 2 (P-value < $5x10^{-8}$). In the fastGWA non-European GWAS, the concordance rate of the P-value was CR-$P_n$=0.73 using the nominal P-value for Heart 1.
*$\beta$ values:*
Compared to the $N_n$ SNPs from the European GWAS, the non-European GWAS showed a concordance rate of CR-$\beta$=1 for Heart 1. The $\beta$ values of the two sets were highly correlated ($r$-$\beta$=0.99) for Heart 1. Detailed results of these statistics are presented in **Supplementary eFile 18**.

**eNote 5: PheWAS for the top lead SNP of the genomic loci linked to the 11 MAEs using the GWAS Atlas platform**

We conducted a PheWAS look-up query to link the top lead SNPs of the 11 MAEs with clinical traits from previous GWASs in the GWAS Atlas platform. We found 1397 significant MAE-trait associations in prior GWASs between 11 MAEs and 434 traits (**Supplementary eFigure 5**). The 434 traits were further categorized into 22 high-level domains, including neurological traits, ophthalmological traits, etc.

Within-organ specificity has been largely observed. For example, the three eye MAEs were linked with various ophthalmological traits, such as Eye 3 vs. age-related macular degeneration (PMID: 26691988). Neurological traits were also largely linked with the top lead SNPs of the 6 brain MAEs, such as left cerebellum exterior vs. Brain 1 (PMID: 31676860), and right supramarginal vs. Brain 2 (PMID: 31676860). Cardiovascular traits were also linked to Heart 1 and Heart 2, such as atrial fibrillation vs. Heart 2 (PMID: 29892015). We also observed cross-organ interactions. For example, rs1333045 was associated with coronary artery disease (PMID: 26343387), which was the top lead SNP of Eye 1. The detailed statistical results are presented in **Supplementary eFile 19**.

Our PheWAS look-up analysis aligns with our secondary PWAS findings, as outlined in **eNote 2**. Unlike our secondary PWAS, we deliberately excluded traits related to imaging-derived phenotypes (IDPs) from the brain, eye, and heart – to avoid circular effects. Instead, the PheWAS incorporated significant SNPs identified in prior GWAS studies where these IDPs were used as the phenotypes of interest.

**eNote 6: Sensitivity check analyses for the Mendelian randomization analysis results**

As Mendelian randomization is sensitive to underlying IV assumptions (**Method 6i**), we performed sensitivity analyses to investigate the potential violation, exemplified by the two-layer causal pathway from the FLRT2 protein to brain1 and migraine disorders (**Fig. 4e**).

For the causal relationship from FLRT2 to Brain 1, we did not observe clear outlier instrumental variables (IV; i.e., independent SNPs) for the effect sizes on the exposure (i.e., FLRT2) and outcome variables (i.e., Brain 1) (**Extended Fig. 4 a**). We then showed the forest plot for the individual-SNP level of the causal effect sizes, indicating that most of the SNPs exert positive effects (**Extended Fig. 4 b**). We then performed a leave-one-IV-out analysis and found that no single SNPs largely dominated the causal effect (**Extended Fig. 4 c**). Finally, we showed potential asymmetry that may indicate potential "directional pleiotropy" (**Extended Fig. 4 d**). To further scrutinize this bias, we applied MR-Egger regression with MAF-corrected weights to the summarized data, yielding an intercept estimate of -0.00469 with an associated P-value of 0.13. The bias-adjusted causal effect estimate from MR-Egger regression is [P-value=$1.51 \times 10^{-3}$; OR (95% CI)= 1.11 (1.05, 1.17); number of IVs=16], a slight increase in magnitude and uncertainty compared with the IVW estimator. All results from the five different MR estimators are presented in **Supplementary eFile 12**. There was also no apparent heterogeneity in the IV estimates from each genetic variant individually, as evidenced by Cochran's Q test (P = 0.27). In summary, there is no evidence that directional pleiotropy is an important factor for these data.

Similarly, for the causal effect from Brain 1 to migraine disorders, we showed the scatter plot between the IV effect sizes from the exposure and outcome variables without clear outliers (**Extended Fig. 4 e**). Most SNPs exerted a positive causal effect in our individual-level IV analysis, as shown in the forest plot (**Extended Fig. 4 f**). No single SNP dominated the overall causal effect in the leave-one-IV-out analysis (**Extended Fig. 4 g**). Finally, we did not observe an asymmetry in the funnel plot (**Extended Fig. 4 h**). MR-Egger regression showed an intercept estimate of -0.00487 with a P-value of 0.62. This indicated that there was no evidence for potential horizontal pleiotropy. There was also no apparent heterogeneity in the IV estimates from each genetic variant, as evidenced by Cochran's Q test (P = 0.11). In summary, we did not observe clear violations that biased the causal relationship factor for these data.

**eNote 7: Comparisons of cis- and trans-pQTLs identified by the current study and from Sun et al. using the UKBB data for the FLRT2 protein**

Our ProGWAS was guided by a previous study by Sun et al. Here, we used the ProGWAS results of the FLRT2 protein to compare these results.

We downloaded the pQTL results of FLRT2 from Sun et al. from the official web portal (https://metabolomips.org/ukbbpgwas/pgwas.table.php). Of note, Sun et al. used and shared results based on the genome build version of GRCh38, whereas our results used the GRCh37 version. We employed the same criteria used by Sun et al. to define our *cis*- and *trans*-pQTLs, and the results are shown in **eFigure 9**. Of note, the same sizes (*N*), GWAS models, criteria to define the genomics loci, etc., may differ between the two studies. For example, our sample sizes were larger than Sun et al., as they used part of the UKBB data as the discovery set. Therefore, this resulted in a higher number of genomic loci (*N*=15 vs. *N*=11) identified by our studies. We tried to harmonize the procedures as much as possible, including using the same definition for the *trans*- and *cis*-pQTLs. A high concordance was observed between the two sets of analysis.

The top lead SNPs for the *cis*-pQTL between the two studies were in a high LD (rs2753616 vs. rs17646457; $R^2$=0.2644; Chi-sq=48.1139; P-value<0.0001), which evaluated the British population from the 1000 Genome reference data. Other examples for the *trans*-pQTLs in high LD included the results between rs12493830 and rs10935473 ($R^2$=1; Chi-sq=182.0; P-value<0.0001). We found many exact-matched *trans*-pQTLs between the two sets of analyses, such as rs77542162 (**eFigure 9**). We also noted that these results' effect sizes ($\beta$ coefficient) are concordant regarding the effect directions, albeit with one exception (rs1260326). For this *trans*-pQTL, we then examined the effect alleles used in both analyses and found that Sun et al. and our study defined the effect allele inversely, resulting in an inverse $\beta$ sign. Two sources from Sun et al. are from the full set of the GWAS summary data (https://www.synapse.org/Synapse:syn51365303) and the pQTL tables shared here: https://metabolomips.org/ukbbpgwas/pgwas.table.php. Below are detailed comparisons between the two versions for this specific top lead SNP (rs1260326), along with our ProGWAS for the FLRT2 protein:

- The full set of GWAS is from Sun et al.:

CHROM GENPOS ID ALLELE0 ALLELE1 A1FREQ INFO N TEST BETA SE CHISQ LOG10P
2 27508073 2:27730940:T:C:imp:v1 T C 0.608386 0.998394 33436 ADD 0.051662 0.00702691 54.0522 12.7094

- The pQTL table:

Pos rs ID Locus OA EA EAF Olink ID Target name Uniprot ID Effect SE p-value
2 27,508,073 rs1260326 GCKR T C 0.61 OID20946 Leucine-rich repeat transmembrane protein FLRT2 O43155 0.052 0.007 1.95e-13

- Our ProGWAS for FLRT2:

CHR SNP POS A1 A2 N AF1 BETA SE P
2 rs1260326 27730940 T C 34833 0.390104 -0.0531032 0.00775046 7.30162e-12

We highlighted the effect allele and respective allele frequency in the three different resources.

**eFigure 1: The expression of the 11 MAEs in the UKBB test dataset**

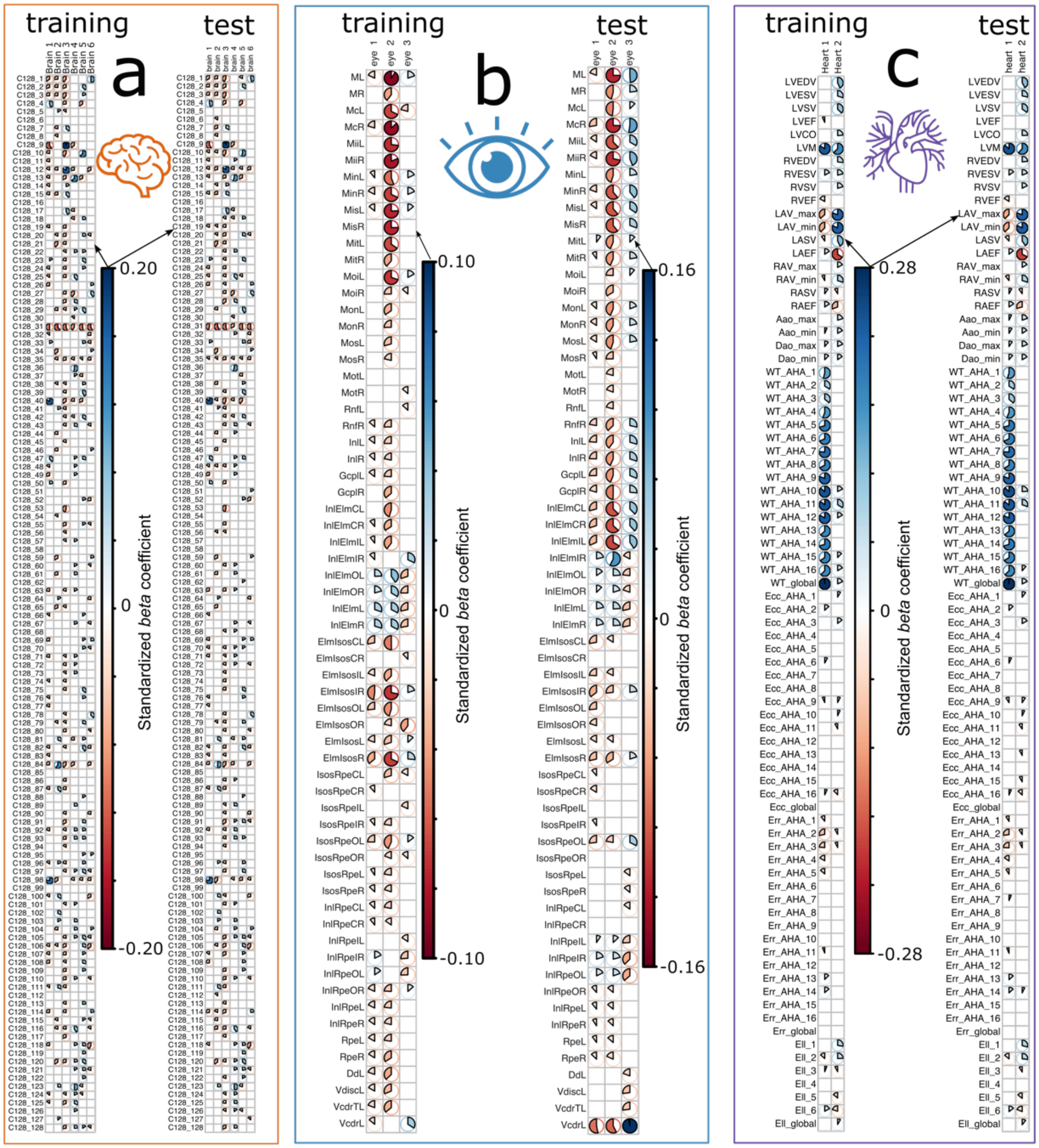


We applied the trained Surreal-GAN model to the UKBB test dataset. **a**) The imaging patterns of the 6 brain MAEs in both the training and test datasets, presenting with the standardized $\beta$ values as effect sizes. **b**) The imaging patterns of the 3 eye MAEs in both the training and test datasets are shown, with standardized $\beta$ values representing effect sizes. **c**) The imaging patterns of the 2 heart MAEs in both training and test datasets are displayed, also using standardized $\beta$ values as effect sizes.

**eFigure 2: The expression of the 6 MAEs in the UKBB test dataset in the image space**

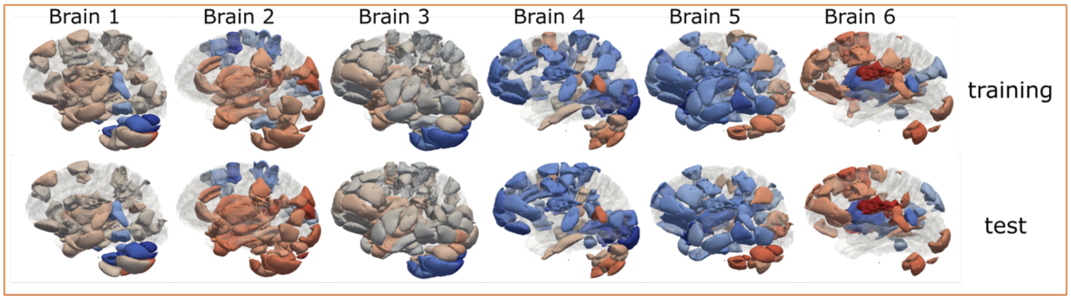


The imaging patterns of the 6 brain MAEs are mapped back into image space, corresponding to **eFigure 1a**.

**eFigure 3: Associations between 11 MAEs in UKBB and clinical features from other organs**

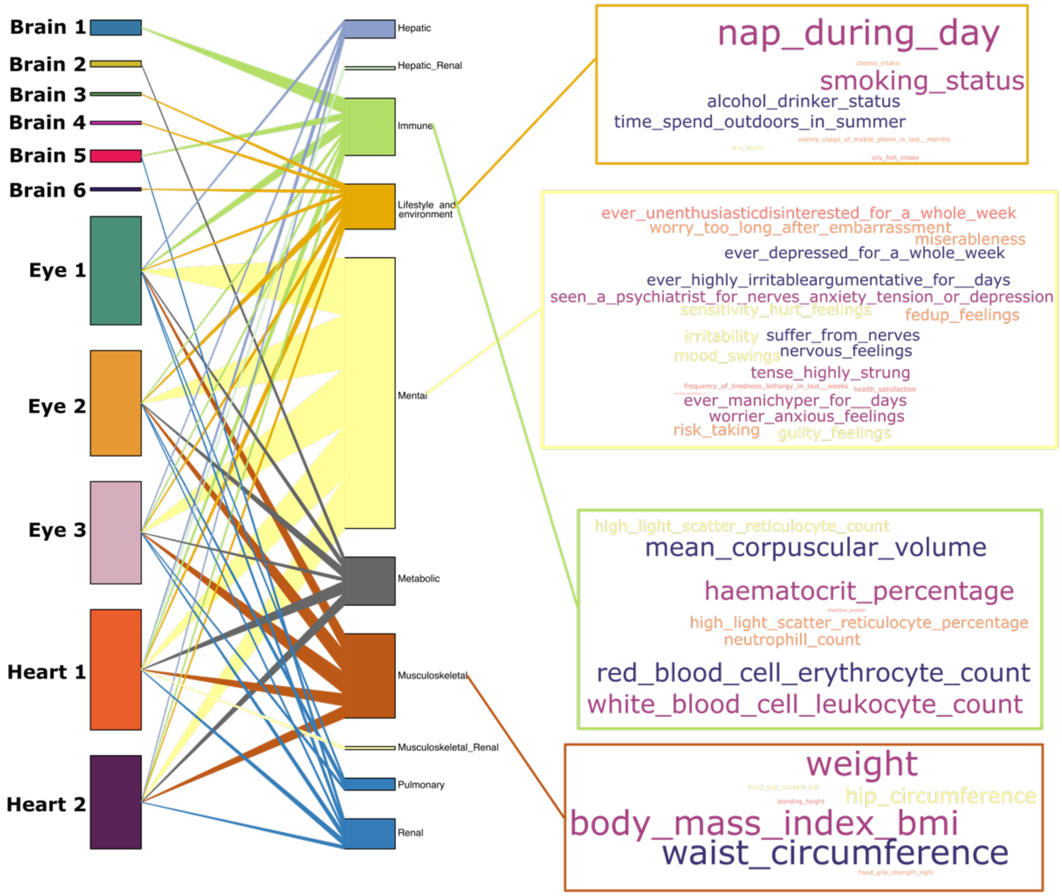


Secondary PWAS associates the 11 MAEs (left panel) to the other 106 clinical phenotypes available in UKBB beyond the brain, eye, and heart IDPs (middle panel). On the right panel, we show the representative traits for each organ system.

**eFigure 4: RNA and protein expression levels of the AP3S2 protein in different tissues using the HPA, GTEx data**

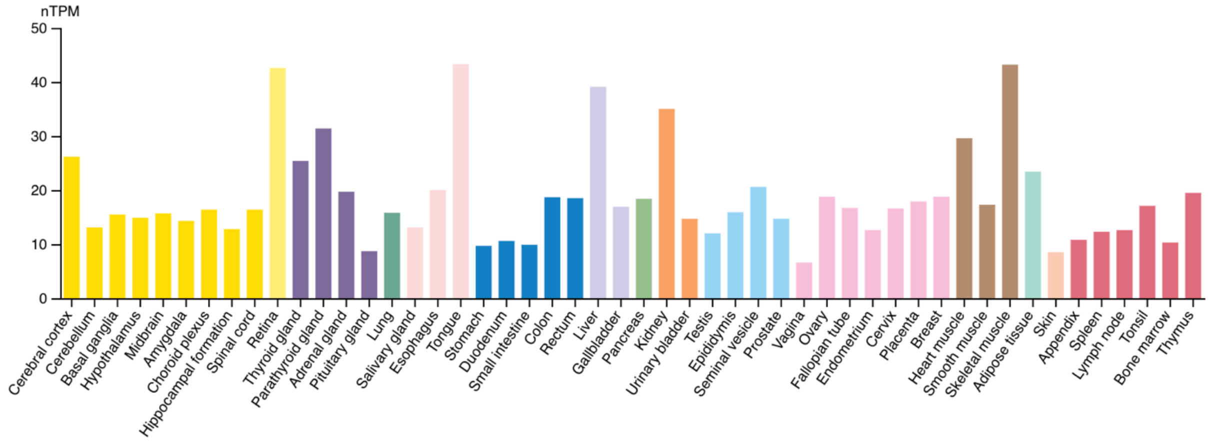


Consensus RNA expression levels of the AP3S2 protein in different tissues using the HPA and GTEx data, quantified by the nTPM.

**eFigure 5: PheWAS results of the top lead SNPs in the GWAS Atlas platform**

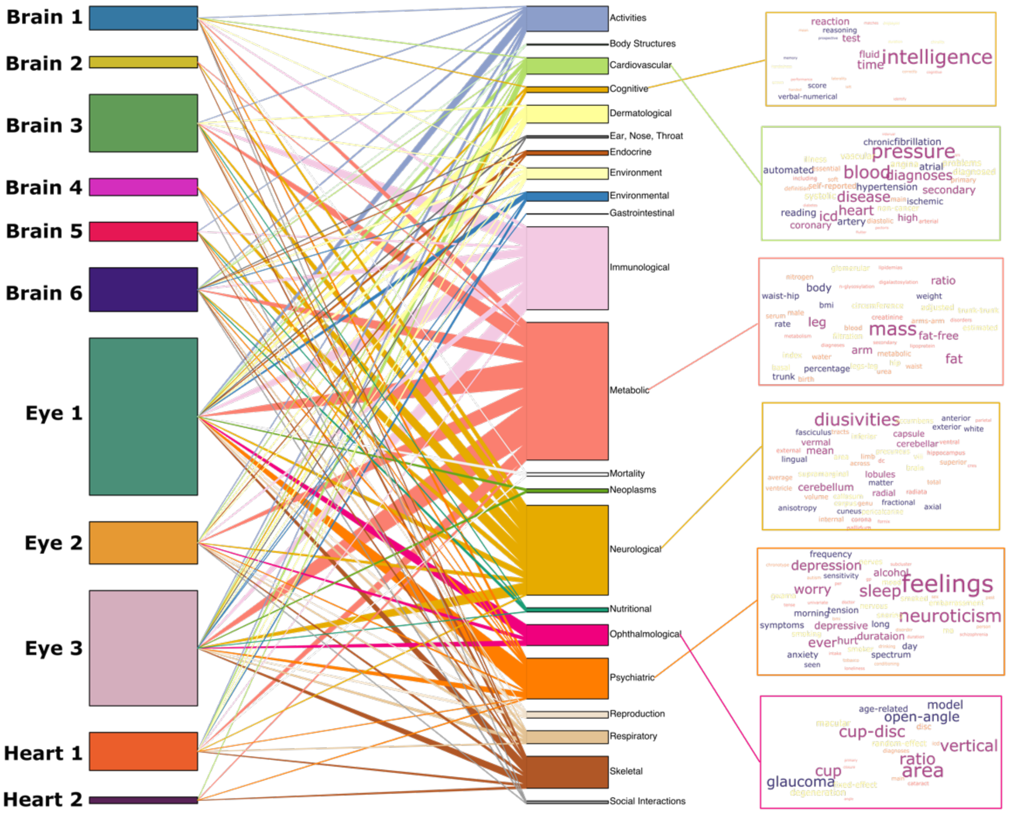


PheWAS associates the 11 MAEs (left panel) to clinical traits in the previous GWASs in the GWAS Atlas platform, denoted by 22 high-level trait domains (middle panel). On the right panel, we show the representative traits for each trait domain.

**eFigure 6a: Manhattan and QQ plots for the brain MAE GWASs**

We present the Manhattan and QQ plots for the 11 MAEs from our primary GWAS using European ancestry populations. All figures are also publicly available at our MEDICINE portal: https://labs-laboratory.com/medicine/eye1_surrealgan (e.g., Eye 1).

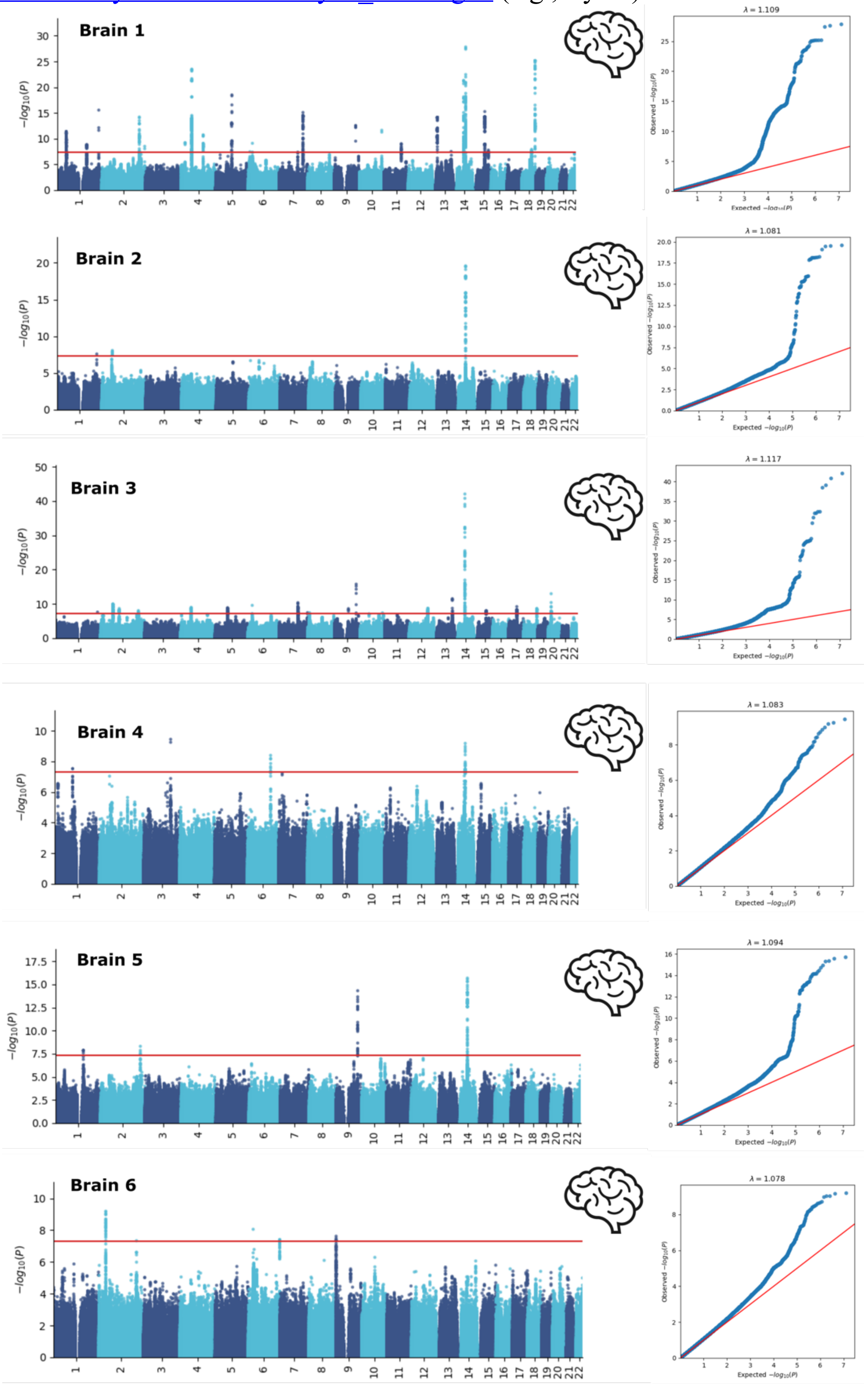

**eFigure 6b: Manhattan and QQ plots for the eye and heart MAE GWASs**

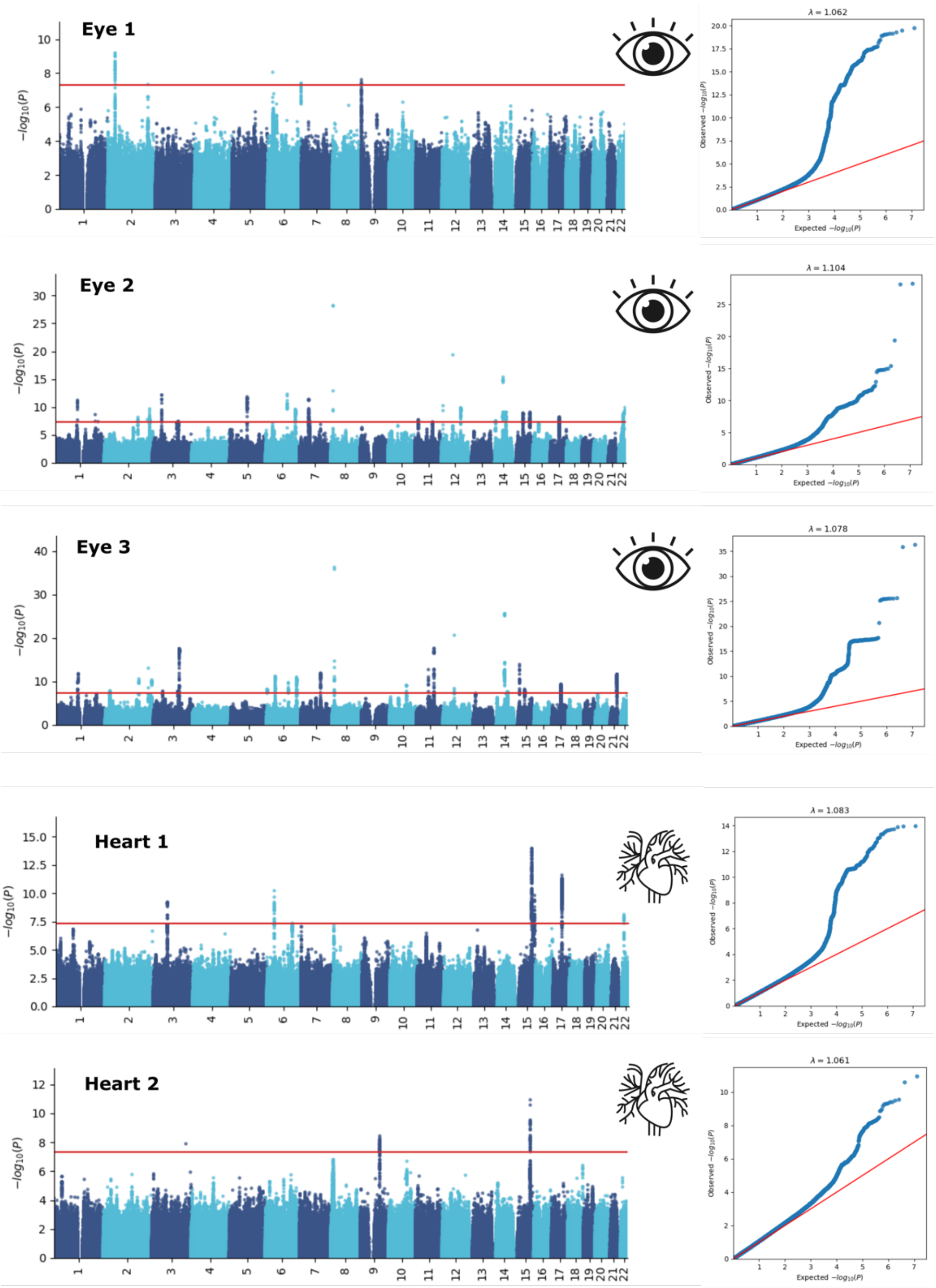

**eFigure 7: The MAPT protein's cancer-type specificity and single-cell specific expression patterns in the brain and breast tissues**

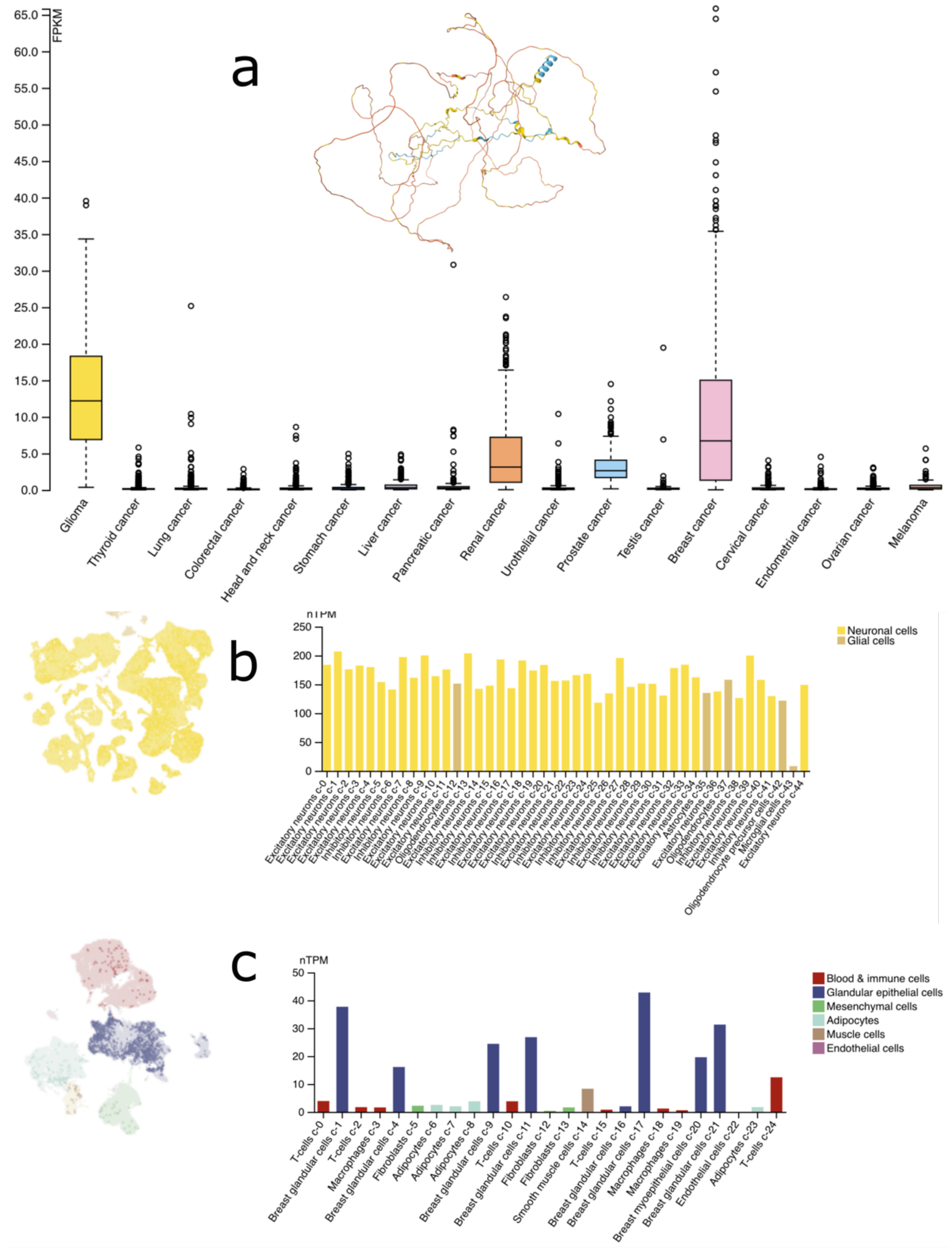


**a**) The RNA expression overview using RNA-seq data in 17 cancer types is reported as the median FPKM (number of Fragments Per Kilobase of exon per million reads) generated by the Cancer Genome Atlas (TCGA). The protein structure was predicted by the AlphaFold model. **b**) Single-cell RNA levels of annotated cell type clusters are shown for the brain tissue via a UMAP plot. The bar chart shows nTPM levels in each annotated cluster of single cells. Color coding is based on cell type groups, each consisting of cell types with functional features in common. **c**) Single-cell RNA levels of annotated cell type clusters are shown for the breast tissue via a UMAP plot. The bar chart shows nTPM levels in each annotated cluster of single cells. Color coding is based on cell type groups, each consisting of cell types with functional features in common.

**eFigure 8: Potential conceptualized causal pathways linking genetics, proteomics, MAEs, and DEs**

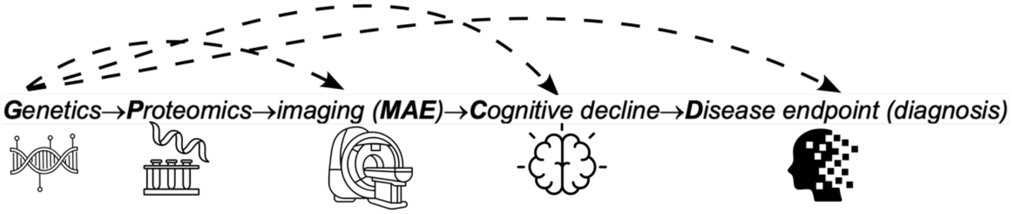


The endophenotype hypothesis suggests that intermediate phenotypes, known as endophenotypes, serve as critical links along the causal pathway from genetic variation to proteomic changes, imaging-derived markers (such as MAEs), cognitive decline, and ultimately, disease outcomes: Genetics → Proteomics → MAE → Cognitive decline → Disease endpoints. Two potential models can cover this pathway: *i*) liability-index model (horizontal pleiotropy), where genetics exert pleiotropic effects on both intermediate phenotypes such as MAE and disease endpoints (dotted arrow line), and *ii*) mediational model (vertical pleiotropy), where assume that genetics exclusively affect disease endpoints via intermediate phenotypes such as MAE. The current study provides empirical evidence to support this hypothesis; however, we do not claim the exclusivity of the two models in real clinical cases.

**eFigure 9: The pQTL identified in the current study vs. those by Sun et al. for the FLRT2 protein**

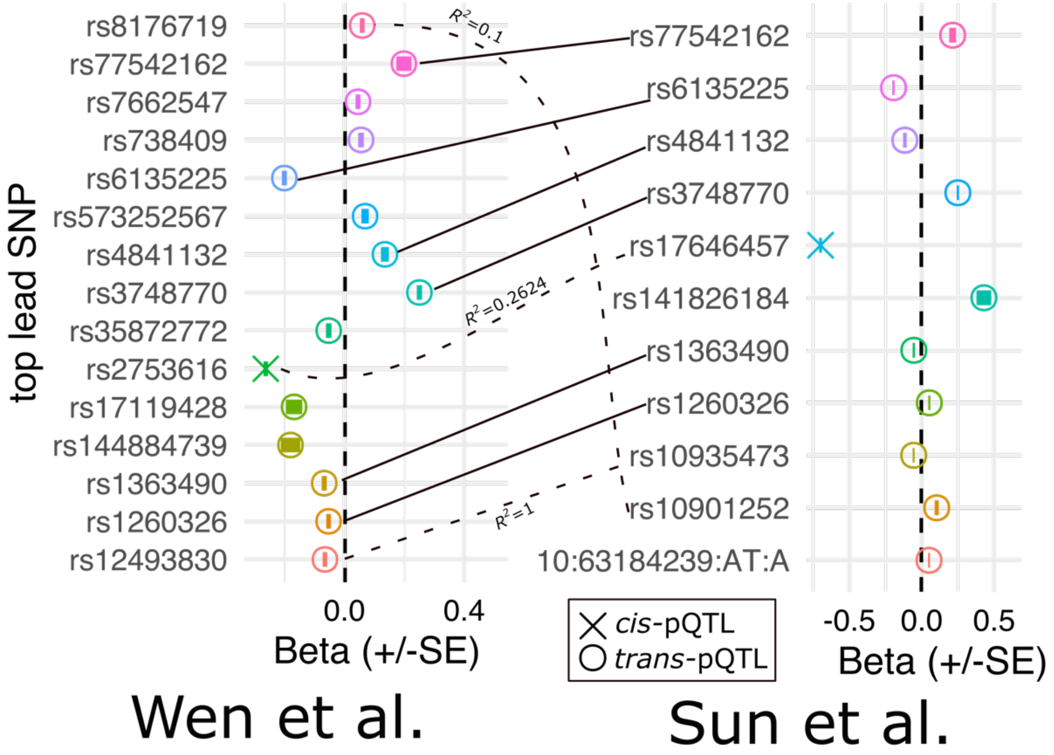


The *cis*- and *trans*-pQTLs identified by the current study and those from Sun et al. are presented. Of note, the same sizes (*N*), GWAS models, criteria to define the genomics loci, etc., may differ between the two studies. We tried to harmonize the procedures as much as possible, including using the same definition for the *trans*- and *cis*-pQTLs. A high concordance was observed between the two sets of analysis.

**eFigure 10: The FLRT2 protein's RNA and single-cell expression profiles**

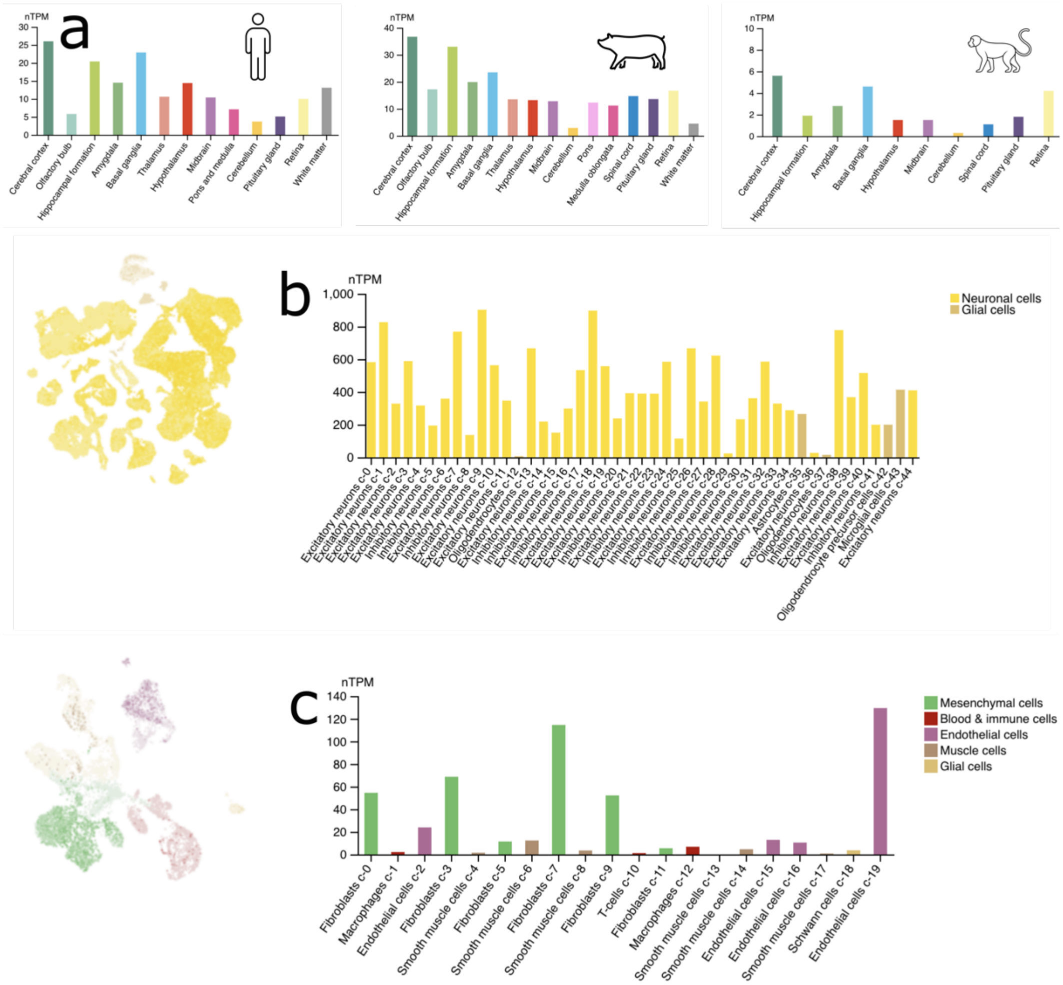


**a**) For the human brain, RNA-seq tissue data generated by the Genotype-Tissue Expression (GTEx) project is reported as mean nTPM, corresponding to mean values of the different individual samples for the respective subregions. For the pig brain, HPA RNA-seq tissue data is reported as mean nTPM (normalized expression) for each of the brain regions analyzed in the pig. For the mouse brain, HPA RNA-seq tissue data is reported as mean nTPM (normalized expression) for each brain region analyzed in the mouse. **b**) Single-cell RNA levels of annotated cell type clusters are shown for the brain tissue via a UMAP plot. The bar chart shows nTPM levels in each annotated cluster of single cells. Color coding is based on cell type groups, each consisting of cell types with functional features in common. **c**) Single-cell RNA levels of annotated cell type clusters are shown for the vascular tissue via a UMAP plot. The bar chart shows nTPM levels in each annotated cluster of single cells. Color coding is based on cell type groups, each consisting of cell types with functional features in common.

**eFigure 11: Group differences for the medication status of Digoxin for Heart 2**

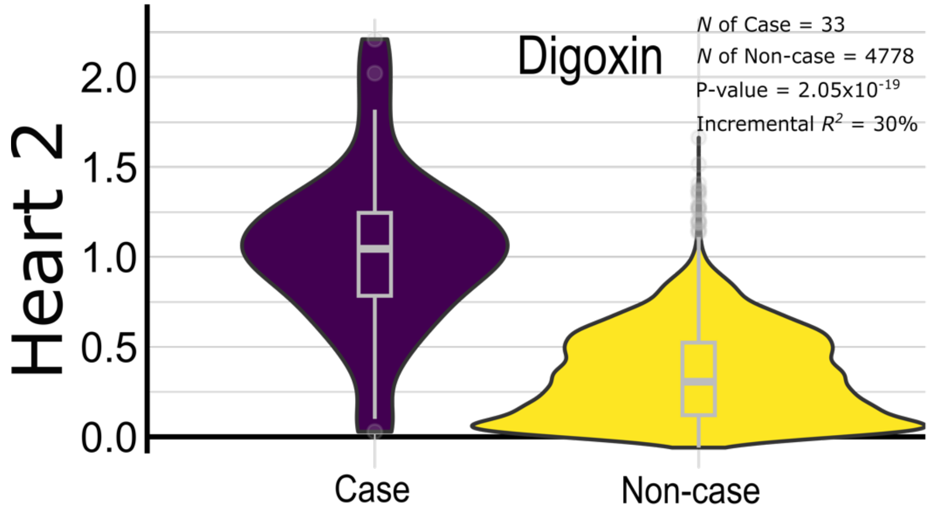


Here, we show the distribution of Heart 2 in the case group (individuals who took the Digoxin drug) vs. in the control group (Non-case or placebo). The box plot (showing the median value) is overlaid on top of the violin plot. All statistics are also shown.

**eFigure 12: Comparisons between the ProWAS results of UKBB Olink and BLSA SomaScan proteins with Brain 4**

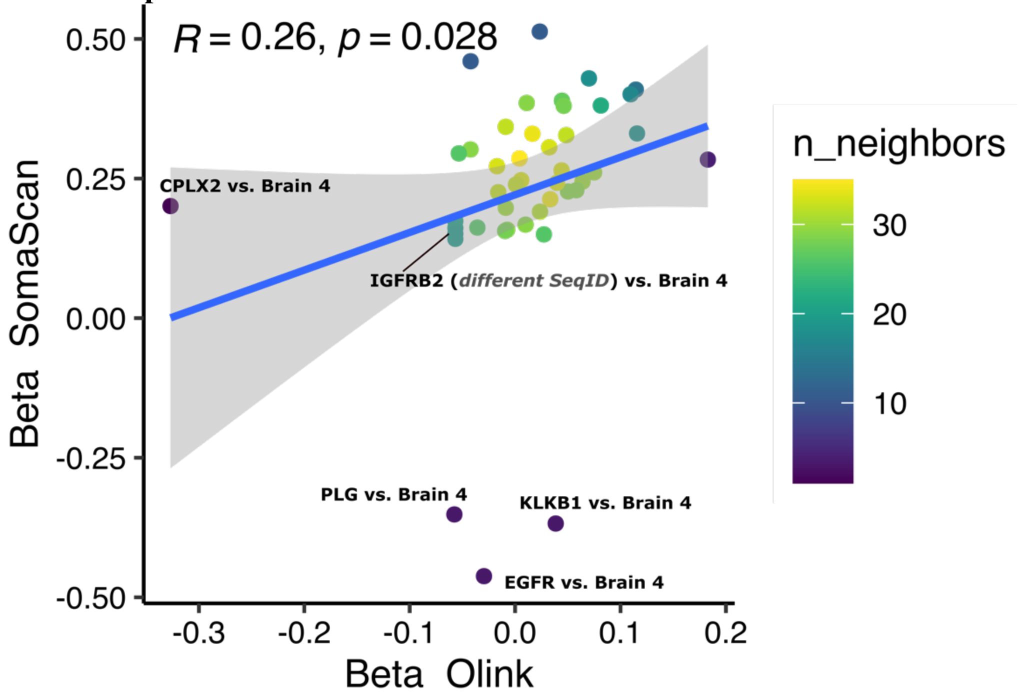


Overall, SomaScan proteins were more sensitive to detecting the brain MAE-protein associations (P-value<0.05/2139 common proteins between the two platforms) than Olink. Here, we show the common significant MAE-protein signals defined by SomaScan for the association with Brain 4 between the $\beta$ coefficient estimates from the two platforms. We also annotated several outliers where the $\beta$ coefficients were opposite across the two platforms. In addition, we also observe that the SomaScan proteins may be quantified under different SeqIDs, each representing distinct aptamers or probes specific to that protein. The associations were robust across different SeqIDs (e.g., IGFRB2 vs. Brain 4).

**eFigure 13: Beta coefficients of the significant SNPs of the GWAS for Brain 1 in UKBB vs. ADNI**

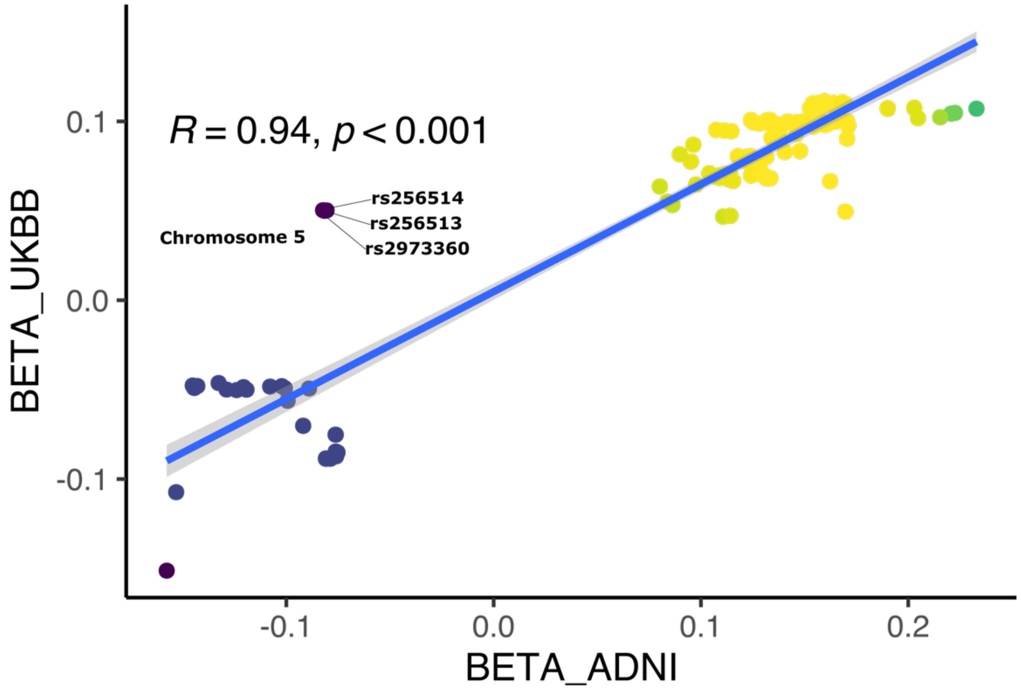


We show the scatter plot for the $\beta$ coefficient from the UKBB GWAS and ADNI GWAS for SNPs that passed the genome-wide significant SNPs. The two sets of estimation showed concordant directions for most SNPs, with exceptions for the annotated three SNPs in high LD on chromosome 5.

**eFigure 14: MAE differences in different ethnic groups compared to European**

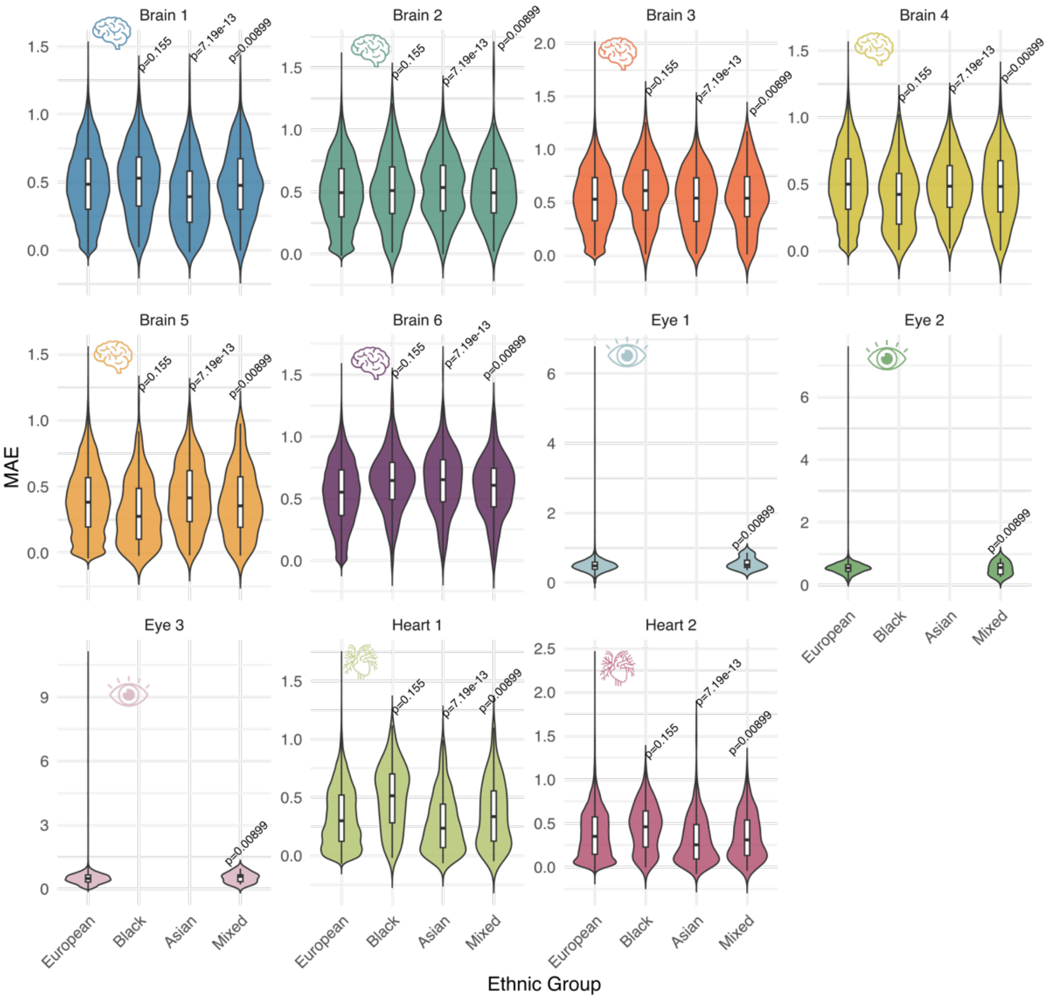


We present MAE differences across ethnic groups, with two-sample t-tests used to assess statistical significance between the European group and each of the other ethnic groups.

**eFigure 15: Model convergence of the 3 Surreal-GAN models based on the R-indices correlation along the training epochs**

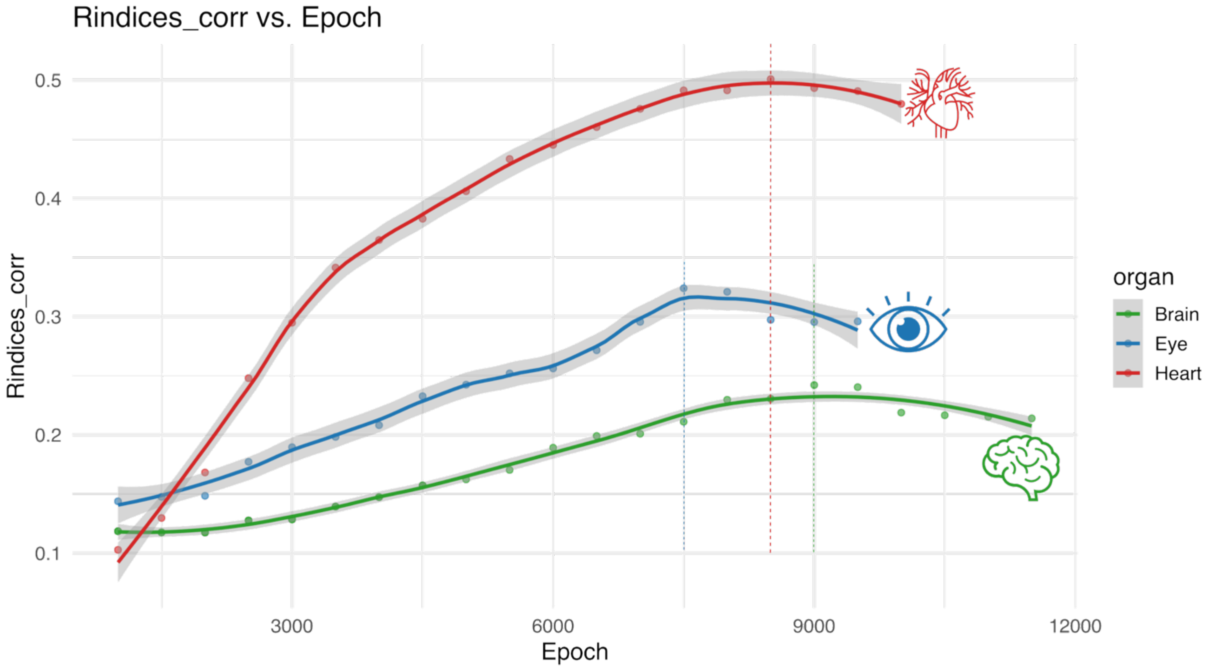


Throughout model training, we report the Rindices-corr, a reproducibility index of the derived MAE. Surreal-GAN models were evaluated every 500 epochs, and training was terminated if no further improvement was observed across 4 consecutive evaluations. Dotted vertical lines indicate the peak Rindices-corr value for each organ-specific Surreal-GAN model.

**eTable 1: The characteristics of the MULTI consortium**

| Data type | Multi-organ/-omics data | Study | BL (*N*) | FU (*N*) | Age [year (mean/std)] | Sex (female) | CN[c] | Pan-disease[e] |
|---|---|---|---|---|---|---|---|---|
| Individual-level data | Brain MRI | UKBB | 42,660 | NA[a] | 63.96±14.11 | 22,505/53% | 6964 | 6959 |
| | | ADNI | 1765[g] | 9752 | 73.66±7.19 | 798/45% | NA[d] | NA |
| | | BLSA | 1114 | NA[a] | 65.44±14.11 | 589/53% | NA[d] | NA |
| | | A4 | 1055 | NA[a] | 72.10±4.75 | 618/59% | NA[d] | NA |
| | Eye OCT | UKBB | 40,063 | NA[a] | 56.71±8.00 | 21,917/55% | 5724 | 5284 |
| | Heart MRI | UKBB | 35,576 | NA[a] | 63.50±7.56 | 18,312/51% | 5791 | 11,746 |
| | Genetics | UKBB | 81,831 | NA[a] | 64.12±7.70[b] | 43,631/53% | NA | NA |
| | | ADNI | 1491 | NA[a] | 73.64±7.15 | 658/44% | NA[d] | NA |
| | Proteomics | UKBB | 50,316 | NA[a] | 64.16±8.04 | 28,581/54% | NA | NA |
| | | BLSA | 924 | NA[a] | 65.30±14.86 | 504/55% | NA[d] | NA |
| | Cognition | UKBB | 39,556 | NA[a] | 63.66±7.55 | 20,939/53% | NA | NA |
| | Clinical trial outcome (PACC) | A4 (treatment) | 516[h] | 5161 | 71.84±4.59 | 297/58% | NA | NA |
| | | A4 (placebo) | 591[h] | 5469 | 71.92±4.97 | 357/60% | NA | NA |
| | Neuropathological biomarkers ($Tau_{181p}$ & $A\beta_{1\text{-}42}$) | UKBB (plasma) | 1156 | NA[a] | 59.37±7.28 | 623/54% | NA | NA |
| | | ADNI (CSF) | 1613 | NA[a] | 71.22±7.09 | 710/44% | NA[d] | NA |
| Summary-level data | GWAS summary statistics | FinnGen | 521[f] | NA | NA | NA | NA | NA |
| | | PGC | 4[f] | NA | NA | NA | NA | NA |

[a]UKBB has released the second session of the imaging data, but the sample sizes are small and were not analyzed in the current study. We only used the MRI/OCT at baseline (BL); BLSA and A4 also have longitudinal follow-up (FU) scans, but we did not include them. ADNI longitudinal FU brain scans were included for individuals with at least 5 scans in the longitudinal analysis, but genetic analysis used only baseline data. [b]We present the mean, minimum, and maximum age at the brain and heart MRI session (2_0). Eye OCT data was collected at the first session (0_0). [c]In UKBB, we define the CN for participants with no ICD code for any diseases or any HER data available to indicate any disease history for the brain, eye, and heart populations. [d]For other independent datasets applied to the trained Surreal-GAN model, the definition of CN[c] was not applied. Some of these participants were healthy controls. [e]Pan-disease of each organ was defined by the ICD-10 code (Data Field: 41270). [f]The number of disease endpoints (DEs) included for the GWAS summary statistics from FinnGen or PGC. [g]For the ADNI brain MRI at baseline, we had 547 CN, 875 MCI, and 343 AD clinical diagnoses. [h]For the PACC (Preclinical Alzheimer's Cognitive Composite) score, we overlapped the baseline measures with the A4 imaging baseline scans and included longitudinal measures for 312 weeks. The number of follow-ups represents the PACC scores, not the imaging scans.

**eTable 2: The associations between the 6 brain MAEs and CSF and plasma neuropathological biomarkers using ADNI and UKBB**

a) **CSF biomarkers in ADNI**: For logP values, the sign indicates the direction of the *beta* coefficient. For *beta* coefficients and Pearson's *r* coefficients, we show only those results that remain significant after correction for multiple comparisons; otherwise, they are represented as 0.

| MAE | Amyloid beta-42 | pTau-181 | Total pTau-181 |
|---|---|---|---|
| Brain1_logP | -1.02716 | 3.166877 | 3.4089 |
| Brain1_beta | 0 | 0.000299 | 3.43E-05 |
| Brain1_r | 0 | 0.162834 | 0.174636 |
| Brain1_samplesize | 1173 | 1173 | 1173 |
| Brain2_logP | -0.00312 | -1.12855 | -0.88663 |
| Brain2_beta | 0 | 0 | 0 |
| Brain2_r | 0 | 0 | 0 |
| Brain2_samplesize | 1173 | 1173 | 1173 |
| Brain3_logP | -0.95584 | 3.902768 | 4.513234 |
| Brain3_beta | 0 | 0.000338 | 4.02E-05 |
| Brain3_r | 0 | 0.151327 | 0.167453 |
| Brain3_samplesize | 1173 | 1173 | 1173 |
| Brain4_logP | 0.236291 | -0.91049 | -0.9979 |
| Brain4_beta | 0 | 0 | 0 |
| Brain4_r | 0 | 0 | 0 |
| Brain4_samplesize | 1173 | 1173 | 1173 |
| Brain5_logP | 5.040649 | -1.10102 | -1.21941 |
| Brain5_beta | 1.59E-05 | 0 | 0 |
| Brain5_r | 0.166839 | 0 | 0 |
| Brain5_samplesize | 1173 | 1173 | 1173 |
| Brain6_logP | -0.72809 | -3.26786 | -3.84501 |
| Brain6_beta | 0 | -0.00028 | -3.4E-05 |
| Brain6_r | 0 | -0.00935 | -0.00288 |
| Brain6_samplesize | 1173 | 1173 | 1173 |

b) **Plasma biomarkers in UKBB**: For logP values, the sign indicates the direction of the *beta* coefficient. For *beta* coefficients and Pearson's *r* coefficients, we show only those results that remain significant after correction for multiple comparisons; otherwise, they are represented as 0.

| MAE | Amyloid beta-42 | pTau-181 |
|---|---|---|
| Brain1_logP | -0.16969 | -0.53928 |
| Brain1_beta | -0.00238 | -0.01484 |
| Brain1_r | -0.01455 | -0.01447 |
| Brain1_samplesize | 1156 | 1156 |
| Brain2_logP | 0.544856 | -0.46775 |
| Brain2_beta | 0.006379 | -0.01381 |
| Brain2_r | 0.032799 | -0.02128 |
| Brain2_samplesize | 1156 | 1156 |
| Brain3_logP | -0.08944 | -0.03777 |
| Brain3_beta | -0.00145 | -0.00158 |
| Brain3_r | -0.00921 | -0.00226 |
| Brain3_samplesize | 1156 | 1156 |
| Brain4_logP | 0.023945 | 0.311258 |
| Brain4_beta | 0.000388 | 0.009827 |
| Brain4_r | 0.000148 | 0.018732 |
| Brain4_samplesize | 1156 | 1156 |

| | | |
|---|---|---|
| Brain5_logP | -0.47182 | 1.080675 |
| Brain5_beta | -0.00536 | 0.023816 |
| Brain5_r | -0.02446 | 0.040052 |
| Brain5_samplesize | 1156 | 1156 |
| Brain6_logP | -0.18665 | -1.80173 |
| Brain6_beta | -0.00256 | -0.03306 |
| Brain6_r | -0.01412 | -0.05738 |
| Brain6_samplesize | 1156 | 1156 |

**eTable 3: The associations between the 6 brain MAEs and 8 cognitive scores using UKBB**

For the logP values, the sign indicates the direction of the *beta* coefficient. For *beta* coefficients and Pearson's *r* coefficients, we only show significant results after correction for multiple comparisons; otherwise, they are represented as 0. In the column header, the code after "_f" indicates the Field ID from the UKBB website (e.g., https://biobank.ndph.ox.ac.uk/showcase/field.cgi?id=21004).

| MAE | number_of_puzzles_correct_f21004 | number_of_symbol_digit_matches_made_correctly_f23324 | number_of_puzzles_correctly_solved_f6373 | duration_to_complete_numeric_path_trail_1_f6348 | duration_to_complete_alphanumeric_path_trail_2_f6350 | fluid_intelligence_score_f20016 | maximum_digits_remembered_correctly_f4282 | mean_time_to_correctly_identify_matches_f20023 |
|---|---|---|---|---|---|---|---|---|
| Brain 1_logP | -0.88571 | -2.54466 | -4.62974 | 2.22784 | 1.805518 | -3.05995 | -2.57144 | 1.870168 |
| Brain 1_beta | 0 | -0.001 | -0.00324 | 5.23E-05 | 0 | -0.00217 | -0.00311 | 0 |
| Brain 1_r | 0 | -0.02564 | -0.02919 | 0.022816 | 0 | -0.0174 | -0.01988 | 0 |
| Brain 1_samplesize | 25510 | 25764 | 25725 | 26001 | 26001 | 36409 | 26725 | 36912 |
| Brain 2_logP | -1.1092 | -1.92104 | -7.03157 | 1.240014 | 0.91905 | -20.1589 | -15.8058 | 0.565144 |
| Brain 2_beta | 0 | 0 | -0.00422 | 0 | 0 | -0.00628 | -0.00883 | 0 |
| Brain 2_r | 0 | 0 | -0.02865 | 0 | 0 | -0.04634 | -0.04724 | 0 |
| Brain 2_samplesize | 25510 | 25764 | 25725 | 26001 | 26001 | 36409 | 26725 | 36912 |
| Brain 3_logP | -4.24585 | -1.63763 | -6.15466 | 1.319683 | 2.507764 | -10.9673 | -2.44256 | -0.53498 |
| Brain 3_beta | -0.00221 | 0 | -0.00413 | 0 | 1.94E-05 | -0.0048 | -0.00329 | 0 |
| Brain 3_r | -0.02571 | 0 | -0.03132 | 0 | 0.018812 | -0.03508 | -0.01794 | 0 |
| Brain 3_samplesize | 25510 | 25764 | 25725 | 26001 | 26001 | 36409 | 26725 | 36912 |
| Brain 4_logP | 1.084621 | 0.676013 | 1.936956 | -2.21034 | -0.83972 | 0.92875 | -0.27489 | -0.87017 |
| Brain 4_beta | 0 | 0 | 0 | -5.3E-05 | 0 | 0 | 0 | 0 |
| Brain 4_r | 0 | 0 | 0 | -0.01856 | 0 | 0 | 0 | 0 |
| Brain 4_samplesize | 25510 | 25764 | 25725 | 26001 | 26001 | 36409 | 26725 | 36912 |
| Brain 5_logP | 0.57791 | -0.59865 | 0.47816 | -0.3325 | -1.20472 | 1.464563 | 1.012731 | 1.144412 |
| Brain 5_beta | 0 | 0 | 0 | 0 | 0 | 0 | 0 | 0 |
| Brain 5_r | 0 | 0 | 0 | 0 | 0 | 0 | 0 | 0 |
| Brain 5_samplesize | 25510 | 25764 | 25725 | 26001 | 26001 | 36409 | 26725 | 36912 |
| Brain 6_logP | -2.88773 | -0.3783 | -1.89179 | 0.9013 | 3.558996 | -2.69358 | -0.21136 | -0.76279 |
| Brain 6_beta | -0.00162 | 0 | 0 | 0 | 2.19E-05 | -0.00201 | 0 | 0 |
| Brain 6_r | -0.02266 | 0 | 0 | 0 | 0.0272 | -0.01609 | 0 | 0 |
| Brain 6_sa | 25510 | 25764 | 25725 | 26001 | 26001 | 36409 | 26725 | 36912 |

| mples ize |
| --- |

730

**eTable 4: The associations between the 11 MAEs and 9 BAGs using UKBB**

We linked the 9 MAEs with 11 multi-organ BAG (MOBAG) from our previous study. P-value is shown at the -log10 scale.

| MAE | Organ | MOBAG | *N* | *Beta* | SE | logP |
|---|---|---|---|---|---|---|
| r1_brain | brain | Brain_age_gap | 34685 | 0.030744 | 0.001636 | 77.7103 |
| r2_brain | brain | Brain_age_gap | 34685 | 0.007166 | 0.00165 | 4.849222 |
| r3_brain | brain | Brain_age_gap | 34685 | 0.01472 | 0.001652 | 18.27012 |
| r4_brain | brain | Brain_age_gap | 34685 | -0.01629 | 0.001653 | 22.14555 |
| r5_brain | brain | Brain_age_gap | 34685 | -0.01326 | 0.001637 | 15.23931 |
| r6_brain | brain | Brain_age_gap | 34685 | 0.001622 | 0.001653 | 0.486276 |
| r1_brain | brain | Eye_age_gap | 3564 | 0.002731 | 0.00505 | 0.230116 |
| r2_brain | brain | Eye_age_gap | 3564 | -0.0021 | 0.005101 | 0.1668 |
| r3_brain | brain | Eye_age_gap | 3564 | 0.002017 | 0.00511 | 0.159259 |
| r4_brain | brain | Eye_age_gap | 3564 | -0.00123 | 0.005119 | 0.091147 |
| r5_brain | brain | Eye_age_gap | 3564 | 0.004542 | 0.00506 | 0.432418 |
| r6_brain | brain | Eye_age_gap | 3564 | -0.00662 | 0.005114 | 0.708581 |
| r1_brain | brain | Cardiovascular_age_gap | 11560 | 0.002169 | 0.002697 | 0.37547 |
| r2_brain | brain | Cardiovascular_age_gap | 11560 | -0.00171 | 0.002703 | 0.277322 |
| r3_brain | brain | Cardiovascular_age_gap | 11560 | 0.003802 | 0.002715 | 0.79176 |
| r4_brain | brain | Cardiovascular_age_gap | 11560 | -0.00315 | 0.002716 | 0.608291 |
| r5_brain | brain | Cardiovascular_age_gap | 11560 | 0.001122 | 0.002688 | 0.169703 |
| r6_brain | brain | Cardiovascular_age_gap | 11560 | 0.003859 | 0.002716 | 0.808715 |
| r1_brain | brain | Pulmonary_age_gap | 11560 | 0.005716 | 0.002725 | 1.444571 |
| r2_brain | brain | Pulmonary_age_gap | 11560 | -0.00186 | 0.002731 | 0.304455 |
| r3_brain | brain | Pulmonary_age_gap | 11560 | 0.00655 | 0.002743 | 1.770527 |
| r4_brain | brain | Pulmonary_age_gap | 11560 | 0.000204 | 0.002745 | 0.026543 |
| r5_brain | brain | Pulmonary_age_gap | 11560 | -0.00273 | 0.002716 | 0.502425 |
| r6_brain | brain | Pulmonary_age_gap | 11560 | -0.00145 | 0.002744 | 0.223422 |
| r1_brain | brain | Musculoskeletal_age_gap | 11560 | 0.004108 | 0.00276 | 0.864257 |
| r2_brain | brain | Musculoskeletal_age_gap | 11560 | 0.003183 | 0.002766 | 0.602318 |
| r3_brain | brain | Musculoskeletal_age_gap | 11560 | 0.005457 | 0.002779 | 1.304938 |
| r4_brain | brain | Musculoskeletal_age_gap | 11560 | 0.001095 | 0.00278 | 0.158791 |
| r5_brain | brain | Musculoskeletal_age_gap | 11560 | -0.00564 | 0.002751 | 1.394905 |
| r6_brain | brain | Musculoskeletal_age_gap | 11560 | -0.004 | 0.002779 | 0.823397 |
| r1_brain | brain | Immune_age_gap | 11560 | 0.001675 | 0.002668 | 0.275699 |
| r2_brain | brain | Immune_age_gap | 11560 | 0.001516 | 0.002673 | 0.243557 |
| r3_brain | brain | Immune_age_gap | 11560 | 0.002408 | 0.002686 | 0.431796 |
| r4_brain | brain | Immune_age_gap | 11560 | -0.00375 | 0.002686 | 0.787233 |
| r5_brain | brain | Immune_age_gap | 11560 | -0.0035 | 0.002659 | 0.725791 |
| r6_brain | brain | Immune_age_gap | 11560 | 0.00055 | 0.002686 | 0.076902 |
| r1_brain | brain | Renal_age_gap | 11560 | 0.001158 | 0.002287 | 0.212741 |
| r2_brain | brain | Renal_age_gap | 11560 | 0.005273 | 0.002291 | 1.669913 |
| r3_brain | brain | Renal_age_gap | 11560 | -0.00234 | 0.002302 | 0.510344 |
| r4_brain | brain | Renal_age_gap | 11560 | 0.001056 | 0.002303 | 0.189448 |
| r5_brain | brain | Renal_age_gap | 11560 | -0.00243 | 0.002279 | 0.544398 |
| r6_brain | brain | Renal_age_gap | 11560 | -0.00463 | 0.002302 | 1.351877 |
| r1_brain | brain | Hepatic_age_gap | 11560 | 0.001578 | 0.002708 | 0.251853 |
| r2_brain | brain | Hepatic_age_gap | 11560 | -0.00231 | 0.002714 | 0.404676 |
| r3_brain | brain | Hepatic_age_gap | 11560 | -0.00196 | 0.002726 | 0.32539 |
| r4_brain | brain | Hepatic_age_gap | 11560 | 0.003088 | 0.002727 | 0.589292 |
| r5_brain | brain | Hepatic_age_gap | 11560 | -0.00028 | 0.002699 | 0.037211 |
| r6_brain | brain | Hepatic_age_gap | 11560 | -0.00122 | 0.002727 | 0.183356 |
| r1_brain | brain | Metabolic_age_gap | 11560 | 0.00341 | 0.002713 | 0.680398 |
| r2_brain | brain | Metabolic_age_gap | 11560 | -0.00499 | 0.002719 | 1.179023 |
| r3_brain | brain | Metabolic_age_gap | 11560 | 0.002036 | 0.002731 | 0.340893 |
| r4_brain | brain | Metabolic_age_gap | 11560 | 0.001209 | 0.002732 | 0.18168 |
| r5_brain | brain | Metabolic_age_gap | 11560 | 0.003854 | 0.002704 | 0.812093 |
| r6_brain | brain | Metabolic_age_gap | 11560 | 0.001293 | 0.002732 | 0.196544 |
| r1_eye | eye | Brain_age_gap | 3222 | 0.015811 | 0.0054 | 2.46425 |
| r2_eye | eye | Brain_age_gap | 3217 | 0.026376 | 0.005356 | 6.052089 |
| r3_eye | eye | Brain_age_gap | 3219 | 0.017714 | 0.005385 | 2.993463 |
| r1_eye | eye | Eye_age_gap | 37494 | 0.037294 | 0.001654 | 111.1567 |
| r2_eye | eye | Eye_age_gap | 37498 | 0.059242 | 0.001635 | 281.9378 |
| r3_eye | eye | Eye_age_gap | 37498 | 0.025034 | 0.001658 | 50.65713 |
| r1_eye | eye | Cardiovascular_age_gap | 12543 | -0.00019 | 0.002466 | 0.028154 |
| r2_eye | eye | Cardiovascular_age_gap | 12537 | 0.002911 | 0.002468 | 0.623277 |
| r3_eye | eye | Cardiovascular_age_gap | 12537 | 0.000355 | 0.002464 | 0.052863 |
| r1_eye | eye | Pulmonary_age_gap | 12543 | 0.010121 | 0.002508 | 4.260645 |

| | | | | | | |
|---|---|---|---|---|---|---|
| r2_eye | eye | Pulmonary_age_gap | 12537 | 0.006702 | 0.002511 | 2.118621 |
| r3_eye | eye | Pulmonary_age_gap | 12537 | 0.011774 | 0.002504 | 5.585402 |
| r1_eye | eye | Musculoskeletal_age_gap | 12543 | 0.001786 | 0.00285 | 0.275067 |
| r2_eye | eye | Musculoskeletal_age_gap | 12537 | 0.002634 | 0.00285 | 0.449246 |
| r3_eye | eye | Musculoskeletal_age_gap | 12537 | 0.001213 | 0.002846 | 0.174017 |
| r1_eye | eye | Immune_age_gap | 12543 | 0.009091 | 0.00295 | 2.684985 |
| r2_eye | eye | Immune_age_gap | 12537 | 0.004529 | 0.002953 | 0.9028 |
| r3_eye | eye | Immune_age_gap | 12537 | 0.006844 | 0.002946 | 1.694809 |
| r1_eye | eye | Renal_age_gap | 12543 | 0.004057 | 0.002068 | 1.302278 |
| r2_eye | eye | Renal_age_gap | 12537 | 0.004641 | 0.002069 | 1.604295 |
| r3_eye | eye | Renal_age_gap | 12537 | 0.003679 | 0.002065 | 1.126004 |
| r1_eye | eye | Hepatic_age_gap | 12543 | 0.000887 | 0.002464 | 0.143316 |
| r2_eye | eye | Hepatic_age_gap | 12537 | 0.000528 | 0.002463 | 0.080801 |
| r3_eye | eye | Hepatic_age_gap | 12537 | 0.001086 | 0.002459 | 0.181352 |
| r1_eye | eye | Metabolic_age_gap | 12543 | 5.27E-05 | 0.002376 | 0.007753 |
| r2_eye | eye | Metabolic_age_gap | 12537 | 0.001692 | 0.002375 | 0.322197 |
| r3_eye | eye | Metabolic_age_gap | 12537 | 0.004215 | 0.00237 | 1.123113 |
| r1_heart | heart | Brain_age_gap | 25513 | 0.009914 | 0.001681 | 8.427728 |
| r2_heart | heart | Brain_age_gap | 25513 | 0.019304 | 0.001794 | 26.22868 |
| r1_heart | heart | Eye_age_gap | 2774 | 0.011297 | 0.004899 | 1.67411 |
| r2_heart | heart | Eye_age_gap | 2774 | 0.008044 | 0.005248 | 0.901598 |
| r1_heart | heart | Cardiovascular_age_gap | 9182 | 0.040362 | 0.002773 | 46.722 |
| r2_heart | heart | Cardiovascular_age_gap | 9182 | 0.042809 | 0.002938 | 46.82388 |
| r1_heart | heart | Pulmonary_age_gap | 9182 | 0.00965 | 0.002655 | 3.554119 |
| r2_heart | heart | Pulmonary_age_gap | 9182 | 0.013161 | 0.002811 | 5.540257 |
| r1_heart | heart | Musculoskeletal_age_gap | 9182 | -0.00553 | 0.002704 | 1.3898 |
| r2_heart | heart | Musculoskeletal_age_gap | 9182 | -0.00931 | 0.002863 | 2.940441 |
| r1_heart | heart | Immune_age_gap | 9182 | -0.00417 | 0.002491 | 1.026417 |
| r2_heart | heart | Immune_age_gap | 9182 | -0.00129 | 0.00264 | 0.204456 |
| r1_heart | heart | Renal_age_gap | 9182 | 0.000706 | 0.002249 | 0.122818 |
| r2_heart | heart | Renal_age_gap | 9182 | 0.002622 | 0.002383 | 0.566842 |
| r1_heart | heart | Hepatic_age_gap | 9182 | -0.00733 | 0.002669 | 2.216397 |
| r2_heart | heart | Hepatic_age_gap | 9182 | -0.00411 | 0.002829 | 0.835639 |
| r1_heart | heart | Metabolic_age_gap | 9182 | 0.010788 | 0.002716 | 4.144093 |
| r2_heart | heart | Metabolic_age_gap | 9182 | 0.00185 | 0.00288 | 0.283554 |

**eTable 5: The SNP-based heritability estimates from three methods**

a) GCTA using individual-level genotype data:

| MAE | Heritability | SE | P-value |
|---|---|---|---|
| r1_brain | 0.469826 | 0.021313 | 2.8E-116 |
| r2_brain | 0.31613 | 0.021084 | 3.5E-56 |
| r3_brain | 0.531127 | 0.021115 | 1.6E-158 |
| r4_brain | 0.285673 | 0.020994 | 2.44E-47 |
| r5_brain | 0.435898 | 0.021234 | 6.3E-105 |
| r6_brain | 0.263428 | 0.0208 | 4.51E-45 |
| r1_eye | 0.303643 | 0.01948 | 9.55E-65 |
| r2_eye | 0.403387 | 0.019692 | 5E-107 |
| r3_eye | 0.363571 | 0.019598 | 5.26E-89 |
| r1_heart | 0.385957 | 0.025693 | 1.99E-60 |
| r2_heart | 0.281457 | 0.025717 | 8.43E-32 |

b) LDSC using GWAS summary data:

| MAE | Organ | Heritability | SE |
|---|---|---|---|
| r1 | brain | 0.263 | 0.0245 |
| r2 | brain | 0.1928 | 0.0233 |
| r3 | brain | 0.2981 | 0.0267 |
| r4 | brain | 0.1544 | 0.019 |
| r5 | brain | 0.23 | 0.0238 |
| r6 | brain | 0.142 | 0.0201 |
| r1 | eye | 0.1621 | 0.0218 |
| r2 | eye | 0.2179 | 0.0218 |
| r3 | eye | 0.1811 | 0.0224 |
| r1 | heart | 0.224 | 0.0256 |
| r2 | heart | 0.1465 | 0.0336 |

c) SBayesS using GWAS summary data:

| MAE | h2_mean | h2_se | Organ |
|---|---|---|---|
| r1_brain | 0.231665 | 0.011651 | brain |
| r2_brain | 0.171683 | 0.010561 | brain |
| r3_brain | 0.287042 | 0.012437 | brain |
| r4_brain | 0.152259 | 0.010392 | brain |
| r5_brain | 0.224361 | 0.00973 | brain |
| r6_brain | 0.130989 | 0.011609 | brain |
| r1_eye | 0.140774 | 0.017924 | eye |
| r2_eye | 0.209126 | 0.014578 | eye |
| r3_eye | 0.159633 | 0.010853 | eye |
| r1_heart | 0.214393 | 0.014245 | heart |
| r2_heart | 0.146555 | 0.01236 | heart |

**eTable 6: The polygenicity and nature selection signatures of the 11 MAEs**

| MAE | S | S_se | Pi | Pi_se | Organ |
|---|---|---|---|---|---|
| r1_brain | -0.64678 | 0.104669 | 0.004576 | 0.000606 | brain |
| r2_brain | -0.41141 | 0.08688 | 0.05176 | 0.00329 | brain |
| r3_brain | -0.47562 | 0.18022 | 0.009497 | 0.003948 | brain |
| r4_brain | -0.64135 | 0.136625 | 0.050595 | 0.002625 | brain |
| r5_brain | -0.67266 | 0.165529 | 0.03259 | 0.007677 | brain |
| r6_brain | -0.60439 | 0.090541 | 0.051486 | 0.003481 | brain |
| r1_eye | -0.6752 | 0.185845 | 0.003521 | 0.003354 | eye |
| r2_eye | -0.48458 | 0.203772 | 0.008018 | 0.005652 | eye |
| r3_eye | -0.88936 | 0.103871 | 0.002367 | 0.00038 | eye |
| r1_heart | -0.31912 | 0.220114 | 0.053044 | 0.007679 | heart |
| r2_heart | -0.55014 | 0.182635 | 0.051122 | 0.00347 | heart |

**eTable 7: The phenotypic and genetic associations between the 11 MAEs**

a) Phenotypic correlation via Pearson's *r*

| MAE1 | MAE2 | Pearson_r | Pvalue |
|---|---|---|---|
| Brain1 | Brain1 | 1 | 0 |
| Brain2 | Brain1 | 0.033777 | 0 |
| Brain3 | Brain1 | 0.032657 | 0 |
| Brain4 | Brain1 | 0.022368 | 0 |
| Brain5 | Brain1 | -0.10684 | 0 |
| Brain6 | Brain1 | 0.098921 | 0 |
| Eye1 | Brain1 | 0.012697 | 0.4548 |
| Eye2 | Brain1 | 0.000968 | 0.9545 |
| Eye3 | Brain1 | 0.007254 | 0.6693 |
| Heart1 | Brain1 | -0.02635 | 0 |
| Heart2 | Brain1 | -0.03312 | 0 |
| Brain1 | Brain2 | 0.033777 | 0 |
| Brain2 | Brain2 | 1 | 0 |
| Brain3 | Brain2 | 0.003009 | 0.5495 |
| Brain4 | Brain2 | 0.069239 | 0 |
| Brain5 | Brain2 | -0.10659 | 0 |
| Brain6 | Brain2 | -0.00081 | 0.8718 |
| Eye1 | Brain2 | 0.00106 | 0.9503 |
| Eye2 | Brain2 | -0.0002 | 0.9906 |
| Eye3 | Brain2 | -0.00273 | 0.8721 |
| Heart1 | Brain2 | 0.021028 | 0.0002 |
| Heart2 | Brain2 | -0.00544 | 0.3282 |
| Brain1 | Brain3 | 0.032657 | 0 |
| Brain2 | Brain3 | 0.003009 | 0.5495 |
| Brain3 | Brain3 | 1 | 0 |
| Brain4 | Brain3 | -0.06137 | 0 |
| Brain5 | Brain3 | -0.12789 | 0 |
| Brain6 | Brain3 | 0.081961 | 0 |
| Eye1 | Brain3 | 0.012233 | 0.4714 |
| Eye2 | Brain3 | 0.001265 | 0.9406 |
| Eye3 | Brain3 | 0.008892 | 0.6006 |
| Heart1 | Brain3 | -0.02081 | 0.0002 |
| Heart2 | Brain3 | -0.01343 | 0.0158 |
| Brain1 | Brain4 | 0.022368 | 0 |
| Brain2 | Brain4 | 0.069239 | 0 |
| Brain3 | Brain4 | -0.06137 | 0 |
| Brain4 | Brain4 | 1 | 0 |
| Brain5 | Brain4 | 0.078412 | 0 |
| Brain6 | Brain4 | -0.00325 | 0.5177 |
| Eye1 | Brain4 | -0.00992 | 0.5594 |
| Eye2 | Brain4 | -0.00613 | 0.7182 |
| Eye3 | Brain4 | -0.00546 | 0.7481 |
| Heart1 | Brain4 | 0.012184 | 0.0285 |
| Heart2 | Brain4 | 0.00116 | 0.8348 |
| Brain1 | Brain5 | -0.10684 | 0 |
| Brain2 | Brain5 | -0.10659 | 0 |
| Brain3 | Brain5 | -0.12789 | 0 |
| Brain4 | Brain5 | 0.078412 | 0 |
| Brain5 | Brain5 | 1 | 0 |
| Brain6 | Brain5 | 0.020302 | 0.0001 |
| Eye1 | Brain5 | 0.007856 | 0.6437 |
| Eye2 | Brain5 | -0.00023 | 0.9893 |
| Eye3 | Brain5 | -0.0003 | 0.986 |
| Heart1 | Brain5 | 0.010716 | 0.054 |
| Heart2 | Brain5 | 0.020631 | 0.0002 |
| Brain1 | Brain6 | 0.098921 | 0 |
| Brain2 | Brain6 | -0.00081 | 0.8718 |
| Brain3 | Brain6 | 0.081961 | 0 |
| Brain4 | Brain6 | -0.00325 | 0.5177 |
| Brain5 | Brain6 | 0.020302 | 0.0001 |
| Brain6 | Brain6 | 1 | 0 |
| Eye1 | Brain6 | 0.005331 | 0.7536 |
| Eye2 | Brain6 | -0.0106 | 0.5328 |
| Eye3 | Brain6 | 5.29E-06 | 0.9998 |
| Heart1 | Brain6 | 0.006164 | 0.2677 |

| | | | |
|---|---|---|---|
| Heart2 | Brain6 | -0.00393 | 0.4799 |
| Brain1 | Eye1 | 0.012697 | 0.4548 |
| Brain2 | Eye1 | 0.00106 | 0.9503 |
| Brain3 | Eye1 | 0.012233 | 0.4714 |
| Brain4 | Eye1 | -0.00992 | 0.5594 |
| Brain5 | Eye1 | 0.007856 | 0.6437 |
| Brain6 | Eye1 | 0.005331 | 0.7536 |
| Eye1 | Eye1 | 1 | 0 |
| Eye2 | Eye1 | 0.738472 | 0 |
| Eye3 | Eye1 | 0.924117 | 0 |
| Heart1 | Eye1 | 0.014355 | 0.423 |
| Heart2 | Eye1 | 0.01875 | 0.2952 |
| Brain1 | Eye2 | 0.000968 | 0.9545 |
| Brain2 | Eye2 | -0.0002 | 0.9906 |
| Brain3 | Eye2 | 0.001265 | 0.9406 |
| Brain4 | Eye2 | -0.00613 | 0.7182 |
| Brain5 | Eye2 | -0.00023 | 0.9893 |
| Brain6 | Eye2 | -0.0106 | 0.5328 |
| Eye1 | Eye2 | 0.738472 | 0 |
| Eye2 | Eye2 | 1 | 0 |
| Eye3 | Eye2 | 0.90418 | 0 |
| Heart1 | Eye2 | 0.023803 | 0.1839 |
| Heart2 | Eye2 | 0.013439 | 0.4532 |
| Brain1 | Eye3 | 0.007254 | 0.6693 |
| Brain2 | Eye3 | -0.00273 | 0.8721 |
| Brain3 | Eye3 | 0.008892 | 0.6006 |
| Brain4 | Eye3 | -0.00546 | 0.7481 |
| Brain5 | Eye3 | -0.0003 | 0.986 |
| Brain6 | Eye3 | 5.29E-06 | 0.9998 |
| Eye1 | Eye3 | 0.924117 | 0 |
| Eye2 | Eye3 | 0.90418 | 0 |
| Eye3 | Eye3 | 1 | 0 |
| Heart1 | Eye3 | 0.008671 | 0.6284 |
| Heart2 | Eye3 | 0.008255 | 0.6449 |
| Brain1 | Heart1 | -0.02635 | 0 |
| Brain2 | Heart1 | 0.021028 | 0.0002 |
| Brain3 | Heart1 | -0.02081 | 0.0002 |
| Brain4 | Heart1 | 0.012184 | 0.0285 |
| Brain5 | Heart1 | 0.010716 | 0.054 |
| Brain6 | Heart1 | 0.006164 | 0.2677 |
| Eye1 | Heart1 | 0.014355 | 0.423 |
| Eye2 | Heart1 | 0.023803 | 0.1839 |
| Eye3 | Heart1 | 0.008671 | 0.6284 |
| Heart1 | Heart1 | 1 | 0 |
| Heart2 | Heart1 | 0.443287 | 0 |
| Brain1 | Heart2 | -0.03312 | 0 |
| Brain2 | Heart2 | -0.00544 | 0.3282 |
| Brain3 | Heart2 | -0.01343 | 0.0158 |
| Brain4 | Heart2 | 0.00116 | 0.8348 |
| Brain5 | Heart2 | 0.020631 | 0.0002 |
| Brain6 | Heart2 | -0.00393 | 0.4799 |
| Eye1 | Heart2 | 0.01875 | 0.2952 |
| Eye2 | Heart2 | 0.013439 | 0.4532 |
| Eye3 | Heart2 | 0.008255 | 0.6449 |
| Heart1 | Heart2 | 0.443287 | 0 |
| Heart2 | Heart2 | 1 | 0 |

b) Genetic correlation via LDSC

| MAE1 | MAE2 | gc_mean | gc_se | Z | P |
|---|---|---|---|---|---|
| r1_brain | r1_brain | 1 | 9.67E-06 | 103436 | 0 |
| r2_brain | r1_brain | -1.3E-05 | 0.0623 | -0.0002 | 0.9998 |
| r3_brain | r1_brain | -0.1973 | 0.0585 | -3.374 | 0.0007 |
| r4_brain | r1_brain | -0.0447 | 0.0703 | -0.6351 | 0.5254 |
| r5_brain | r1_brain | -0.2484 | 0.0604 | -4.1137 | 3.89E-05 |
| r6_brain | r1_brain | 0.0025 | 0.071 | 0.0356 | 0.9716 |
| r1_eye | r1_brain | -0.0553 | 0.0657 | -0.8423 | 0.3996 |
| r2_eye | r1_brain | -0.0413 | 0.056 | -0.7372 | 0.461 |
| r3_eye | r1_brain | -0.0868 | 0.0622 | -1.3959 | 0.1628 |
| r1_heart | r1_brain | -0.1956 | 0.0582 | -3.3592 | 0.0008 |
| r2_heart | r1_brain | -0.1571 | 0.0827 | -1.8999 | 0.0574 |
| r1_brain | r2_brain | -1.3E-05 | 0.0623 | -0.0002 | 0.9998 |
| r2_brain | r2_brain | 1 | 1.19E-06 | 840788.1 | 0 |
| r3_brain | r2_brain | 0.0776 | 0.074 | 1.0496 | 0.2939 |
| r4_brain | r2_brain | 0.3485 | 0.0799 | 4.3632 | 1.28E-05 |
| r5_brain | r2_brain | -0.3725 | 0.0723 | -5.1549 | 2.54E-07 |
| r6_brain | r2_brain | -0.3215 | 0.0804 | -3.9995 | 6.35E-05 |
| r1_eye | r2_brain | -0.0366 | 0.0891 | -0.4113 | 0.6809 |
| r2_eye | r2_brain | 0.0556 | 0.0713 | 0.7799 | 0.4355 |
| r3_eye | r2_brain | 0.0051 | 0.0688 | 0.0743 | 0.9408 |
| r1_heart | r2_brain | -0.0896 | 0.0698 | -1.2836 | 0.1993 |
| r2_heart | r2_brain | -0.222 | 0.1088 | -2.0395 | 0.0414 |
| r1_brain | r3_brain | -0.1973 | 0.0585 | -3.374 | 0.0007 |
| r2_brain | r3_brain | 0.0776 | 0.074 | 1.0496 | 0.2939 |
| r3_brain | r3_brain | 1 | 2.55E-07 | 3916574 | 0 |
| r4_brain | r3_brain | -0.226 | 0.072 | -3.1407 | 0.0017 |
| r5_brain | r3_brain | -0.2844 | 0.069 | -4.124 | 3.72E-05 |
| r6_brain | r3_brain | -0.1142 | 0.0781 | -1.4619 | 0.1438 |
| r1_eye | r3_brain | 0.1355 | 0.0677 | 2.0001 | 0.0455 |
| r2_eye | r3_brain | 0.1318 | 0.0617 | 2.1365 | 0.0326 |
| r3_eye | r3_brain | 0.1131 | 0.0603 | 1.8767 | 0.0606 |
| r1_heart | r3_brain | -0.082 | 0.0655 | -1.2512 | 0.2109 |
| r2_heart | r3_brain | -0.1561 | 0.1105 | -1.4125 | 0.1578 |
| r1_brain | r4_brain | -0.0447 | 0.0703 | -0.6351 | 0.5254 |
| r2_brain | r4_brain | 0.3485 | 0.0799 | 4.3632 | 1.28E-05 |
| r3_brain | r4_brain | -0.226 | 0.072 | -3.1407 | 0.0017 |
| r4_brain | r4_brain | 1 | 1.41E-06 | 711050.5 | 0 |
| r5_brain | r4_brain | -0.0927 | 0.0733 | -1.2648 | 0.2059 |
| r6_brain | r4_brain | -0.0738 | 0.0811 | -0.9099 | 0.3629 |
| r1_eye | r4_brain | -0.0869 | 0.0906 | -0.9591 | 0.3375 |
| r2_eye | r4_brain | 0.0653 | 0.0762 | 0.8573 | 0.3913 |
| r3_eye | r4_brain | -0.0832 | 0.0809 | -1.029 | 0.3035 |
| r1_heart | r4_brain | 0.0371 | 0.0845 | 0.4392 | 0.6605 |
| r2_heart | r4_brain | -0.1127 | 0.1154 | -0.9763 | 0.3289 |
| r1_brain | r5_brain | -0.2484 | 0.0604 | -4.1137 | 3.89E-05 |
| r2_brain | r5_brain | -0.3725 | 0.0723 | -5.1549 | 2.54E-07 |
| r3_brain | r5_brain | -0.2844 | 0.069 | -4.124 | 3.72E-05 |
| r4_brain | r5_brain | -0.0927 | 0.0733 | -1.2648 | 0.2059 |
| r5_brain | r5_brain | 1 | 3.51E-08 | 28512886 | 0 |
| r6_brain | r5_brain | -0.0154 | 0.0888 | -0.1739 | 0.862 |
| r1_eye | r5_brain | -0.028 | 0.0795 | -0.352 | 0.7248 |
| r2_eye | r5_brain | 0.006 | 0.0684 | 0.0884 | 0.9296 |
| r3_eye | r5_brain | -0.0286 | 0.0709 | -0.4042 | 0.6861 |
| r1_heart | r5_brain | 0.1822 | 0.066 | 2.7605 | 0.0058 |
| r2_heart | r5_brain | 0.2434 | 0.1024 | 2.3757 | 0.0175 |
| r1_brain | r6_brain | 0.0025 | 0.071 | 0.0356 | 0.9716 |
| r2_brain | r6_brain | -0.3215 | 0.0804 | -3.9995 | 6.35E-05 |
| r3_brain | r6_brain | -0.1142 | 0.0781 | -1.4619 | 0.1438 |
| r4_brain | r6_brain | -0.0738 | 0.0811 | -0.9099 | 0.3629 |
| r5_brain | r6_brain | -0.0154 | 0.0888 | -0.1739 | 0.862 |
| r6_brain | r6_brain | 1 | 1.07E-05 | 93306.06 | 0 |
| r1_eye | r6_brain | 0.0311 | 0.0831 | 0.3742 | 0.7083 |
| r2_eye | r6_brain | 0.009 | 0.0766 | 0.117 | 0.9069 |
| r3_eye | r6_brain | 0.0899 | 0.086 | 1.0456 | 0.2957 |
| r1_heart | r6_brain | -0.0638 | 0.0957 | -0.6663 | 0.5052 |
| r2_heart | r6_brain | 0.0251 | 0.1127 | 0.2225 | 0.824 |
| r1_brain | r1_eye | -0.0553 | 0.0657 | -0.8423 | 0.3996 |

| | | | | | |
|---|---|---|---|---|---|
| r2_brain | r1_eye | -0.0366 | 0.0891 | -0.4113 | 0.6809 |
| r3_brain | r1_eye | 0.1355 | 0.0677 | 2.0001 | 0.0455 |
| r4_brain | r1_eye | -0.0869 | 0.0906 | -0.9591 | 0.3375 |
| r5_brain | r1_eye | -0.028 | 0.0795 | -0.352 | 0.7248 |
| r6_brain | r1_eye | 0.0311 | 0.0831 | 0.3742 | 0.7083 |
| r1_eye | r1_eye | 1 | 7.61E-07 | 1313651 | 0 |
| r2_eye | r1_eye | 0.8122 | 0.0463 | 17.5482 | 6.14E-69 |
| r3_eye | r1_eye | 0.9167 | 0.018 | 50.8843 | 0 |
| r1_heart | r1_eye | -0.0038 | 0.07 | -0.054 | 0.957 |
| r2_heart | r1_eye | -0.2567 | 0.1138 | -2.2565 | 0.024 |
| r1_brain | r2_eye | -0.0413 | 0.056 | -0.7372 | 0.461 |
| r2_brain | r2_eye | 0.0556 | 0.0713 | 0.7799 | 0.4355 |
| r3_brain | r2_eye | 0.1318 | 0.0617 | 2.1365 | 0.0326 |
| r4_brain | r2_eye | 0.0653 | 0.0762 | 0.8573 | 0.3913 |
| r5_brain | r2_eye | 0.006 | 0.0684 | 0.0884 | 0.9296 |
| r6_brain | r2_eye | 0.009 | 0.0766 | 0.117 | 0.9069 |
| r1_eye | r2_eye | 0.8122 | 0.0463 | 17.5482 | 6.14E-69 |
| r2_eye | r2_eye | 1 | 5.03E-09 | 1.99E+08 | 0 |
| r3_eye | r2_eye | 0.8973 | 0.0267 | 33.5697 | 4.6E-247 |
| r1_heart | r2_eye | 0.0032 | 0.0677 | 0.0476 | 0.962 |
| r2_heart | r2_eye | 0.0168 | 0.115 | 0.1462 | 0.8838 |
| r1_brain | r3_eye | -0.0868 | 0.0622 | -1.3959 | 0.1628 |
| r2_brain | r3_eye | 0.0051 | 0.0688 | 0.0743 | 0.9408 |
| r3_brain | r3_eye | 0.1131 | 0.0603 | 1.8767 | 0.0606 |
| r4_brain | r3_eye | -0.0832 | 0.0809 | -1.029 | 0.3035 |
| r5_brain | r3_eye | -0.0286 | 0.0709 | -0.4042 | 0.6861 |
| r6_brain | r3_eye | 0.0899 | 0.086 | 1.0456 | 0.2957 |
| r1_eye | r3_eye | 0.9167 | 0.018 | 50.8843 | 0 |
| r2_eye | r3_eye | 0.8973 | 0.0267 | 33.5697 | 4.6E-247 |
| r3_eye | r3_eye | 1 | 9.47E-08 | 10562746 | 0 |
| r1_heart | r3_eye | -0.0036 | 0.0684 | -0.053 | 0.9577 |
| r2_heart | r3_eye | -0.1267 | 0.1016 | -1.2472 | 0.2123 |
| r1_brain | r1_heart | -0.1956 | 0.0582 | -3.3592 | 0.0008 |
| r2_brain | r1_heart | -0.0896 | 0.0698 | -1.2836 | 0.1993 |
| r3_brain | r1_heart | -0.082 | 0.0655 | -1.2512 | 0.2109 |
| r4_brain | r1_heart | 0.0371 | 0.0845 | 0.4392 | 0.6605 |
| r5_brain | r1_heart | 0.1822 | 0.066 | 2.7605 | 0.0058 |
| r6_brain | r1_heart | -0.0638 | 0.0957 | -0.6663 | 0.5052 |
| r1_eye | r1_heart | -0.0038 | 0.07 | -0.054 | 0.957 |
| r2_eye | r1_heart | 0.0032 | 0.0677 | 0.0476 | 0.962 |
| r3_eye | r1_heart | -0.0036 | 0.0684 | -0.053 | 0.9577 |
| r1_heart | r1_heart | 1 | 1.06E-07 | 9401192 | 0 |
| r2_heart | r1_heart | 0.4638 | 0.0795 | 5.8373 | 5.31E-09 |
| r1_brain | r2_heart | -0.1571 | 0.0827 | -1.8999 | 0.0574 |
| r2_brain | r2_heart | -0.222 | 0.1088 | -2.0395 | 0.0414 |
| r3_brain | r2_heart | -0.1561 | 0.1105 | -1.4125 | 0.1578 |
| r4_brain | r2_heart | -0.1127 | 0.1154 | -0.9763 | 0.3289 |
| r5_brain | r2_heart | 0.2434 | 0.1024 | 2.3757 | 0.0175 |
| r6_brain | r2_heart | 0.0251 | 0.1127 | 0.2225 | 0.824 |
| r1_eye | r2_heart | -0.2567 | 0.1138 | -2.2565 | 0.024 |
| r2_eye | r2_heart | 0.0168 | 0.115 | 0.1462 | 0.8838 |
| r3_eye | r2_heart | -0.1267 | 0.1016 | -1.2472 | 0.2123 |
| r1_heart | r2_heart | 0.4638 | 0.0795 | 5.8373 | 5.31E-09 |
| r2_heart | r2_heart | 1 | 6.96E-07 | 1437545 | 0 |

#### eTable 8: The genetic correlation between the 11 MAEs and the 9 BAGs

| BAG | MAE | gc_mean | gc_se | Z | P |
|---|---|---|---|---|---|
| Brain_age_gap | r1_brain | 0.1758 | 0.0552 | 3.1836 | 0.0015 |
| Cardiovascular_age_gap | r1_brain | -0.0163 | 0.0434 | -0.3757 | 0.7071 |
| Eye_age_gap | r1_brain | -0.0426 | 0.0517 | -0.8242 | 0.4098 |
| Hepatic_age_gap | r1_brain | -0.0105 | 0.0554 | -0.1904 | 0.849 |
| Immune_age_gap | r1_brain | 0.0918 | 0.0558 | 1.6457 | 0.0998 |
| Metabolic_age_gap | r1_brain | -0.0955 | 0.0485 | -1.9699 | 0.0488 |
| Musculoskeletal_age_gap | r1_brain | 0.0966 | 0.0482 | 2.0051 | 0.045 |
| Pulmonary_age_gap | r1_brain | 0.0295 | 0.045 | 0.6565 | 0.5115 |
| Renal_age_gap | r1_brain | 0.0525 | 0.0391 | 1.3444 | 0.1788 |
| Brain_age_gap | r2_brain | 0.0158 | 0.0713 | 0.2218 | 0.8244 |
| Cardiovascular_age_gap | r2_brain | -0.1038 | 0.0505 | -2.0581 | 0.0396 |
| Eye_age_gap | r2_brain | 0.0244 | 0.095 | 0.2565 | 0.7976 |
| Hepatic_age_gap | r2_brain | -0.0493 | 0.0589 | -0.8378 | 0.4022 |
| Immune_age_gap | r2_brain | 0.111 | 0.0891 | 1.2456 | 0.2129 |
| Metabolic_age_gap | r2_brain | 0.0338 | 0.0618 | 0.5477 | 0.5839 |
| Musculoskeletal_age_gap | r2_brain | 0.0585 | 0.0517 | 1.1303 | 0.2583 |
| Pulmonary_age_gap | r2_brain | 0.0445 | 0.0467 | 0.952 | 0.3411 |
| Renal_age_gap | r2_brain | 0.0578 | 0.0501 | 1.1522 | 0.2492 |
| Brain_age_gap | r3_brain | 0.1555 | 0.049 | 3.1754 | 0.0015 |
| Cardiovascular_age_gap | r3_brain | 0.0087 | 0.0489 | 0.1773 | 0.8593 |
| Eye_age_gap | r3_brain | 0.1093 | 0.0808 | 1.3532 | 0.176 |
| Hepatic_age_gap | r3_brain | -0.0063 | 0.0522 | -0.1207 | 0.9039 |
| Immune_age_gap | r3_brain | -0.0458 | 0.0722 | -0.6351 | 0.5254 |
| Metabolic_age_gap | r3_brain | -0.0257 | 0.0533 | -0.4817 | 0.63 |
| Musculoskeletal_age_gap | r3_brain | 0.0773 | 0.0451 | 1.7119 | 0.0869 |
| Pulmonary_age_gap | r3_brain | 0.0638 | 0.0461 | 1.3859 | 0.1658 |
| Renal_age_gap | r3_brain | 0.0337 | 0.0398 | 0.8457 | 0.3977 |
| Brain_age_gap | r4_brain | -0.1042 | 0.0662 | -1.5734 | 0.1156 |
| Cardiovascular_age_gap | r4_brain | -0.1862 | 0.0571 | -3.2621 | 0.0011 |
| Eye_age_gap | r4_brain | 0.0192 | 0.0813 | 0.2362 | 0.8133 |
| Hepatic_age_gap | r4_brain | -0.0353 | 0.0662 | -0.5335 | 0.5937 |
| Immune_age_gap | r4_brain | 0.0532 | 0.0809 | 0.6573 | 0.511 |
| Metabolic_age_gap | r4_brain | 0.0128 | 0.0659 | 0.1936 | 0.8465 |
| Musculoskeletal_age_gap | r4_brain | -0.012 | 0.0564 | -0.2122 | 0.832 |
| Pulmonary_age_gap | r4_brain | 0.0536 | 0.0538 | 0.9962 | 0.3192 |
| Renal_age_gap | r4_brain | -0.0373 | 0.0511 | -0.7303 | 0.4652 |
| Brain_age_gap | r5_brain | 0.0014 | 0.065 | 0.0219 | 0.9825 |
| Cardiovascular_age_gap | r5_brain | 0.0531 | 0.0514 | 1.0339 | 0.3012 |
| Eye_age_gap | r5_brain | -0.0734 | 0.083 | -0.8836 | 0.3769 |
| Hepatic_age_gap | r5_brain | -0.0695 | 0.0628 | -1.107 | 0.2683 |
| Immune_age_gap | r5_brain | -0.0766 | 0.0769 | -0.996 | 0.3193 |
| Metabolic_age_gap | r5_brain | 0.0506 | 0.0539 | 0.9374 | 0.3486 |
| Musculoskeletal_age_gap | r5_brain | -0.0932 | 0.0467 | -1.9956 | 0.046 |
| Pulmonary_age_gap | r5_brain | -0.0116 | 0.0515 | -0.2248 | 0.8222 |
| Renal_age_gap | r5_brain | 0.0337 | 0.0402 | 0.838 | 0.402 |
| Brain_age_gap | r6_brain | 0.142 | 0.0783 | 1.8143 | 0.0696 |
| Cardiovascular_age_gap | r6_brain | -0.0203 | 0.059 | -0.344 | 0.7309 |
| Eye_age_gap | r6_brain | -0.1937 | 0.0808 | -2.3971 | 0.0165 |
| Hepatic_age_gap | r6_brain | -0.0317 | 0.0754 | -0.421 | 0.6738 |
| Immune_age_gap | r6_brain | -0.0729 | 0.0809 | -0.9008 | 0.3677 |
| Metabolic_age_gap | r6_brain | -0.1638 | 0.0679 | -2.4131 | 0.0158 |
| Musculoskeletal_age_gap | r6_brain | -0.0642 | 0.0588 | -1.093 | 0.2744 |
| Pulmonary_age_gap | r6_brain | -0.0271 | 0.0567 | -0.4777 | 0.6329 |
| Renal_age_gap | r6_brain | -0.0466 | 0.0573 | -0.814 | 0.4156 |
| Brain_age_gap | r1_eye | -0.0128 | 0.0603 | -0.2126 | 0.8317 |
| Cardiovascular_age_gap | r1_eye | -0.0209 | 0.0554 | -0.3778 | 0.7056 |
| Eye_age_gap | r1_eye | 0.3149 | 0.0823 | 3.8247 | 0.0001 |
| Hepatic_age_gap | r1_eye | -0.104 | 0.0676 | -1.5384 | 0.1239 |
| Immune_age_gap | r1_eye | -0.0437 | 0.0802 | -0.5453 | 0.5855 |
| Metabolic_age_gap | r1_eye | 0.1526 | 0.0592 | 2.5757 | 0.01 |
| Musculoskeletal_age_gap | r1_eye | -0.0874 | 0.0505 | -1.7302 | 0.0836 |
| Pulmonary_age_gap | r1_eye | -0.0437 | 0.0483 | -0.9044 | 0.3658 |
| Renal_age_gap | r1_eye | 0.0018 | 0.0473 | 0.0371 | 0.9704 |
| Brain_age_gap | r2_eye | 0.0293 | 0.0585 | 0.5008 | 0.6165 |
| Cardiovascular_age_gap | r2_eye | -0.0745 | 0.0496 | -1.5026 | 0.1329 |
| Eye_age_gap | r2_eye | 0.3837 | 0.0595 | 6.4441 | 1.16E-10 |
| Hepatic_age_gap | r2_eye | -0.018 | 0.0563 | -0.3194 | 0.7494 |

| | | | | | |
|---|---|---|---|---|---|
| Immune_age_gap | r2_eye | 0.0653 | 0.0712 | 0.9162 | 0.3596 |
| Metabolic_age_gap | r2_eye | 0.0709 | 0.0489 | 1.4483 | 0.1475 |
| Musculoskeletal_age_gap | r2_eye | 0.0133 | 0.0435 | 0.3058 | 0.7598 |
| Pulmonary_age_gap | r2_eye | -0.0216 | 0.0403 | -0.5355 | 0.5923 |
| Renal_age_gap | r2_eye | 0.1241 | 0.0472 | 2.6304 | 0.0085 |
| Brain_age_gap | r3_eye | 0.0154 | 0.0563 | 0.2729 | 0.785 |
| Cardiovascular_age_gap | r3_eye | -0.0431 | 0.0493 | -0.8748 | 0.3817 |
| Eye_age_gap | r3_eye | 0.2441 | 0.0749 | 3.2602 | 0.0011 |
| Hepatic_age_gap | r3_eye | -0.0356 | 0.0621 | -0.574 | 0.566 |
| Immune_age_gap | r3_eye | 0.0714 | 0.0673 | 1.0609 | 0.2887 |
| Metabolic_age_gap | r3_eye | 0.1572 | 0.0518 | 3.034 | 0.0024 |
| Musculoskeletal_age_gap | r3_eye | -0.0664 | 0.05 | -1.3289 | 0.1839 |
| Pulmonary_age_gap | r3_eye | -0.0275 | 0.0449 | -0.612 | 0.5405 |
| Renal_age_gap | r3_eye | 0.076 | 0.0476 | 1.5964 | 0.1104 |
| Brain_age_gap | r1_heart | 0.0108 | 0.0645 | 0.1667 | 0.8676 |
| Cardiovascular_age_gap | r1_heart | 0.1825 | 0.0525 | 3.4747 | 0.0005 |
| Eye_age_gap | r1_heart | 0.0066 | 0.0712 | 0.0925 | 0.9263 |
| Hepatic_age_gap | r1_heart | -0.1164 | 0.0637 | -1.828 | 0.0675 |
| Immune_age_gap | r1_heart | -0.13 | 0.066 | -1.9705 | 0.0488 |
| Metabolic_age_gap | r1_heart | 0.0031 | 0.0588 | 0.0524 | 0.9582 |
| Musculoskeletal_age_gap | r1_heart | -0.061 | 0.0541 | -1.1282 | 0.2593 |
| Pulmonary_age_gap | r1_heart | 0.0062 | 0.0484 | 0.1278 | 0.8983 |
| Renal_age_gap | r1_heart | -0.0712 | 0.0506 | -1.4073 | 0.1593 |
| Brain_age_gap | r2_heart | -0.0418 | 0.0774 | -0.5399 | 0.5893 |
| Cardiovascular_age_gap | r2_heart | 0.2763 | 0.0699 | 3.9556 | 7.64E-05 |
| Eye_age_gap | r2_heart | -0.1104 | 0.153 | -0.7216 | 0.4706 |
| Hepatic_age_gap | r2_heart | -0.1138 | 0.1043 | -1.0916 | 0.275 |
| Immune_age_gap | r2_heart | 0.023 | 0.1371 | 0.1679 | 0.8667 |
| Metabolic_age_gap | r2_heart | -0.0719 | 0.0851 | -0.8439 | 0.3987 |
| Musculoskeletal_age_gap | r2_heart | -0.0391 | 0.061 | -0.6408 | 0.5217 |
| Pulmonary_age_gap | r2_heart | 0.0462 | 0.0672 | 0.6876 | 0.4917 |
| Renal_age_gap | r2_heart | -0.0499 | 0.064 | -0.7804 | 0.4352 |

**eTable 9: The results of survival analyses for predicting AD progression**

a) CN→MCI progression

| Hazard_ratio | CI_lower_bound | CI_upper_bound | P_value | MAE | N_case | N_noncase |
|---|---|---|---|---|---|---|
| 0.867305 | 0.69655 | 1.07992 | 0.203144 | r1 | 119 | 496 |
| 0.981331 | 0.724295 | 1.329583 | 0.903204 | r2 | 119 | 496 |
| 1.302353 | 0.892186 | 1.901087 | 0.171049 | r3 | 119 | 496 |
| 1.10269 | 0.86426 | 1.406898 | 0.431639 | r4 | 119 | 496 |
| 0.909696 | 0.75721 | 1.092889 | 0.311981 | r5 | 119 | 496 |
| 1.018401 | 0.776977 | 1.334841 | 0.894923 | r6 | 119 | 496 |

b) MCI→AD progression

| Hazard_ratio | CI_lower_bound | CI_upper_bound | P_value | MAE | N_case | N_noncase |
|---|---|---|---|---|---|---|
| 1.334182 | 1.149922 | 1.547967 | 0.000143 | r1 | 326 | 649 |
| 1.376578 | 1.081748 | 1.751764 | 0.009351 | r2 | 326 | 649 |
| 1.858877 | 1.406245 | 2.457198 | 1.33E-05 | r3 | 326 | 649 |
| 0.848032 | 0.716702 | 1.003426 | 0.054843 | r4 | 326 | 649 |
| 0.696539 | 0.62055 | 0.781834 | 8.48E-10 | r5 | 326 | 649 |
| 0.863292 | 0.724296 | 1.028961 | 0.100752 | r6 | 326 | 649 |

**eTable 10: The results of survival analyses for predicting mortality**

| hazard_ratio | CI_lower_bound | CI_upper_bound | p_value | MAE | N_case | N_noncase | Organ |
|---|---|---|---|---|---|---|---|
| 0.925493 | 0.831076 | 1.030636 | 0.158442 | r1_prs | 333 | 33237 | brain |
| 1.053108 | 0.945236 | 1.173291 | 0.347991 | r2_prs | 333 | 33237 | brain |
| 1.071036 | 0.962275 | 1.192089 | 0.209078 | r3_prs | 333 | 33237 | brain |
| 1.045934 | 0.940007 | 1.163798 | 0.40974 | r4_prs | 333 | 33237 | brain |
| 0.964838 | 0.867484 | 1.073118 | 0.509511 | r5_prs | 333 | 33237 | brain |
| 0.982948 | 0.883448 | 1.093654 | 0.752109 | r6_prs | 333 | 33237 | brain |
| 1.057039 | 0.950121 | 1.175989 | 0.307943 | r1 | 333 | 33237 | brain |
| 1.051421 | 0.944266 | 1.170736 | 0.360558 | r2 | 333 | 33237 | brain |
| 1.143454 | 1.028798 | 1.270888 | 0.012897 | r3 | 333 | 33237 | brain |
| 0.992024 | 0.891374 | 1.104038 | 0.883357 | r4 | 333 | 33237 | brain |
| 0.986813 | 0.886696 | 1.098233 | 0.807837 | r5 | 333 | 33237 | brain |
| 0.969423 | 0.870926 | 1.079059 | 0.569987 | r6 | 333 | 33237 | brain |
| 0.997906 | 0.957916 | 1.039564 | 0.919966 | r1_prs | 2321 | 34333 | eye |
| 1.04078 | 0.998747 | 1.084582 | 0.057384 | r2_prs | 2321 | 34333 | eye |
| 1.027301 | 0.986096 | 1.070229 | 0.197192 | r3_prs | 2321 | 34333 | eye |
| 1.034517 | 0.995847 | 1.074689 | 0.080836 | r1 | 2321 | 34333 | eye |
| 1.04512 | 1.006555 | 1.085163 | 0.021415 | r2 | 2321 | 34333 | eye |
| 1.023184 | 0.982542 | 1.065508 | 0.26773 | r3 | 2321 | 34333 | eye |
| 1.037233 | 0.956846 | 1.124375 | 0.374439 | r1_prs | 568 | 29682 | heart |
| 1.091559 | 1.010858 | 1.178702 | 0.025381 | r2_prs | 568 | 29682 | heart |
| 1.142798 | 1.055885 | 1.236866 | 0.000942 | r1 | 568 | 29682 | heart |
| 1.207107 | 1.124397 | 1.295901 | 2.02E-07 | r2 | 568 | 29682 | heart |

**eTable 11: Preclinical AD drug outcome (PACC) is linked to Brain 1-3**

Mean PACC group change from baseline at week 240 by treatment group estimated from the spline model. For fair comparison, the placebo group was also stratified by median MAE.

**A**): Over-expressed vs. Under-expressed

| Contrast (at week 240) | Estimate | SE | df | t-statistic | P-value |
|---|---|---|---|---|---|
| Brain 1 Over-expressed - Under-expressed | -0.984 | 0.407 | 5130 | -2.414 | 0.0158 |
| Brain 2 Over-expressed - Under-expressed | -1.41 | 0.4 | 5130 | -3.519 | 0.0004 |
| Brain 3 Over-expressed - Under-expressed | -0.92 | 0.408 | 5130 | -2.254 | 0.0242 |

**B**): Placebo - Over-expressed

| Contrast (at week 240) | Estimate | SE | df | t-statistic | P-value |
|---|---|---|---|---|---|
| Placebo - Brain 1 Over-expressed | 0.112 | 0.401 | 5073 | 0.278 | 0.7807 |
| Placebo - Brain 2 Over-expressed | 0.275 | 0.405 | 5121 | 0.679 | 0.4972 |
| Placebo - Brain 3 Over-expressed | -0.0296 | 0.396 | 5108 | -0.075 | 0.9403 |

**C**): Placebo - Under-expressed

| Contrast (at week 240) | Estimate | SE | df | t-statistic | P-value |
|---|---|---|---|---|---|
| Placebo - Brain 1 Under-expressed | 0.285 | 0.336 | 5367 | 0.848 | 0.3969 |
| Placebo - Brain 2 Under-expressed | 0.0374 | 0.325 | 5384 | 0.115 | 0.9084 |
| Placebo - Brain 3 Under-expressed | 0.494 | 0.341 | 5315 | 1.447 | 0.1480 |

**eTable 12: The 128 brain PSCs, 80 heart IDPs, and 64 eye IDPs included to derive the 11 MAEs via Surreal-GAN**

a) **128 Brain PSCs**: The brain PSCs originated from our prior investigation utilizing six scales of the MuSIC atlas[5] (C=32, 64, 128, 256, 512, and 1024, with 13 PSCs at C1024 omitted during optimization). We used the scale of C=128. The visualization of each PSC can be found here: https://labs-laboratory.com/bridgeport.

| Scale (C) | Number of PSCs | Name |
|---|---|---|
| 128 | 128 | C128_*N* |

b) **Heart IDPs**: After QC, the 80 heart IDPs were downloaded directly from UKBB. On the UKBB showcase website, we present the full names of the heart IDPs, Field ID, abbreviations, and high-level groups. An explanation for each heart IDP can be found here: https://labs-laboratory.com/medicine/cardiovascular.

| IDP | Abb | Group | Field ID | Heart field |
|---|---|---|---|---|
| lv_end_diastolic_volume_f24100_2_0 | LVEDV | LV | 24100 | left_ventricle |
| lv_end_systolic_volume_f24101_2_0 | LVESV | LV | 24101 | left_ventricle |
| lv_stroke_volume_f24102_2_0 | LVSV | LV | 24102 | left_ventricle |
| lv_ejection_fraction_f24103_2_0 | LVEF | LV | 24103 | left_ventricle |
| lv_cardiac_output_f24104_2_0 | LVCO | LV | 24104 | left_ventricle |
| lv_myocardial_mass_f24105_2_0 | LVM | LV | 24105 | left_ventricle |
| rv_end_diastolic_volume_f24106_2_0 | RVEDV | RV | 24106 | right_ventricle |
| rv_end_systolic_volume_f24107_2_0 | RVESV | RV | 24107 | right_ventricle |
| rv_stroke_volume_f24108_2_0 | RVSV | RV | 24108 | right_ventricle |
| rv_ejection_fraction_f24109_2_0 | RVEF | RV | 24109 | right_ventricle |
| la_maximum_volume_f24110_2_0 | LAV_max | LA | 24110 | left_atrium |
| la_minimum_volume_f24111_2_0 | LAV_min | LA | 24111 | left_atrium |
| la_stroke_volume_f24112_2_0 | LASV | LA | 24112 | left_atrium |
| la_ejection_fraction_f24113_2_0 | LAEF | LA | 24113 | left_atrium |
| ra_maximum_volume_f24114_2_0 | RAV_max | RA | 24114 | left_atrium |
| ra_minimum_volume_f24115_2_0 | RAV_min | RA | 24115 | left_atrium |
| ra_stroke_volume_f24116_2_0 | RASV | RA | 24116 | left_atrium |
| ra_ejection_fraction_f24117_2_0 | RAEF | RA | 24117 | left_atrium |
| ascending_aorta_maximum_area_f24118_2_0 | Aao_max | AA | 24118 | ascending_aorta |
| ascending_aorta_minimum_area_f24119_2_0 | Aao_min | AA | 24119 | ascending_aorta |
| descending_aorta_maximum_area_f24121_2_0 | Dao_max | DA | 24121 | descending_aorta |
| descending_aorta_minimum_area_f24122_2_0 | Dao_min | DA | 24122 | descending_aorta |
| lv_mean_myocardial_wall_thickness_aha_1_f24124_2_0 | WT_AHA_1 | LV | 24124 | left_ventricle |

| | | | | |
|---|---|---|---|---|
| lv_mean_myocardial_wall_thickness_aha_2_f24125_2_0 | WT_AHA_2 | LV | 24125 | left_ventricle |
| lv_mean_myocardial_wall_thickness_aha_3_f24126_2_0 | WT_AHA_3 | LV | 24126 | left_ventricle |
| lv_mean_myocardial_wall_thickness_aha_4_f24127_2_0 | WT_AHA_4 | LV | 24127 | left_ventricle |
| lv_mean_myocardial_wall_thickness_aha_5_f24128_2_0 | WT_AHA_5 | LV | 24128 | left_ventricle |
| lv_mean_myocardial_wall_thickness_aha_6_f24129_2_0 | WT_AHA_6 | LV | 24129 | left_ventricle |
| lv_mean_myocardial_wall_thickness_aha_7_f24130_2_0 | WT_AHA_7 | LV | 24130 | left_ventricle |
| lv_mean_myocardial_wall_thickness_aha_8_f24131_2_0 | WT_AHA_8 | LV | 24131 | left_ventricle |
| lv_mean_myocardial_wall_thickness_aha_9_f24132_2_0 | WT_AHA_9 | LV | 24132 | left_ventricle |
| lv_mean_myocardial_wall_thickness_aha_10_f24133_2_0 | WT_AHA_10 | LV | 24133 | left_ventricle |
| lv_mean_myocardial_wall_thickness_aha_11_f24134_2_0 | WT_AHA_11 | LV | 24134 | left_ventricle |
| lv_mean_myocardial_wall_thickness_aha_12_f24135_2_0 | WT_AHA_12 | LV | 24135 | left_ventricle |
| lv_mean_myocardial_wall_thickness_aha_13_f24136_2_0 | WT_AHA_13 | LV | 24136 | left_ventricle |
| lv_mean_myocardial_wall_thickness_aha_14_f24137_2_0 | WT_AHA_14 | LV | 24137 | left_ventricle |
| lv_mean_myocardial_wall_thickness_aha_15_f24138_2_0 | WT_AHA_15 | LV | 24138 | left_ventricle |
| lv_mean_myocardial_wall_thickness_aha_16_f24139_2_0 | WT_AHA_16 | LV | 24139 | left_ventricle |
| lv_mean_myocardial_wall_thickness_global_f24140_2_0 | WT_global | LV | 24140 | left_ventricle |
| lv_circumferential_strain_aha_1_f24141_2_0 | Ecc_AHA_1 | LV | 24141 | left_ventricle |
| lv_circumferential_strain_aha_2_f24142_2_0 | Ecc_AHA_2 | LV | 24142 | left_ventricle |
| lv_circumferential_strain_aha_3_f24143_2_0 | Ecc_AHA_3 | LV | 24143 | left_ventricle |
| lv_circumferential_strain_aha_4_f24144_2_0 | Ecc_AHA_4 | LV | 24144 | left_ventricle |
| lv_circumferential_strain_aha_5_f24145_2_0 | Ecc_AHA_5 | LV | 24145 | left_ventricle |
| lv_circumferential_strain_aha_6_f24146_2_0 | Ecc_AHA_6 | LV | 24146 | left_ventricle |
| lv_circumferential_strain_aha_7_f24147_2_0 | Ecc_AHA_7 | LV | 24147 | left_ventricle |
| lv_circumferential_strain_aha_8_f24148_2_0 | Ecc_AHA_8 | LV | 24148 | left_ventricle |
| lv_circumferential_strain_aha_9_f24149_2_0 | Ecc_AHA_9 | LV | 24149 | left_ventricle |
| lv_circumferential_strain_aha_10_f24150_2_0 | Ecc_AHA_10 | LV | 24150 | left_ventricle |
| lv_circumferential_strain_aha_11_f24151_2_0 | Ecc_AHA_11 | LV | 24151 | left_ventricle |
| lv_circumferential_strain_aha_12_f24152_2_0 | Ecc_AHA_12 | LV | 24152 | left_ventricle |
| lv_circumferential_strain_aha_13_f24153_2_0 | Ecc_AHA_13 | LV | 24153 | left_ventricle |
| lv_circumferential_strain_aha_14_f24154_2_0 | Ecc_AHA_14 | LV | 24154 | left_ventricle |
| lv_circumferential_strain_aha_15_f24155_2_0 | Ecc_AHA_15 | LV | 24155 | left_ventricle |
| lv_circumferential_strain_aha_16_f24156_2_0 | Ecc_AHA_16 | LV | 24156 | left_ventricle |
| lv_circumferential_strain_global_f24157_2_0 | Ecc_global | LV | 24157 | left_ventricle |
| lv_radial_strain_aha_1_f24158_2_0 | Err_AHA_1 | LV | 24158 | left_ventricle |
| lv_radial_strain_aha_2_f24159_2_0 | Err_AHA_2 | LV | 24159 | left_ventricle |

| | | | | |
|---|---|---|---|---|
| lv_radial_strain_aha_3_f24160_2_0 | Err_AHA_3 | LV | 24160 | left_ventricle |
| lv_radial_strain_aha_4_f24161_2_0 | Err_AHA_4 | LV | 24161 | left_ventricle |
| lv_radial_strain_aha_5_f24162_2_0 | Err_AHA_5 | LV | 24162 | left_ventricle |
| lv_radial_strain_aha_6_f24163_2_0 | Err_AHA_6 | LV | 24163 | left_ventricle |
| lv_radial_strain_aha_7_f24164_2_0 | Err_AHA_7 | LV | 24164 | left_ventricle |
| lv_radial_strain_aha_8_f24165_2_0 | Err_AHA_8 | LV | 24165 | left_ventricle |
| lv_radial_strain_aha_9_f24166_2_0 | Err_AHA_9 | LV | 24166 | left_ventricle |
| lv_radial_strain_aha_10_f24167_2_0 | Err_AHA_10 | LV | 24167 | left_ventricle |
| lv_radial_strain_aha_11_f24168_2_0 | Err_AHA_11 | LV | 24168 | left_ventricle |
| lv_radial_strain_aha_12_f24169_2_0 | Err_AHA_12 | LV | 24169 | left_ventricle |
| lv_radial_strain_aha_13_f24170_2_0 | Err_AHA_13 | LV | 24170 | left_ventricle |
| lv_radial_strain_aha_14_f24171_2_0 | Err_AHA_14 | LV | 24171 | left_ventricle |
| lv_radial_strain_aha_15_f24172_2_0 | Err_AHA_15 | LV | 24172 | left_ventricle |
| lv_radial_strain_aha_16_f24173_2_0 | Err_AHA_16 | LV | 24173 | left_ventricle |
| lv_radial_strain_global_f24174_2_0 | Err_global | LV | 24174 | left_ventricle |
| lv_longitudinal_strain_segment_1_f24175_2_0 | Ell_1 | LV | 24175 | left_ventricle |
| lv_longitudinal_strain_segment_2_f24176_2_0 | Ell_2 | LV | 24176 | left_ventricle |
| lv_longitudinal_strain_segment_3_f24177_2_0 | Ell_3 | LV | 24177 | left_ventricle |
| lv_longitudinal_strain_segment_4_f24178_2_0 | Ell_4 | LV | 24178 | left_ventricle |
| lv_longitudinal_strain_segment_5_f24179_2_0 | Ell_5 | LV | 24179 | left_ventricle |
| lv_longitudinal_strain_segment_6_f24180_2_0 | Ell_6 | LV | 24180 | left_ventricle |
| lv_longitudinal_strain_global_f24181_2_0 | Ell_global | LV | 24181 | left_ventricle |

c) **Eye IDPs**: After QC, the 64 eye IDPs were downloaded directly from UKBB. We present the full name of the eye IDPs, Field ID on the UKBB showcase website, abbreviations, high-level groups, and PubMed ID. An explanation for each eye IDP can be found here: https://labs-laboratory.com/medicine/eye.

| **Eye IDP** | **Field ID** | **Abb** | **Group** | **Pubmed** |
|---|---|---|---|---|
| overall_macular_thickness_left_f27800_0_0 | 27800 | ML | MT | 26746598 |
| overall_macular_thickness_right_f27801_0_0 | 27801 | MR | MT | 26746598 |
| macular_thickness_at_the_central_subfield_left_f27802_0_0 | 27802 | McL | MT | 26746598 |
| macular_thickness_at_the_central_subfield_right_f27803_0_0 | 27803 | McR | MT | 26746598 |
| macular_thickness_at_the_inner_inferior_subfield_left_f27804_0_0 | 27804 | MiiL | MT | 26746598 |
| macular_thickness_at_the_inner_inferior_subfield_right_f27805_0_0 | 27805 | MiiR | MT | 26746598 |
| macular_thickness_at_the_inner_nasal_subfield_left_f27806_0_0 | 27806 | MinL | MT | 26746598 |
| macular_thickness_at_the_inner_nasal_subfield_right_f27807_0_0 | 27807 | MinR | MT | 26746598 |
| macular_thickness_at_the_inner_superior_subfield_left_f27808_0_0 | 27808 | MisL | MT | 26746598 |
| macular_thickness_at_the_inner_superior_subfield_right_f27809_0_0 | 27809 | MisR | MT | 26746598 |
| macular_thickness_at_the_inner_temporal_subfield_left_f27810_0_0 | 27810 | MitL | MT | 26746598 |
| macular_thickness_at_the_inner_temporal_subfield_right_f27811_0_0 | 27811 | MitR | MT | 26746598 |

| | | | | |
|---|---|---|---|---|
| macular_thickness_at_the_outer_inferior_subfield_left_f27812_0_0 | 27812 | MoiL | MT | 26746598 |
| macular_thickness_at_the_outer_inferior_subfield_right_f27813_0_0 | 27813 | MoiR | MT | 26746598 |
| macular_thickness_at_the_outer_nasal_subfield_left_f27814_0_0 | 27814 | MonL | MT | 26746598 |
| macular_thickness_at_the_outer_nasal_subfield_right_f27815_0_0 | 27815 | MonR | MT | 26746598 |
| macular_thickness_at_the_outer_superior_subfield_left_f27816_0_0 | 27816 | MosL | MT | 26746598 |
| macular_thickness_at_the_outer_superior_subfield_right_f27817_0_0 | 27817 | MosR | MT | 26746598 |
| macular_thickness_at_the_outer_temporal_subfield_left_f27818_0_0 | 27818 | MotL | MT | 26746598 |
| macular_thickness_at_the_outer_temporal_subfield_right_f27819_0_0 | 27819 | MotR | MT | 26746598 |
| average_retinal_nerve_fibre_layer_thickness_left_f28500_0_0 | 28500 | RnfL | RNF | 26746598 |
| average_retinal_nerve_fibre_layer_thickness_right_f28501_0_0 | 28501 | RnfR | RNF | 26746598 |
| average_inner_nuclear_layer_thickness_left_f28502_0_0 | 28502 | InlL | INL | 26746598 |
| average_inner_nuclear_layer_thickness_right_f28503_0_0 | 28503 | InlR | INL | 26746598 |
| average_ganglion_cellinner_plexiform_layer_thickness_left_f28504_0_0 | 28504 | GcplL | GCPL | 26746598 |
| average_ganglion_cellinner_plexiform_layer_thickness_right_f28505_0_0 | 28505 | GcplR | GCPL | 26746598 |
| inlelm_thickness_of_the_central_subfield_left_f28506_0_0 | 28506 | InlElmCL | INL/ELM | 26746598 |
| inlelm_thickness_of_the_central_subfield_right_f28507_0_0 | 28507 | InlElmCR | INL/ELM | 26746598 |
| inlelm_thickness_of_the_inner_subfield_left_f28508_0_0 | 28508 | InlElmIL | INL/ELM | 26746598 |
| inlelm_thickness_of_the_inner_subfield_right_f28509_0_0 | 28509 | InlElmIR | INL/ELM | 26746598 |
| inlelm_thickness_of_the_outer_subfield_left_f28510_0_0 | 28510 | InlElmOL | INL/ELM | 26746598 |
| inlelm_thickness_of_the_outer_subfield_right_f28511_0_0 | 28511 | InlElmOR | INL/ELM | 26746598 |
| average_inlelm_thickness_left_f28512_0_0 | 28512 | InlElmL | INL/ELM | 26746598 |
| average_inlelm_thickness_right_f28513_0_0 | 28513 | InlElmR | INL/ELM | 26746598 |
| elmisos_thickness_of_central_subfield_left_f28514_0_0 | 28514 | ElmIsosCL | ELM/ISOS | 26746598 |
| elmisos_thickness_of_central_subfield_right_f28515_0_0 | 28515 | ElmIsosCR | ELM/ISOS | 26746598 |
| elmisos_thickness_of_inner_subfield_left_f28516_0_0 | 28516 | ElmIsosIL | ELM/ISOS | 26746598 |
| elmisos_thickness_of_inner_subfield_right_f28517_0_0 | 28517 | ElmIsosIR | ELM/ISOS | 26746598 |
| elmisos_thickness_of_outer_subfield_left_f28518_0_0 | 28518 | ElmIsosOL | ELM/ISOS | 26746598 |
| elmisos_thickness_of_outer_subfield_right_f28519_0_0 | 28519 | ElmIsosOR | ELM/ISOS | 26746598 |
| average_elmisos_thickness_left_f28520_0_0 | 28520 | ElmIsosL | ELM/ISOS | 26746598 |
| average_elmisos_thickness_right_f28521_0_0 | 28521 | ElmIsosR | ELM/ISOS | 26746598 |
| isosrpe_thickness_of_central_subfield_left_f28522_0_0 | 28522 | IsosRpeCL | ISOS/RPE | 26746598 |
| isosrpe_thickness_of_central_subfield_right_f28523_0_0 | 28523 | IsosRpeCR | ISOS/RPE | 26746598 |
| isosrpe_thickness_of_inner_subfield_left_f28524_0_0 | 28524 | IsosRpeIL | ISOS/RPE | 26746598 |
| isosrpe_thickness_of_inner_subfield_right_f28525_0_0 | 28525 | IsosRpeIR | ISOS/RPE | 26746598 |
| isosrpe_thickness_of_outer_subfield_left_f28526_0_0 | 28526 | IsosRpeOL | ISOS/RPE | 26746598 |
| isosrpe_thickness_of_outer_subfield_right_f28527_0_0 | 28527 | IsosRpeOR | ISOS/RPE | 26746598 |
| average_isosrpe_thickness_left_f28528_0_0 | 28528 | IsosRpeL | ISOS/RPE | 26746598 |
| average_isosrpe_thickness_right_f28529_0_0 | 28529 | IsosRpeR | ISOS/RPE | 26746598 |
| inlrpe_thickness_of_central_subfield_left_f28530_0_0 | 28530 | InlRpeCL | INL/RPE | 26746598 |
| inlrpe_thickness_of_central_subfield_right_f28531_0_0 | 28531 | InlRpeCR | INL/RPE | 26746598 |

| | | | | |
|---|---|---|---|---|
| inlrpe_thickness_of_inner_subfield_left_f28532_0_0 | 28532 | InlRpeIL | INL/RPE | 26746598 |
| inlrpe_thickness_of_inner_subfield_right_f28533_0_0 | 28533 | InlRpeIR | INL/RPE | 26746598 |
| inlrpe_thickness_of_outer_subfield_left_f28534_0_0 | 28534 | InlRpeOL | INL/RPE | 26746598 |
| inlrpe_thickness_of_outer_subfield_right_f28535_0_0 | 28535 | InlRpeOR | INL/RPE | 26746598 |
| average_inlrpe_thickness_left_f28536_0_0 | 28536 | InlRpeL | INL/RPE | 26746598 |
| average_inlrpe_thickness_right_f28537_0_0 | 28537 | InlRpeR | INL/RPE | 26746598 |
| overall_average_retinal_pigment_epithelium_thickness_left_f27822_0_0 | 27822 | RpeL | RPE | 26746598 |
| overall_average_retinal_pigment_epithelium_thickness_right_f27823_0_0 | 27823 | RpeR | RPE | 26746598 |
| disc_diameter_after_inverse_rank_normal_transformation_left_f27851_0_0 | 27851 | DdL | DISC | 31809533 |
| mean_of_vertical_disc_diameter_left_f27853_0_0 | 27853 | VdiscL | DISC | 31809533 |
| vertical_cup_to_disc_ratio_vcdr_regressed_and_transformed_left_f27855_0_0 | 27855 | VcdrTL | DISC | 31809533 |
| vertical_cup_to_disc_ratio_vcdr_left_f27857_0_0 | 27857 | VcdrL | DISC | 31809533 |

**eFile 1-19: The Online Supplementary files contain large tables**

This was submitted to the MTS as a zip file: “eFile-20250619T174905Z-1-001.zip”

**eFolder 1: Sensitivity analyses for the MR analyses**

This was submitted to the MTS as a zip file: “eFolder-20250619T174808Z-1-001.zip”

**GWAS summary statistics**

We updated our MEDICINE page for the 11 MAEs (https://labs-laboratory.com/medicine/) and shared the GWAS summary data on Google Drive: https://drive.google.com/drive/u/1/folders/1wQdWv9FqZEBXQQ0p5bgTjYAXeSgQUmZP; we will make this publicly available after the paper's acceptance.
